# Genomics-enhanced contact tracing enabled the characterization of SARS-CoV-2 transmission chains and infection contexts in the general population during community transmission

**DOI:** 10.1101/2025.06.13.25329413

**Authors:** Johannes Ptok, Jonas Weber, Moritz Pigulla, Coco Röhl, Nils Lüschow, Dirk Nagels, Torsten Houwaart, Samir Belhaj, Lena-Sophie Schneider, Dominik Regorz, Pascal Kreuzer, Yara Fröhlich, Patrick Finzer, Emran Tawalbeh, Jessica Nicolai, Mygg Stiller, Jacqueline Blum, Christian Lange, Roland Geisel, Daniel Wind, Lisanna Hülse, Alona Tysha, Tobias Wienemann, Malte Kohns Vasconcelos, Katrin Hoffmann, Nadine Lübke, Sandra Hauka, Klaus Pfeffer, Jörg Timm, Lutz Ehlkes, Andreas Walker, Alexander T. Dilthey

## Abstract

Understanding pathogen transmission is key to effective infection prevention. From February to December 2021, we implemented genomics-enhanced contact tracing for SARS-CoV-2 in Düsseldorf, Germany, integrating data on 32,380 cases, 49,906 contact tracing records, 162 outbreaks, and 8,028 viral genomes (sequencing coverage 24.5%). Combining epidemiological and genetic data, we found a putative infection source for 19% of sequenced and 44% of all cases. Household-associated transmission accounted for up to 40% of all cases; classical contact tracing had limited sensitivity for non-household contacts, and gastronomy, hospital, school and kindergarten contexts were genetically found to be likely enriched for undetected transmissions. Outbreaks were associated with 8% of cases; school, kindergarten and nightlife outbreaks were strongly connected to the community, with nightlife outbreaks showing a strong post-outbreak increase in genetically associated cases. Sequencing detected previously unrecognized links between school outbreaks and 18% additional outbreak-associated sequenced cases. In conclusion, in addition to classical contact tracing, SARS-CoV-2 sequencing was required to achieve an improved resolution of transmission dynamics; future implementations of genomics-enhanced contact tracing should aim for sequencing rates of at least 15% to enable effective characterization of infection contexts and outbreaks.

## Introduction

Understanding when, where and between whom pathogens are transmitted is the foundational question of infectious disease epidemiology. Knowledge of pathogen transmission patterns can contribute to the implementation of effective infection prevention measures. However, during community transmission, the structure of transmission chains and of the structural factors driving them – such as the contribution of outbreaks – often remains elusive to both the affected individual and the public health system. Reasons for this include the fact that there are often multiple potential sources for each individual infection event, which are not easily disambiguated; that some transmissions take place between individuals not known to each other, limiting the efficacy of classical contact tracing; and resource constraints at the level of public health systems, which limit the ability to comprehensively track and investigate transmission chains and their associated infection contexts at the population level.

Pathogen genome sequencing can play an important role in the characterization of pathogen transmission [Harris et al. 2013, Andersen et al. 2015, Mate et al. 2015, Faria et al. 2016, Mellmann et al. 2016, COVID-19 Genomics UK (COG-UK) 2020, Lane et al. 2021, Hjorleifsson et al. 2022, McCrone et al. 2022]. When combined with classical contact tracing, pathogen genome sequencing can enable the tracking of individual transmission chains in the population during community transmission (“genomics-enhanced contact tracing”; Seemann et al. [2020], Hjorleifsson et al. [2022], Walker et al. [2022], Sivertsen et al. [2024], Bludau et al. [2025]). In this context, pathogen genome sequencing can be used to validate or disambiguate epidemiological links detected by routine contact tracing; enable the discovery of links missed by routine contact tracing by directing in-depth epidemiological investigation efforts (e.g., based on in-depth case interviews) to case pairs with otherwise unexplained high genetic similarity [Houwaart et al. 2022]; and contribute to the characterization of large-scale transmission patterns, such as quantifying the overall impact of outbreaks. The implementation of genomics-enhanced contact tracing requires high contact tracing and case sequencing rates; of note, neither pathogen sequencing by itself nor other modeling approaches (e.g. of individual-level mobility patterns, Pullano et al. [2020]) are sufficient to enable the tracking of individual transmission chains.

In 2021, the city of Düsseldorf, the capital of Germany’s most populous federal state, implemented a system for genomics-enhanced contact tracing of SARS-CoV-2, which combined best-practice classical contact tracing with one of the highest-intensity SARS-CoV-2 genomic surveillance regimes in continental Europe [Houwaart et al. 2022, Walker et al. 2022]. Here, we report on the first retrospective analysis of the complete dataset (Fig. 1), with the specific aim of characterizing to which extent the integration of genetic data and classical contact tracing can enable insights into SARS-CoV-2 transmission that would not be obtained from classical contact tracing alone.

**Figure 1:**
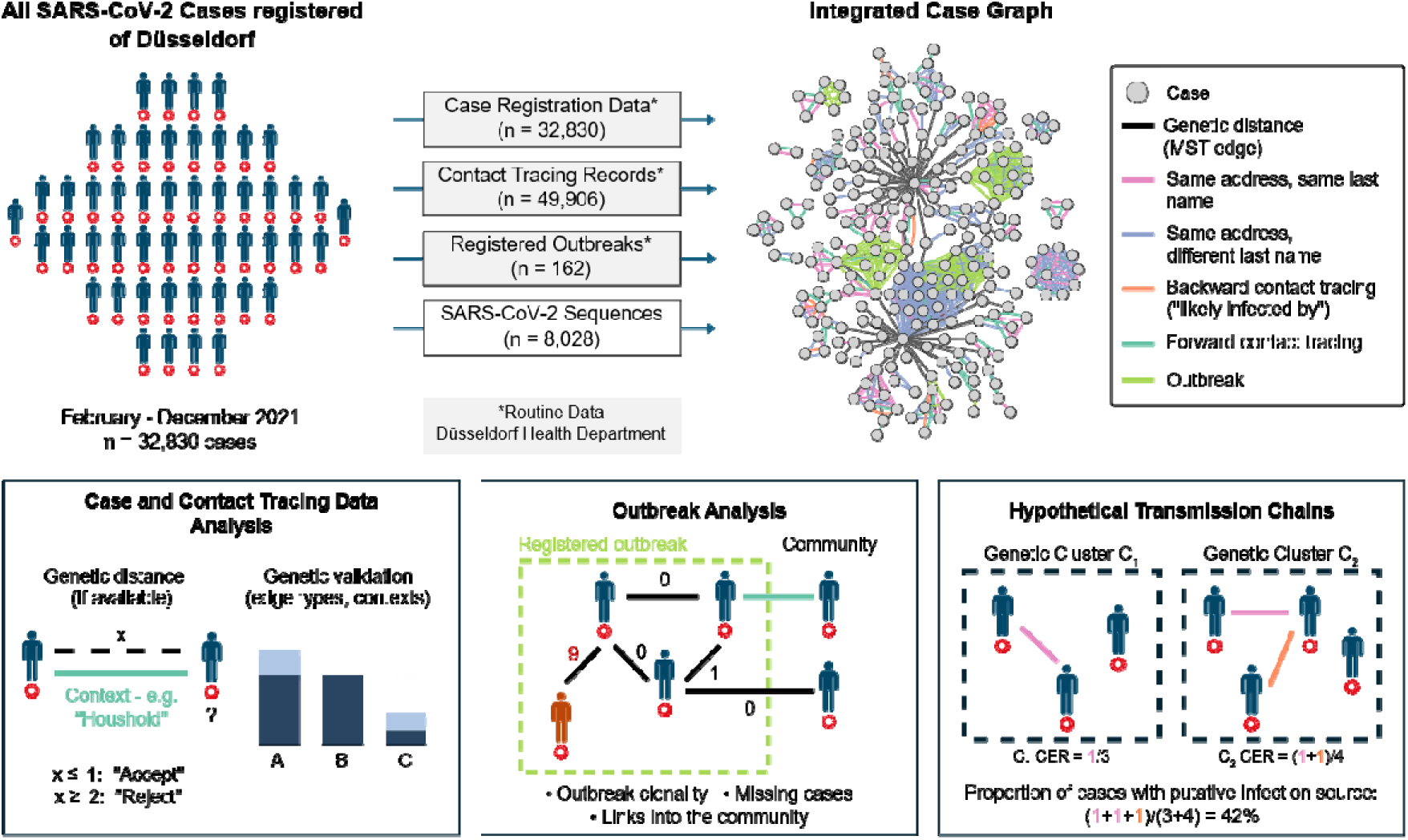
Characterization of the community transmission of SARS-CoV-2 based on genomics-enhanced contact tracing. Cases and their epidemiological connections, determined based on Düsseldorf Health Authority data, were represented in the form of an Integrated Case Graph. Genetic distance edges between sequenced cases are included for illustrative purposes; pairwise genetic distances are not stored as part of the graph. Based on the integrated dataset, we mapped epidemiological connections; investigated all registered institutional outbreaks; and attempted to identify a putative infection source for each registered case through hypothetical transmission chains (CER: cluster explanation rate). Across analyses, we used genetic data to evaluate whether epidemiological links between cases were likely associated with transmission.

## Results

### Implementation of genomics-enhanced contact tracing of SARS-CoV-2 in Düsseldorf

Between February and December 2021, the city of Düsseldorf implemented genomics-enhanced contact tracing for SARS-CoV-2. Best-practice classical contact tracing for all diagnosed SARS-CoV-2 cases from Düsseldorf was carried out by Düsseldorf Health Authority based on guidelines by Germany’s Robert Koch Institute, involving (i) the registration of all detected cases and their personal data (n = 32,830 cases); (ii) contact tracing for all registered cases, with a minimum of three contact attempts using available landline or mobile phone numbers by Düsseldorf Health Authority personnel before contact tracing was deemed unsuccessful (n = 48,131 forward and n = 875 backward contact tracing records; Supplementary Figure 1 and Supplementary Figure 2); and (iii) the registration and investigation of suspected outbreaks in institutional settings such as schools (n = 162 outbreaks with 919 associated cases). Rapid SARS-CoV-2 sequencing for as many local SARS-CoV-2 isolates as possible (n = 11,349 collected viral genome sequences, of which n = 8,763 passed quality control with <= 3,000 ambiguous or undefined nucleotides; Fig. 2A; Supplementary Table 1) was carried out in collaboration between two large commercial diagnostic labs, Düsseldorf Health Authority, and Heinrich Heine University [Houwaart et al. 2022, Walker et al. 2022]. The median turnaround time from sample arrival at Heinrich Heine University to availability of a first sequence was 63 hours (tracked for a subset of the samples collected between April and June 2021; Supplementary Fig. 3). All personally identifiable data remained at Düsseldorf Health Authority.

**Figure 2:**
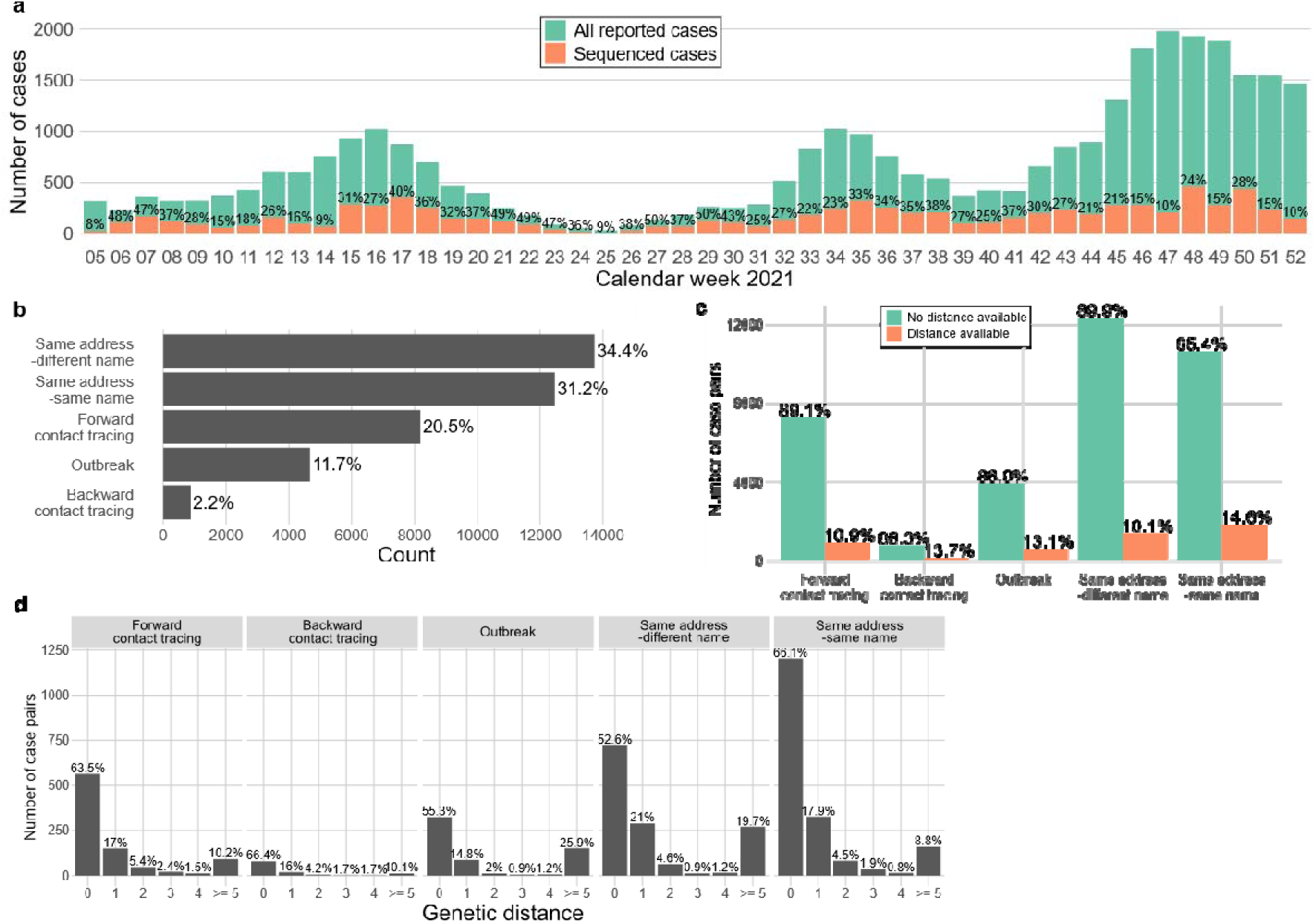
SARS-CoV-2 sequencing and epidemiological connections represented in the Integrated Case Graph. **a,** Shown is the number of registered and sequenced SARS-CoV-2 cases in Düsseldorf on a weekly basis between February and December 2021. **b,** Edges in the Integrated Case Graph, stratified by edge type. **c,** Number of case pairs connected by different types of links in the Integrated Case Graph; shown in red is the proportion of case pairs for which a genetic distance was, based on the sequencing status of the corresponding cases, available. **d,** Distribution of SARS-CoV-2 genetic distances for case pairs linked by different types of edges in the Integrated Case Graph, stratified by edge type and only shown for case pairs for which a genetic distance was available. **e,** Infection contexts captured by forward contact tracing (case pairs with genetic distances only). Shown is the number of case pairs that were connected by a forward contact tracing edge assigned to the corresponding putative context, stratified by whether the genetic distance between the connected cases was (“accepted”) or (“rejected”).

### Unified representation of all epidemiological and genomic data in the Integrated Case Graph

We developed a scalable data integration approach to enable the analysis of the collected data. Case, contact tracing and pathogen genome sequencing data were represented in the form of an Integrated Case Graph (Fig. 1), in which cases were represented as nodes (n = 32,830 case nodes); epidemiological connections as edges (n = 39,927 links between pairs of cases); and in which pathogen genome sequences (n = 8,028 viral genome sequences after quality control and record linkage) were attached to the nodes of their corresponding cases (Methods, Supplementary Fig. 4). The included epidemiological connections (Fig. 2b) were established based on i) forward and backward contact tracing interviews (20.5% and 2.2% of edges, respectively); ii) assignment of cases to the same outbreak (11.7% of edges); and iii) the registered address and last name data of cases (65.6% of edges), inducing an epidemiological edge between two case nodes if the cases were registered at the same address and shared (“sameAddress-sameName”), or did not share (“sameAddress-differentName”), the same last name. The overall proportion of sequenced cases was 24.5%; the proportion of sequenced cases on a weekly varying varied between 7.9% and 50.4% (Fig. 2a, Supplementary Table 2) and the sequenced viral genomes were dominated by the Alpha, Delta, and Omicron variants [Rambaut et al. 2020], depending on the time of the year (Supplementary Figure 5).

Forward and backward contact tracing connections, as well as “sameAddress”-type edges, exhibited genetic validation rates (genetic distance *≤* 1) of approximately 80%, whereas “outbreak” edges showed a genetic validation rate of 70% (Fig. 2c, Fig. 2d, Supplementary Figure 6). A set of randomly scrambled contact tracing links (Methods) showed a markedly different distribution of genetic differences (average genetic distance 21; Supplementary Fig. 7e), confirming that the observed genetic distance distribution for the real epidemiological links did not result from incidental genetic similarity between cases sequenced at approximately the same time. The contact tracing connections in the graph mainly represented household-associated transmission (84%); household-associated transmission also accounted for the majority (59%) of registered contacts (independent of whether they led to a transmission or not) and exhibited the highest context-specific transmission rate of infection contexts captured by contact tracing (30%; Supplementary Table 3; Supplementary Figure 8). “Private household” and “recreational context”, the infection contexts with the highest frequency in the forward contact tracing data, did not significantly differ with respect to genetic validation (82% compared to 86%; p = 0.42, Fisher’s Exact Test; Supplementary Figure 9).

### Household-associated transmission dominated explained infections, but classical contact tracing lacked sensitivity for other infection contexts

First, we set out to characterize how well we could explain SARS-CoV-2 transmission in the general population based on the collected data. We first focused on explicitly traced contacts (forward and backward contact tracing, as well as “outbreak”-type connections) within the set of sequenced cases. We clustered sequenced cases by genetic similarity (genetic distance threshold ; n = 1,206 clusters; Supplementary Figure 10) and constructed hypothetical transmission chains (Fig. 3a) from the detected links between cases for each cluster, only allowing for connections between cases that were genetically supported and consistent with respect to symptom onset of the connected cases (i.e. ; Methods) or relying on the given directionality of transmission assessed during backward contact tracing. Using only forward and backward contact tracing data, the constructed chains implied a putative infection source for 12% of cases in clusters (referred to as “Cluster Explanation Rate” or “CER”) and 7% of all sequenced cases; also including “outbreak”-type edges increased the CER to 14% and the proportion of sequenced cases with a putative infection source to 8% (Fig. 3b).

**Figure 3:**
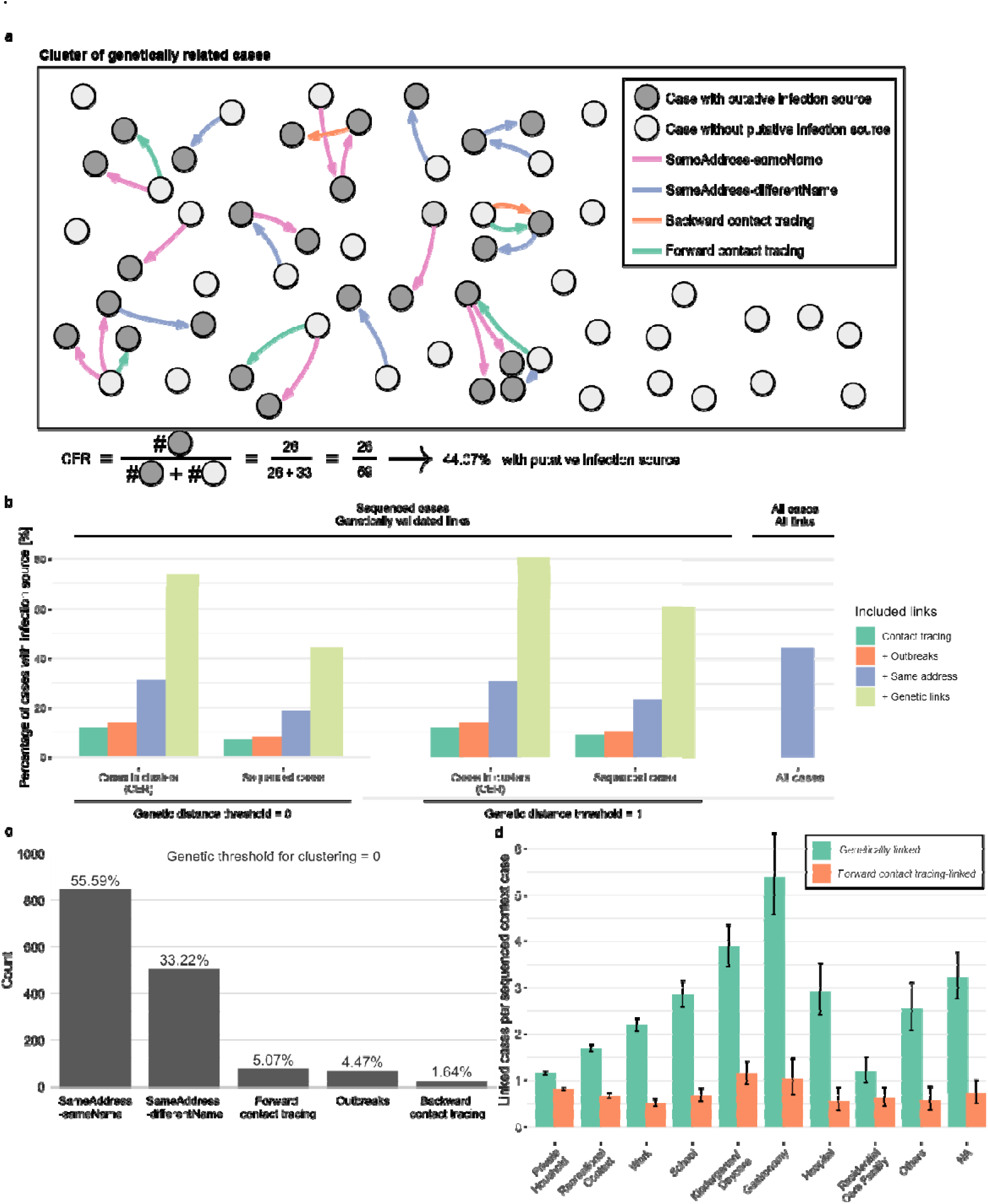
Integrated characterization of SARS-CoV-2 community transmission. **a,** Visualization of a real cluster of genetically related cases (cluster “27” of the, cluster computation; see Supplementary Table 6) and illustration of the CER computation. Nodes correspond to cases and edges to epidemiological connections of different types (after directionalization). Edges rendered with full opacity are part of the hypothetical transmission chain computed for the cluster; faded edges were not selected. We manually added 4 edges to illustrate that some case pairs maintain bidirectional links after directionalization. **b,** Proportion of cases with a putative infection source, stratified by case sequencing and cluster membership status and based on hypothetical transmission chains (see Methods) computed based on genetic clustering similarity thresholds *t_g_* of 0 and 1 and. **c,** Edges contained in the computed hypothetical transmission chains for *t_g_* = *t_j_* =0 and *t_d_* = 0, stratified by edge type. **d,** Average number of genetically linked cases (genetic distance 0 and difference in registration in dates ≤ 14 days) compared to the average number of sequenced cases linked via classical contact tracing for cases with contacts of the given category.

The fact that the genetic validation rate of “sameAddress”-type edges was comparable to that of contact tracing edges indicated that these proxy indicators of joint household membership captured relevant epidemiological relationships. We therefore re-computed hypothetical transmission chains including “sameAddress”-type links, observing an increase in the CER to 31% and in the proportion of sequenced cases with an infection source to 19%. Within the computed chains, 91% of utilized connections were household-associated (Fig. 3c), translating to 28% of sequenced cases in genetic clusters with a household-associated infection source. These results were robust (Supplementary Table 4) to using different thresholds for genetic distance (*t_g_ =* 1), the symptom onset date difference (*t_d_ =* 3), and allowing for one or two intermediate non-sequenced cases (*tj =* 1*/*2); the computed CERs were also relatively similar for different cluster sizes and over the study period (Supplementary Figure 11). *t_d_ =*0 was chosen to enrich the set of considered edges for true-positive transmission events, excluding an expected 75% of edges with incorrect directionality of transmission according to a Bayesian analysis based on backward contact tracing (Supplementary Figure 12), while retaining an expected 89% of edges with correct directionality. *t_d_ =* 3, by contrast, represented a higher expected proportion of edges with correct directionality retained (99% in expectation), while the removal of edges with incorrect directionality was less stringent (46%; Supplementary Figure 12).

To investigate to which extent the computation of hypothetical transmission chains was constrained by the fact that it was carried out within the subset of sequenced cases, we treated the complete Integrated Case Graph of all n = 32,830 cases as one large cluster of genetically identical cases. This led to a substantial increase in the proportion of cases for which a potential infection source could be identified (44.4%; Fig. 3b), with 89% of putative transmissions being household-associated (Supplementary Table 5). However, more than half of cases remained without a plausible infection source, indicating that many transmission-relevant epidemiological connections within the city population were not captured by classical contact tracing (as also suggested by the very large proportion of apparent household-associated transmission), and/or substantial importation of cases.

We used genetic data to gain further insights into missed epidemiological links. Transmissions within the city population would be associated with low genetic distances between cases; imported infections, by contrast, would typically exhibit increased genetic distances. We therefore recomputed hypothetical transmission chains assuming that all genetically linked samples were also epidemiologically connected. For *t_g_ =* 0, the proportion of sequenced cases with a defined infection source increased to 43.9%; for *t_g_* = 1, it increased to 60.5%, consistent with the hypothesis that a substantial number of epidemiological links within the city population were missed by contact tracing (Fig. 3b). To identify the infection contexts that may contribute to these, we labeled each sequenced case with the contexts of all its recorded forward contacts tracing contacts (independent of whether the traced contact was associated with a transmission or not); for each context and the corresponding sequenced cases, we then compared the average number of genetically linked cases (genetic distance = 0 and difference in registration in dates *≤* 14 days) to the average number of sequenced cases linked by classical contact tracing (Fig. 3d). While cases with contacts in the “private household” category had approximately 1.4 as many genetically linked cases as classically traced contacts, cases with contacts in the gastronomy, hospital, school and kindergarten contexts had 3 – 6 times as many genetically linked cases, indicating that these contexts were likely enriched for infection-associated contacts not captured by classical contact tracing.

In conclusion, based on high-confidence sequencing-supported hypothetical transmission chains and transmission chains computed based on only epidemiological connections, we found a potential infection source for 19% of sequenced and 44% of all cases, respectively; at approximately 90%, household-associated transmission dominated both sets of transmission chains, implying a household-associated infection source for up to 40% of all cases. Analysis of genetic data suggested limited sensitivity of classical contact tracing for non-household contacts and that the gastronomy, hospital, school and kindergarten contexts were likely enriched for undetected transmissions.

### Outbreaks were associated with approximately 8% of overall SARS-CoV-2 case load and different impacts on community transmission

Next, we focused on SARS-CoV-2 outbreaks, characterizing transmission both within outbreaks and between outbreaks and the community. We analyzed all n = 162 outbreaks with 919 associated cases registered by Düsseldorf Health Authority during the study period (Supplementary Table 7; employing the definition of an outbreak as three or more cases with an epidemiological link outside of the household setting). The number of cases varied by outbreak category (Fig. 4a, Fig. 4b); 49 school outbreaks accounted for 230 registered cases, followed by 13 care home outbreaks accounting for 161 cases, and 23 outbreaks in kindergartens/daycares accounting for 121 cases; care home outbreaks had the highest number of cases per outbreak (mean = 12.4; Supplementary Table 8). In 48 outbreaks more than one sequenced case was present; of these, 37.5% (n = 18 outbreaks) were classified as not genetically homogeneous because they contained at least one case that was not closely related to the other outbreak cases (genetic distance threshold of ≤ 1; Methods); 12 outbreaks contained more than one SARS-CoV-2 Pango lineage (Supplementary Table 8). Based on a genetic distance threshold of ≤ 1, 15% (n = 31) of the 203 sequenced cases in the outbreaks with more than one sequenced case were found to be likely mis-assigned, i.e. not closely related to the other cases in their corresponding outbreak (Methods).

**Figure 4.**
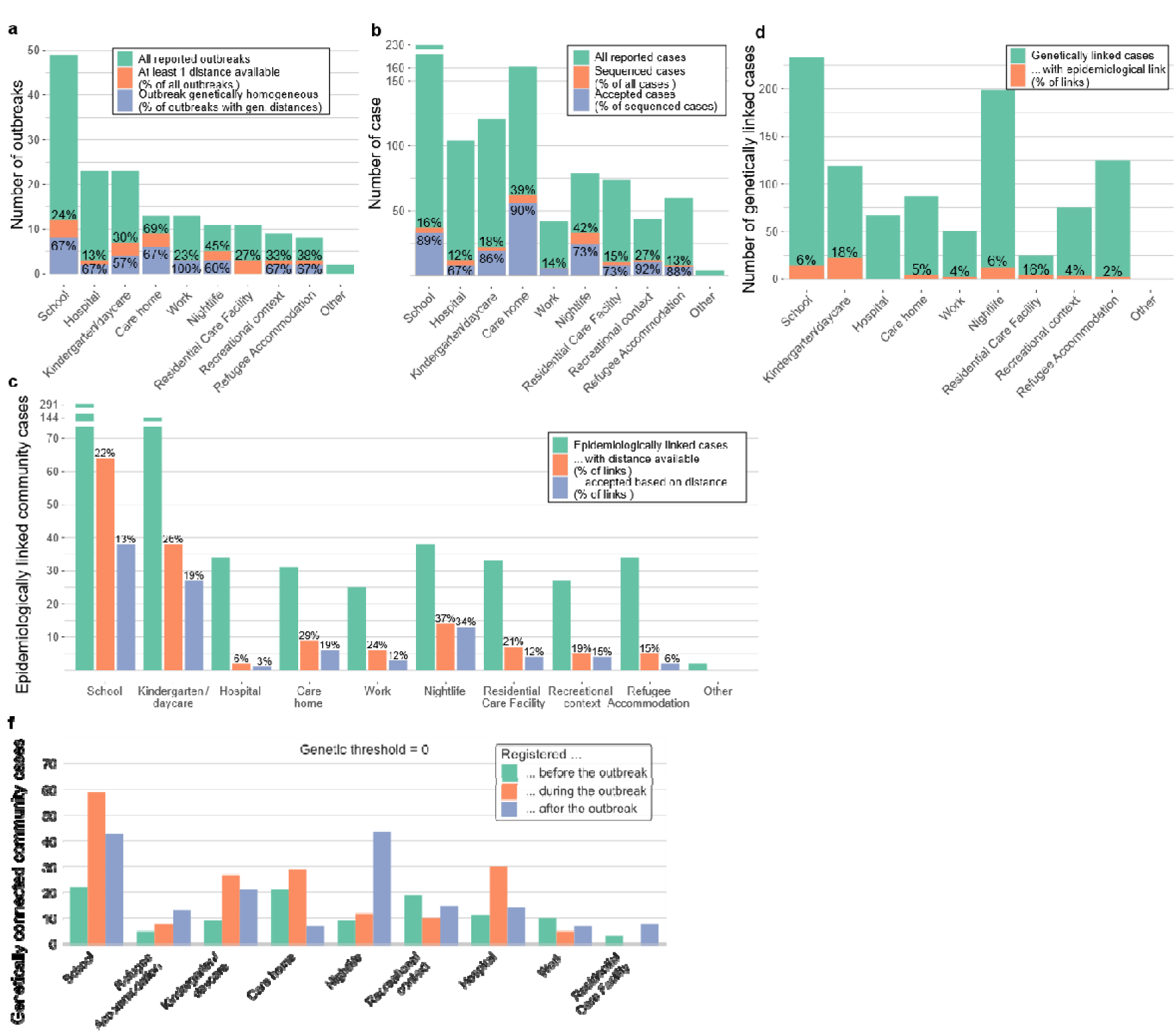
Genomics-based investigation of SARS-CoV-2 outbreaks and their links into the community. **a,** SARS-CoV-2 outbreaks by outbreak type; also shown is the proportion of outbreaks in the respective category with ≥2 sequenced cases and the proportion of outbreaks classified as genetically homogeneous based on a genetic distance threshold of 1 (see Methods). **b,** SARS-CoV-2 cases in outbreaks by outbreak type; also shown is the proportion of sequenced cases in the respective outbreak category and the proportion of sequenced cases in the corresponding outbreak category classified as “accepted” (i.e., part of the largest genetic cluster in the outbreak, based on a clustering threshold of 1; Methods). **c,** Cases epidemiologically linked to outbreaks, by outbreak type; also shown is the proportion of epidemiologically linked cases in the corresponding outbreak category for which a genetic distance to their associated outbreak could be computed, and the proportion of such cases for which the computed distance was 1 (“accepted”). The genetic distance between a case epidemiologically linked to an outbreak and the outbreak was defined as the minimum genetic distance between the linked case and any case of the outbreak; computation of a such a distance was possible if the outbreak contained at least one sequenced case and if the linked case itself was sequenced. **d,** Cases genetically linked to outbreaks (genetic distance 0, case registration date difference ≤ 14 days), by outbreak type; also shown is the proportion of genetically linked cases that also exhibited an epidemiological link to their corresponding outbreak. **e,** Cases genetically linked to outbreaks and stratified by outbreak type and relative case registration date; 8 outbreaks that were associated with the Delta and Omicron variants and that overlapped, with their surrounding 2-week windows for the inclusion of linked variants, with the expansion phases of these variants were not included.

As a first-order measure of the role of outbreaks in the wider transmission network of SARS-CoV-2, we determined the number of community cases connected to outbreaks (represented by any one of their cases) via links of type contact tracing and “sameAddress” in the Integrated Case Graph. 659 community cases were found to be epidemiologically linked to at least one outbreak (Fig. 4c), accounting, when combined with outbreak cases, for approximately 5% of cases in Düsseldorf over the considered period. Of the identified outbreak-linked cases, 22% (n = 150 / 659) could be evaluated genetically based on the availability of a genetic distance between the outbreak-linked case and any one outbreak case; of these, 65% (n = 98 / 150) were confirmed based on a genetic distance of *≤* 1. Schools and kindergartens combined accounted for approximately two thirds of overall and sequencing-confirmed linked cases (Supplementary Table 8); these two outbreak types also had the highest number of linked community cases per outbreak case (5.9 and 6.2, summed over all school and kindergarten outbreaks; Supplementary Table 8, Supplementary Figure 13 L). By contrast, care homes exhibited the lowest proportion of linked community cases relative to registered outbreak cases (average = 0.2; Supplementary Table 8) and were therefore, despite high overall case count, associated with a relatively low absolute number of linked community cases. “sameAddress”-type links accounted for 1.7 times as many overall and approximately twice as many sequencing-confirmed linked community cases as contact tracing-implied links, respectively (Supplementary Table 8).

As an orthogonal measure of the connectedness between outbreaks and the community that was not affected by potential biases of classical contact tracing, we also determined the number of community cases that were, within a two-week time window, genetically linked to an outbreak (Fig 4d; Methods). In total, 1,000 community cases were found to be linked to an outbreak. Of these, 94% did not exhibit an epidemiological link to the outbreak they were genetically linked to, increasing the total proportion of outbreak-associated cases (outbreak cases plus genetic and epidemiological links) to 8%. School and nightlife outbreaks were associated with the largest number of genetically linked community cases (n = 243 and n = 200, respectively; Supplementary Table 5); nightlife outbreaks also had the highest number of genetically linked community cases when normalized by outbreak size (Supplementary Table 8, Supplementary Figure 13). To assess the potential role of outbreaks as drivers of community transmission, we stratified the genetically linked community cases by whether they were registered before, during or after their associated outbreaks (Fig. 4e); to avoid confounding by variant-associated growth effects, we removed 8 outbreaks that were associated with the Delta and Omicron variants and that overlapped, with their surrounding 2-week windows, with the expansion phases of these variants (before week 29 for Delta and all 4 Omicron outbreaks in weeks 48 to 51; see Supplementary Figure 5). Nightlife outbreaks were associated with a four-fold increase in the number of associated cases in the post-compared to the pre-outbreak period; schools and kindergartens, with an approximately two-fold increase. Care homes, by contrast, were associated with a comparatively low number of genetically linked post-outbreak cases.

In summary, outbreaks were associated with 8% of total cases. Genetic data confirmed the accuracy of the outbreak documentation process of Düsseldorf Health Authority implemented according to best-practice guidelines, with only 15% of cases found to be incorrectly assigned to outbreaks. Based on classical epidemiological and genetics-based measures, school, kindergarten, and nightlife outbreaks were strongly linked to the community and associated with a post-outbreak increase in genetically linked cases, indicating a likely role in driving community transmission. The high degree of community connectedness of nightlife outbreaks was only apparent in the genetic analysis, indicating that classical outbreak control and investigation approaches were less effective for these.

### In-depth manual analysis of school outbreaks identified additional outbreak-linked cases and links between outbreaks

To assess whether genetic data could contribute to the discovery of epidemiological connections not captured by routine contact tracing, and to investigate the potentially important role of schools for SARS-CoV-2 transmission, we carried out an in-depth manual investigation of SARS-CoV-2 transmission within the n = 49 registered school outbreaks and their links to the community (Supplementary Table 7; Supplementary Table 8). We (i) manually reviewed the complete Düsseldorf Health Authority database records of all relevant cases and (ii) searched for additional outbreak-linked community cases based on genetic data (Methods; Supplementary Figure 14). According to routine contact tracing, the investigated outbreaks comprised 226 cases, of which 51 were sequenced and 4, based on genetic distances, falsely assigned (i.e. not part of the outbreak they were assigned to by routine contact tracing); as well as epidemiological links to 291 community samples, of which 38 were genetically validated (Fig. 5a).

**Figure 5.**
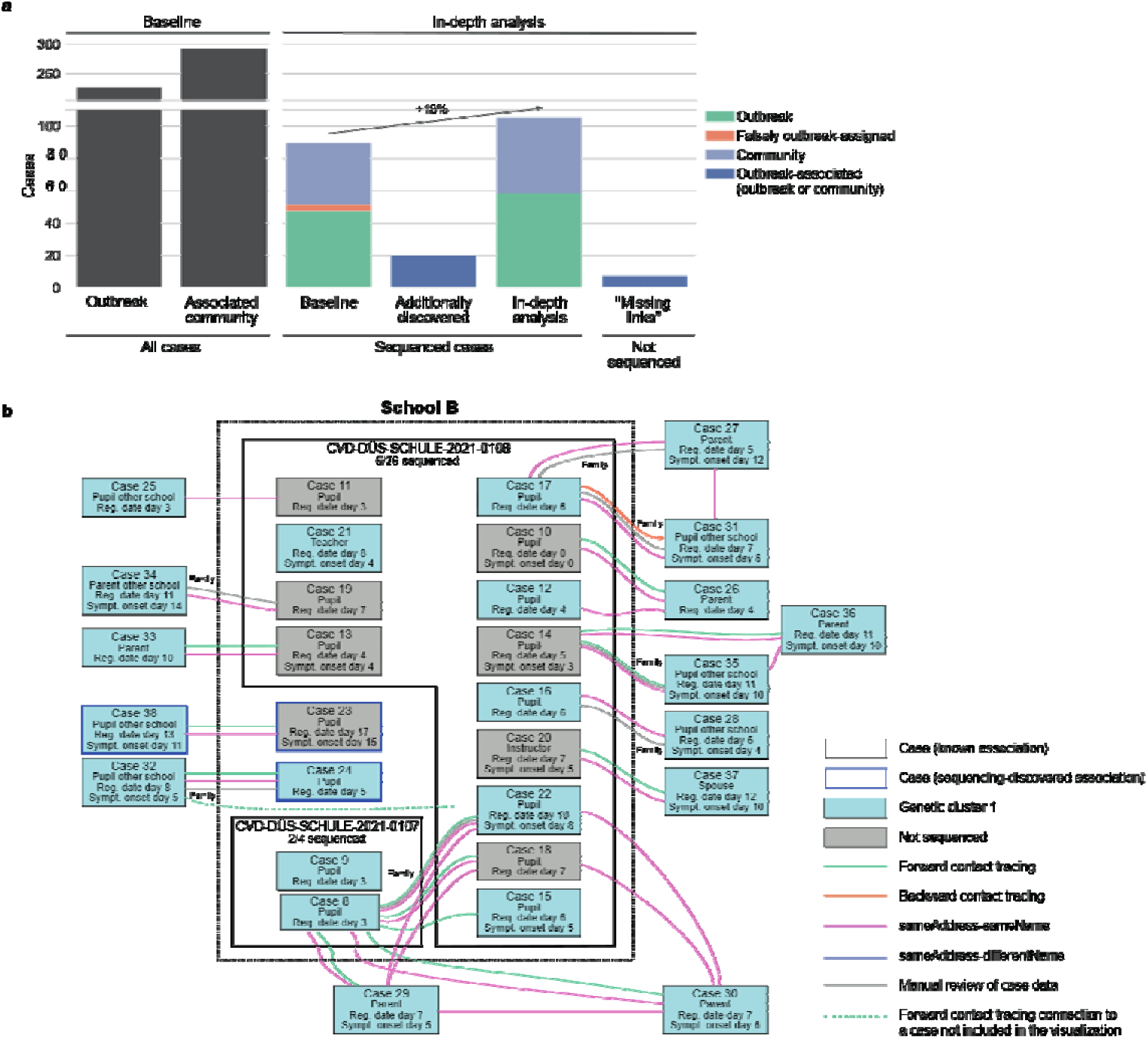
In-depth analysis of school outbreaks. **a,** School outbreak-associated cases before and after in-depth manual analysis. Black bars show the total number of school outbreak-associated cases before manual analysis and independent of sequencing status; colored bars show the results of in-depth analysis for the subset of sequenced outbreak-associated cases, as well as the number of non-sequenced cases that were classified as outbreak-linked because they represented the required “missing epidemiological link” between an outbreak and a genetically linked third case. **b,** Visualization of two school outbreaks (0107 and 0108), registered at the same school, and of the epidemiological links between the associated cases. All sequenced cases (boxes with blue background) were part of the same genetic cluster (see Supplementary Note 1 for a full genetic distance matrix); for outbreak 0108, only a subset of cases is shown. Cases 23, 24, and 38 were classified as outbreak-associated based on the analysis of genetic data; Case 23 is an example of a “missing epidemiological link” that was classified as outbreak-associated because it represented the only detected epidemiological link that could account for the genetic link between Case 38 and the two shown outbreaks.

First, we carried out a search for additional outbreak-linked cases. In some instances, free-text database entry fields were used to record the results of contact tracing interviews; we evaluated whether contacts recorded in these (i.e., in a non-structured manner and thus not amenable to automated extraction) increased the number of outbreak-linked community cases, but found only a single instance of a non-structured contact tracing record implying a link between a community case and a school outbreak that was not already represented in the Integrated Case Graph (via an edge of any type between the contact and any case of the associated outbreak). We turned to the analysis of genetic data for the identification of additional outbreak-linked cases and found 20 sequenced cases (Fig. 5a) that were not linked to a school outbreak in the Integrated Case Graph or mentioned in free-text contact tracing records, but for which a genetic link to a school outbreak (genetic distance threshold ≤ 1) could be epidemiologically confirmed through manual review of case database records, typically based on information found in other free-text entry fields. For example, in the analysis of outbreaks 0107 and 0108 (Fig. 5b), Case 24 was classified as outbreak-linked because it genetically clustered with the outbreaks and because the case was found to be a pupil at the school at which the outbreaks occurred, confirming an epidemiological link. In addition, we found 7 non-sequenced cases that were classified as outbreak-linked because they represented the “missing epidemiological link” between an outbreak and a genetically linked third case (Fig. 5a, Methods); such as Case 23, which represented the only detected epidemiological link that could account for the observed genetic link between Case 38 and outbreaks 0107 and 0108 (Fig. 5b).

Second, we evaluated whether the assignment of cases to outbreaks was complete. We focused on n = 58 sequenced cases that were not registered as school outbreak cases according to the original outbreak documentation carried out by Düsseldorf Health Authority, but genetically and epidemiologically linked to a school outbreak (including the 20 genetically identified outbreak-linked cases described above). Of the 58 investigated cases, 11 met the criteria for being classified as outbreak cases – for example Cases 23 and 24, as pupils at the school at which outbreaks 0107 and 0108 occurred (Fig. 5b). Correct registration of the 11 cases would have increased the number of sequenced outbreak cases by 22% (compared to a baseline of 51 sequenced outbreak cases, of which at least 4 were misclassified according to genetic data; Fig. 5a and Supplementary Table 8).

Third, we investigated whether there were connections between separately registered school outbreaks and found that 8 school outbreaks were epidemiologically and genetically connected to at least one other school outbreak (Supplementary Table 9; Supplementary Note 1). Examples include the genetic and epidemiological links between outbreaks 0107 and 0108 (Fig. 5b), which were two separately registered multi-class outbreaks at the same school, as well as the detected links between the separately registered outbreaks 0212, 0231, and 0231 at the same school. Due to the detected epidemiological links, these, like the other outbreaks found to be linked in our analysis, should have been registered as single outbreaks.

In summary, the in-depth analysis of school outbreaks demonstrated that sequencing could enable the discovery of novel epidemiological connections not captured by the extracted Düsseldorf Health Authority routine data. The sequencing-based analysis showed that that the number of cases associated with school outbreaks was higher than initially estimated (18% increase within the set of sequenced cases, Fig. 5b) and that independently registered outbreaks were in fact epidemiologically connected.

### High sequencing coverage was required for high-resolution characterization of transmission

Last, we asked which case sequencing coverage was required for the effective implementation genomics-enhanced contact tracing (Figure 6, Supplementary Figure 15, Supplementary Figure 16). Downsampling the set of available SARS-CoV-2 sequences when actual pairwise genetic distances were required, or randomly increasing the proportion of sequenced cases otherwise, we analyzed the influence of the sequencing rate on four key areas that benefit from the availability of sequencing data: (i) genetic validation of epidemiological connections (Fig. 6a); (ii) outbreak investigations with respect to genetic homogeneity (Fig. 6b) and genetic links to the community (Fig. 6c); (iii) infection source analysis, based on high-confidence sequencing-supported hypothetical transmission chains (Fig. 6d); and (iv) context analyses, with respect to the identification of infection contexts underrepresented in classical contact tracing (Fig. 6e) and of outbreak types associated with an increase in post-outbreak genetically linked cases (Fig. 6f). Across analyses, higher case sequencing rates consistently led to improved characterization performance. For the context-specific analyses specifically, we found that a sequencing rate of 15% was sufficient to recover the three infection contexts (schools, kindergartens, gastronomy; Fig. 6e) likely enriched for undetected transmissions and two (nightlife, schools; Fig. 6f) of three outbreak types associated with a post-outbreak increase in linked cases; at a sequencing rate of 10%, by contrast, context-specific confidence intervals were still largely overlapping and increases in post-outbreak linked cases less pronounced. A sequencing rate of 15% would thus have been sufficient to recover the majority of results about SARS-CoV-2 transmission dynamics.

**Figure 6:**
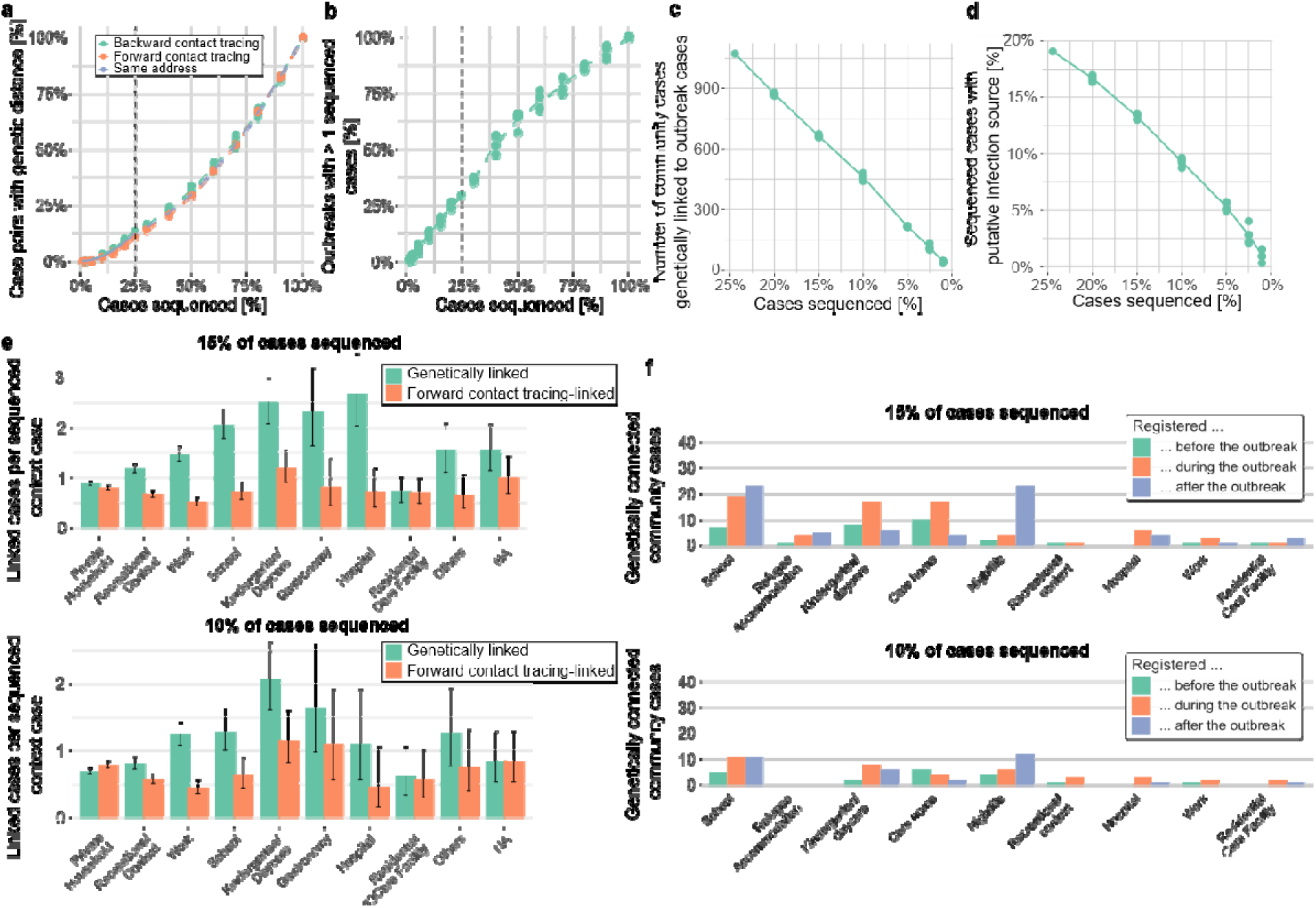
Influence of the proportion of sequenced cases on the ability to characterize SARS-CoV-2 transmission. **a,** Proportion of edges in the Integrated Case Graph that could be genetically validated at different case sequencing rates (5 replicates), shown for different edge types. **b,** Proportion of registered outbreaks that could be genetically validated (based on > 1 sequenced outbreak case, 5 replicates). **c,** Number of community cases genetically linked to outbreaks (5 replicates). **d,** Proportion of sequenced cases with a putative infection source, based on hypothetical transmission chains re-computed for each replicate (5 replicates). **e,** Average numbers of genetically linked and sequenced cases linked via classical contact for cases with contacts of the given category at different case sequencing rates; see Fig. 3d for results based on the complete data. **f,** Cases genetically linked to outbreaks, stratified by outbreak type and relative case registration date, at different case sequencing rates; see Fig 4e for results based on the complete data.

## Discussion

In 2021, we carried out one of the highest-intensity SARS-CoV-2 integrated genomic surveillance programmes in continental Europe and implemented genomics-enhanced contact tracing (Fig. 1, Fig. 2). The resulting data set of more than 30,000 cases, almost 50,000 contact tracing records, and 8,000 viral genomes represents a rich resource on the structure of SARS-CoV-2 community transmission in an internationally connected city and against the backdrop of a public health mitigation strategy. We used the collected data to characterize SARS-CoV-2 transmission in the general population and in and around outbreaks under these conditions.

We characterized SARS-CoV-2 transmission using two complementary approaches. First, we used the combined epidemiological and genetic data in the Integrated Case Graph to reconstruct hypothetical transmission chains of SARS-CoV-2. We found a potential infection source for 19% of sequenced and 44% of all cases, respectively, with household-associated transmission accounting for up to 40% of all cases. Many cases that remained unexplained based on the available epidemiological links had close genetic links to other cases, suggesting that many relevant transmissions were not captured by classical contact tracing. Further analysis of the genetic data then showed that the gastronomy, hospital, school and kindergarten contexts were likely enriched for such undetected transmissions. Second, based on a large set of n = 162 outbreaks, we characterized SARS-CoV-2 transmission in and around outbreaks. Schools, kindergartens and nightlife outbreaks exhibited a high, and care homes a relatively low, degree of linkage to the community. In a targeted analysis of school outbreaks, we found previously unrecognized connections between outbreaks and showed that routine classical contact tracing had underestimated the number of outbreak-associated cases (19% increase within the set of sequenced cases). Time-stratified analysis of genetically connected community cases suggested a particular effect of nightlife outbreaks on driving post-outbreak community transmission. Both arms of our analysis thus consistently identified schools, kindergartens and gastronomy and nightlife settings as likely playing an important role in transmission dynamics.

Sequencing data was required for or supported key aspects of our analysis. Sequencing enabled the validation of epidemiological relationships and was key to confirming the connectedness of cases in the investigated outbreaks. For hypothetical transmission chains, genetic validation supported the inclusion of “sameAddress”-type edges, which accounted for the majority of utilized links and connected a substantial number of cases not otherwise connected (Supplementary Figure 17), alongside explicitly traced contacts. The identification of infection contexts that were likely enriched for transmissions not detected by classical contact tracing depended on the availability of sequencing data. For outbreaks, sequencing provided an additional measure of the connectedness between outbreaks and the community and enabled the detection of novel epidemiological connections; for school outbreaks, sequencing was required to obtain a more accurate assessment of the total number of school outbreak-connected cases. For nightlife outbreaks, their high degree of community connectedness was only visible in the genetic, but not in the classical contact tracing, data.

Similar proportions of household-associated transmission were observed based on digital contact tracing [Ferretti et al. 2024], during Iceland’s first SARS-CoV-2 wave once the impact of travel-imported infections had been replaced by endogenous transmission [Gudbjartsson et al. 2020], and in other epidemiological studies [Manica et al. 2023]. Compared to surveys of contact patterns [Mossong et al. 2008], including surveys of contact patterns carried out in Germany during the pandemic [Phuong et al. 2025], and digital contact tracing [Ferretti et al. 2024], household contacts were likely overrepresented by at least a factor of 2 in our dataset, consistent with the interpretation that gaps in contact tracing accounted for a substantial fraction of unexplained cases. Complementing these studies, however, we could identify specific infection contexts that were likely enriched for undetected transmission events. Outbreaks [Leclerc et al. 2020, Wong et al. 2020] have been identified as important contributors to SARS-CoV-2 transmission, but most analyses described individual outbreaks [Allen et al. 2022, Nicolai et al. 2025]; relied on epidemiological modelling [Rodiah et al. 2023]; or did not have access to genomic data to validate the association of cases to individual outbreaks [Manica et al. 2023, Garcia-Bernardo et al. 2024]. The results reported here, by contrast, provide a comprehensive characterization of SARS-CoV-2 outbreaks based on a large data set of more than 150 institutional outbreaks registered a single city and employing a combination of epidemiological and genetic approaches.

Our study also extends the existing literature on genomics-enhanced contact tracing. Most previous successful implementations [Gudbjartsson et al. 2020, Seemann et al. 2020, Lane et al. 2021, Hjorleifsson et al. 2022, Hall et al. 2023] were carried out in the context of island nations with the ability to effectively control viral importations, relatively low incidences, and highly effective contact tracing carried out to support a public health viral elimination strategy. By contrast, our study was carried out in the epidemiologically more open context of a single city in a large metropolitan area, under conditions of relatively higher incidences, and against the backdrop of a public health mitigation strategy with the aim of using contact tracing to reduce, but not eliminate, infections. Our results demonstrate that genomics-enhanced contact tracing under these conditions, which were broadly representative of the current or expected conditions in many non-island localities during a pandemic or major outbreak, enabled insights into transmission dynamics which could not be obtained from classical contact tracing alone.

The availability of large-scale sequencing data could also contribute to the pandemic public health response on multiple levels. First, sequencing showed high genetic validation rates for both contact tracing and outbreak registration, providing a measure of internal quality control and implicitly validating the contact tracing guidelines of Germany’s Robert Koch Institute. Second, the improved characterization of outbreaks could inform the implementation of improved infection prevention measures. For example, in the aftermath of the sequencing-informed investigation of a large nightlife outbreak [Houwaart et al. 2022], Düsseldorf Health Authority implemented a more stringent regime of monitoring the compliance of the affected bars with mandatory infection prevention measures. Third, sequencing could help prioritize high-risk transmission settings for in-depth tracing and investigation activities. Based on the results presented here, improved surveillance in e.g. nightlife and gastronomy contexts may be particularly important. Last, sequencing could contribute to an improved calibration of policy decisions via evidence generation, e.g. by informing decisions on school closures with more accurate information on contribution of schools to overall community transmission. Of note, in our study, we successfully implemented large-scale genomics-enhanced contact tracing in a retrospective manner based on moderately low turnaround times (generally < 1 week; Supplementary Figure 3); additional applications could be enabled by even lower turnaround times, which may include e.g. the detection of active infection clusters in real time.

With respect to future implementations of genomics-enhanced contact tracing, our results imply that the effective characterization of infection contexts underrepresented in classical contact tracing and of outbreak types associated with an increase in post-outbreak genetically linked cases require a sequencing rate of at least 15%; these applications are of high public health relevance even if the ability to reconstruct individual transmission chains remains limited. Even higher sequencing rates may be required for a fine-scale characterization of transmission at the level of individual transmission chains in and around outbreak contexts of particular public interest, such as schools; the role of schools during the pandemic remains controversial [Lordan et al. 2021, Amodio et al. 2022, von Bismarck-Osten et al. 2022, Neil-Sztramko et al. 2024], and, at a case sequencing coverage of 24.5%, we almost certainly missed additional cases connected to school outbreaks. With respect to the collection of epidemiological information, our results suggest that information on joint household membership obtained from explicit case interviews may largely overlap with information that could be obtained from case registration data; increased reliance on routinely available data for capturing household contacts may thus free up resources for intensified surveillance and investigation in other epidemiological contexts. The feasibility of investigating a higher proportion of otherwise unexplained sequencing-indicated links between cases may be further increased by the use of large language models for gathering epidemiological information, for example through automating parts of the case interview process.

In conclusion, we demonstrated the successful implementation of genomics-enhanced contact tracing for SARS-CoV-2 under the conditions of high-incidence community transmission in a non-isolated geographical entity. We showed that the integration of sequencing yielded information relevant outbreak control, infection prevention and public health policy that could not be obtained from classical contact tracing alone, and that these applications require a sequencing rate of at least 15%. Our study thus informs future implementations and applications of genomics-enhanced contact tracing.

Limitations of this study: A link between two cases in the constructed hypothetical transmission chains does not imply an actual transmission event; in particular when computed independent of genetic data, the proportion of cases with a potential infection source should be interpreted as an upper bound on the ability to explain individual cases with the available epidemiological data. Edges of type “sameAddress” are imperfect indicators of joint household membership and edges of type “sameAddress-differentName”, in particular, may also sometimes capture transmission in e.g. residential facilities or large building complexes (Supplementary Figure 18, Supplementary Figure 17). Non-Düsseldorf contacts could not be analyzed in our study; however, non-Düsseldorf contacts only accounted for 19% of overall contacts and their improved analysis would, given the limited role of forward contact tracing-induced edges in the computed hypothetical transmission chains, likely not have led to a significant increase in the proportion of cases with a potential infection source or different conclusions with respect to infection contexts. Household contacts were substantially overrepresented in the collected forward and backward contact tracing data and confounding by other epidemiological factors could not be ruled out in the contact tracing informed-analyses of genetic data. The case sequencing coverage in our study was only 24.5%; increasing the proportion of sequenced cases while keeping the available contact tracing information constant might have led to an increase in CERs and in the proportion of cases with a potential infection source, but not beyond the 44.4% reported above for the complete Integrated Case Graph. Our study was based on data collected over a period of 11 months in 2021 and our results may not generalize to viral variants of SARS-CoV-2 not covered by the study period, which may exhibit different transmission properties. Our analysis did not incorporate a potential effect of vaccination on SARS-CoV-2 transmission. Finally, the generalizability of the results to other respiratory pathogens beyond SARS-CoV-2 is an important direction for future research.

## Methods

### Implementation of genomics-enhanced contact tracing in Düsseldorf

Genomics-enhanced contact tracing was implemented using the Integrated Genomic Surveillance System Düsseldorf (IGSD), which has been fully described elsewhere [Houwaart et al. 2022, Walker et al. 2022]. Briefly, the high-level aims of the IGSD were to monitor viral evolution in the Düsseldorf area and to investigate whether a combination of high-density genomic surveillance with classical epidemiological approaches could enable an improved understanding of SARS-CoV-2 transmission in the population at large. The IGSD was implemented as a collaboration between Heinrich Heine University Düsseldorf (HHU), Zotz | Klimas (ZK) and Medizinische Laboratorien Düsseldorf (MLD), two large commercial diagnostic laboratories from Düsseldorf, and Düsseldorf Health Authority. Viral samples provided by ZK and MLD, as well as viral samples collected from patients treated at Düsseldorf University Hospital, were rapidly Nanopore-sequenced using HHU’s SARS-CoV-2 sequencing platform; SARS-CoV-2 sequences generated by MLD were also integrated.

### Case registration and contact tracing carried out by Düsseldorf Health Authority

SARS-CoV-2 case data collection and contact tracing, using the SurvNet application [Faensen et al. 2006, Krause et al. 2007] for data storage and management, were carried out by Düsseldorf Health Authority under the mandate of the Law on the Prevention of Infection (Infektionsschutzgesetz (IfSG) based on the classification of SARS-CoV-2 as a notifiable pathogen in Germany since 2020 and implementing on guidelines by Germany’s Robert Koch Institute. This involved (i) the reporting to Düsseldorf Health Authority of all Düsseldorf residents tested positive for SARS-CoV-2 by diagnostic laboratories; (ii) the registration of the personal data of all registered cases by Düsseldorf Health Authority; (iii) contact tracing interviews with all Düsseldorf cases conducted by Düsseldorf Health Authority, the participation in which was mandatory; (iv) the reporting of all suspected outbreaks (see “Retrospective investigation of outbreaks” for the applied definition of an outbreak) and their involved cases to Düsseldorf Health Authority. Contact tracing was based on the official contact tracing guidelines in place at the respective time, as published by the Robert-Koch-Institute. It always involved a forward contact tracing component and also typically a backward contact tracing component. Forward contact tracing was carried out for quarantine and isolation purposes, aiming at the identification of all individuals that a case had been in contact with around a time window of 2 days before symptoms onset (if symptomatic) or 2 days before the case’s positive diagnostic test (if asymptomatic), irrespective of the potential directionality of transmission. Personal data of unvaccinated contacts, and of vaccinated contacts registered up until 20 April 2021, were always entered into SurvNet in a structured manner (and thus represented as edges in the Integrated Case Graph, see below); on 21 April 2021, mandatory quarantine requirements were lifted for fully vaccinated contacts, and whether and in which form (structured or using free-text entry fields) personal data of fully vaccinated contacts were entered into SurvNet after that date was decided by the individual carrying out the contact tracing interview. Of note, the number of registered contacts per case remained relatively similar over the course of 2021, except for November and December 2021 (Supplementary Fig. 5). During backward contact tracing, the infected individual was asked whether they could identify whom they thought they had likely acquired their infection from. Backward contact tracing data were usually entered into SurvNet in a structured manner. During contact tracing interviews, responses by the interviewed individuals were assessed for plausibility and likelihood that the contact led to a transmission; contacts that were deemed unlikely to have led to a transmission (e.g., between two masked individuals in a well-ventilated area) were not recorded. Contact tracing also involved an assessment of the context in which the detected contacts had taken place in; for structured forward contact tracing records, context information was stored in a structured manner as part of the contact tracing records using a controlled vocabulary (Supplementary Table 11); otherwise, context information was sometimes stored in free-text entry fields.

### Genetic distance matrix

A matrix of genetic distances between all analyzed sequences was calculated as previously defined [Houwaart et al. 2022]. Briefly, based on a multiple sequence alignment (MSA) of the included sequences computed using MAFFT [Katoh and Standley 2013] following GISAID [Shu and McCauley 2017] instructions, the distance *d(x, y)* between samples x and y was defined as the count of differences between the entries of *x* and *y* in the MSA, (i) ignoring leading or trailing “gap” characters, (ii) accounting for International Union of Pure and Applied Chemistry (IUPAC) ambiguity codes, (iii) treating subsequent non-matching gaps columns as a single difference, (iv) ignoring deletions in the MSA entry of x aligned to ‘N’ characters in y and vice versa, and (v) not counting any differences found between the beginning of the MSA and the 20th A/C/G/T character of either sequence and the end of the MSA and the 20 last A/C/G/T characters of either sequence.

### Integrating genomic and viral genome sequencing data into a case graph

Integration of genomic, case, and contact tracing data was carried out using a custom Microsoft Access database application deployed within the protected IT infrastructure of Düsseldorf Health Authority. All personally identifiable information remained at Düsseldorf Health Authority. See Supplementary Fig. 2 for a schematic of the data integration process.

As a first step, case and contact tracing data for SARS-CoV-2 cases from Düsseldorf were exported from the SurvNet application and imported into the custom Access database. Routinely collected data included (a) case-identifying information, such as name, date of birth and registered address; (b) forward contact tracing data; forward contact tracing data comprised the SurvNet database ID of the corresponding case (i.e. specifying the individual with whom the contact tracing interview was carried out), as well as the first and last names, the date of birth, and the residential status (Düsseldorf resident or not) of the registered contacts; the exported forward contact tracing data records, however, did not comprise information on the subsequent infection or case registration status of the contacts (see next section); (c) backward contact tracing data (“InfectedBy” field), only available for a small proportion of cases; backward contact tracing records comprised the SurvNet case ID of the individual with whom the contact tracing interview was carried out (the infected individual) as well as the SurvNet case ID of the individual who was registered as the putative source of the infection (i.e. both individuals were necessarily registered as cases); (d) outbreak data, specifying, for each registered outbreak, details on the specific institutional context as well as a list of case IDs assigned to the outbreak.

As a second step, the following post-processing steps were carried out. (i) The total number of registered forward contact tracing records on a per-case basis was tabulated and attached to each case record, separately for all registered contacts and registered contacts from Düsseldorf. (ii) For each registered forward contact tracing contact from Düsseldorf, the case database was queried for cases with matching first and last names and a matching date of birth; for each match, a case-resolved forward contact tracing record, comprising the case IDs of both the interviewed individual and of the registered contact, as well as the database ID(s) of the contact tracing record(s), was created and stored in the database (with additional filters on the maximum difference between the registration dates of any two cases so-connected applied later, see below; these date-based filtering steps also resolved instances in which a contact tracing record matched more than one case record, e.g. because the same individual tested SARS-CoV-2-positive more than once); (iii) For each case (“query case”), other cases with case registration dates within a 30-day window around the registration date of the query case (≤ 30 days absolute date difference) and (a) with the same registered address as the query case or (b) sharing the same registered address and the same last name as the query case were identified and stored as “SameAddress” and “SameAddressAndLastname” records, comprising the case IDs of the linked cases, in the database. (iv) For each outbreak, the set of all pairs of cases assigned to the outbreak was determined and each pair was stored as a record of type “SameOutbreak”, comprising the case IDs of the two member cases.

As a third step, SARS-CoV-2 genome sequencing records were matched with cases, based on the following procedure. (i) All sequenced samples, and therefore all sequenced SARS-CoV-2 genomes, were labelled with a sample collection date and a PCR test ID by the diagnostic laboratory providing the sample. (ii) Based on these data, we queried data sheets provided by the diagnostic laboratories for the first and last name and the date of birth of the individuals that had provided the sequenced sample. (iii) We matched these data against the SurvNet case registration records; if exactly one match covering all 3 criteria was identified, the viral genome sequence was attached to the SurvNet case record; otherwise, manual curation was carried out to account e.g. for last name spelling mistakes in one of the two data sources. (iv) For all viral genome sequences attached to case records, the difference between case registration date and sample collection date was calculated, and all n = 36 viral genome sequences with a difference < -21 or > 7 were removed (Supplementary Figure 19)

As a fourth step, for all n = 533 cases *c* with more than one assigned viral genome sequencing record, a unique record was selected based on the following procedure. (i) If all sequences attached to a case had genetic distance *≤* 1 between each other, we selected the sequence with the smallest number of undefined nucleotides (“N” characters). (ii) Otherwise, define the set *C_contacts-sequenced_*as the set of other cases that were linked to case *c* via contact tracing and that had attached viral genome sequencing data (there we no instances of cases in *C_contacts-sequenced_* that had more than one attached viral genome sequencing record). (iii) For each viral genome sequencing record *s* attached to *c* (more than one by definition), define *compatible(c, s)* as the set of cases in *C_contacts-sequenced_* that had an associated viral genome sequencing record with genetic distance *≤* 1 to *s* (see Supplementary Figure 19). (iv) Determine *arg max compatible(c, s)* and attach the corresponding viral genome sequencing record *s* to *c*.

Finally, the integrated dataset was exported as an Integrated Case Graph in which (a) each registered case from Düsseldorf, together with non-identifiable metadata such as the number of total forward contact tracing contacts and forward contact tracing contacts from Düsseldorf, was represented as a node; (b) connections between pairs of cases, i.e. forward and backward contact tracing relationships, “sameAddress”-type information, as well assignment of two cases to the same outbreak, were represented as undirected edges; and (c) SARS-CoV-2 genome sequencing records of cases were attached to nodes in the case graph. Edges of type “sameAddress-sameName” were created based on records of type “SameAddressAndLastname”; edges of type “sameAddress-differentName” were created for all sample pairs that appeared in “SameAddress”-type records, but not in “SameAddressAndLastname”-type records. All edges *(c*_1_*, c*_2_) of type “backward contact tracing” between cases *c*_1_ and *c*_2_ were annotated with the information on whether *c*_1_ specified *c*_2_, and *c*_2_, *c*_1_, as their assumed infection sources (note that these two cases were not mutually exclusive). All edges between pairs of cases with an absolute difference in case registration dates of more than 14 days were removed [Ryu et al. 2022]. A distance matrix for all SARS-CoV-2 genomes attached to the nodes of the graph was exported with the graph.

### Infection context analysis

To characterize context-specific transmission probabilities, we carried out a separate database export of all forward contact tracing records stored in SurvNet, comprising, for all exported records, the SurvNet database ID of the index case (i.e. specifying the individual with whom the contact tracing interview was carried out); the residential status (Düsseldorf resident or not) of the contact; the database ID of the contact tracing record; and the contact context (determined by Düsseldorf Health Authority staff during contact tracing; see Supplementary Table 11). We filtered the exported records for index cases with a case registration date *≤* 17 December 2021 and represented in the Integrated Case Graph and removed all records with contacts not registered at a Düsseldorf address. A recorded contact was classified as associated with a potential transmission event if and only if (i) a matching case-resolved forward contact tracing record (see previous section; matching was carried out based on the case ID of the index case and the database ID of the contact tracing record) was identified and (ii) the identified case-resolved forward contact tracing record was represented by an edge of type “forward contact tracing” in the Integrated Case Graph after application of all filtering steps; in all instances satisfying the two conditions, the individual that the contact was registered with had also tested SARS-CoV-2 positive in a period of 14 days around the registration date of the index case, compatible with a direct transmission event. Context-specific transmission probabilities were then calculated by dividing the number of contacts associated with a potential transmission event by the total number of registered contacts for each context. Context annotation for edges of type “forward contact tracing” was carried out based on the same database export and matching process, but omitting the date filtering step (i.e., the annotation process was also applied to edges associated with index cases registered after 17 December).

For backward contact tracing records, export of infection context information from SurvNet in a structured manner was not possible. We therefore employed a manual classification approach, based on review of the data stored in SurvNet (involving all structured case data as well as free-text notes taken during routine contact tracing interview, if available). Each backward contact tracing contact was, based on the definitions given below, assigned to one of the following categories: (i) “Private household”: Contact between two individuals in the context of a private household, involving at least one overnight stay; (ii) “Recreational activity”: Contact associated with a recreational activity, such as taking a walk, attending a family party, visiting a friend’s place (without an associated overnight stay), or contact during participation in a shared hobby (e.g. football); (iii) “School”: Contact in the context of a primary or secondary school, including contacts between students and teachers; (iv) “Kindergarten/Daycare”: Contact in the context of a kindergarten or daycare nursery (German “KiTa”), including contacts between children and daycare personnel; (v) “Workplace”: The involved individuals interacted in a work context; (vi) “Asylum/homeless care home”: The individuals had overlapping stays at a care home for asylum seekers or homeless individuals; (vii) “Elderly care home”: Contact in the context of a care home for the elderly, including contacts between care home personnel and inhabitants; (viii) “Hospital”: Contact in the context of a hospital, including contacts between patients and staff; (ix) “Assisted living care home”: Contact in the context of a care home for assisted living for non-elderly individuals (German “betreutes Wohnen”), including contacts between inhabitants and staff.

### Generation of a scrambled-edges control dataset for analysis of contact tracing genetic distances

To investigate whether genetic distances observed for case pairs connected by contact tracing edges may be the result of incidental genetic similarity of cases sequenced at approximately the same time, we generated a matched control dataset with randomized connections between sequenced cases registered at approximately the same time; the genetic distances along the randomized edges could then be compared to the genetic distances observed for real contact tracing edges. To generate the randomized dataset, we applied the following procedure. We identified all 3,862 cases that were sequenced and that either had a forward or backward contact tracing edge in the Integrated Case Graph. We created candidate randomized case pairs by first generating all 14,915,044 possible ordered case pairs from the identified 3,862 cases and removed all pairs that exhibited a registration date difference of more than 14 days. Furthermore, we removed all pairs that were connected by real forward or backward contact tracing links in the Integrated Case Graph, as well as all pairs linking a case to itself and case pairs *(a, b)* for which forward or backward contact tracing edges *(a, x)* and *(b, x)* – where *x* denotes a joint shared contact of *a* and *b* – existed. From the set of remaining 1,616,999 candidate randomized case pairs, we selected the 1,011 pairs that exhibited the highest degree of similarity (see Supplementary Fig. 9) to the 1,011 real case pairs connected by edges of type “forward contact tracing” or “backward contact tracing” (and with a genetic distance available) in the Integrated Case Graph, taking into account i) vaccination status, ii) date of index case registration, iii) mean case registration date, iv) distance between the case registration dates, v) number of registered contacts. The matching procedure was implemented using a custom R script and R packages “lubridate”, “stringr” and “data.table”.

### Retrospective investigation of outbreaks

For all outbreaks recorded by Düsseldorf Health Authority, routinely collected outbreak data, including the type of institution or context, the involved cases and the duration of the outbreak, were extracted from the database of Düsseldorf Heath Department and combined with the data represented in the Integrated Case Graph for analysis. During the study period, an outbreak record was created for three or more cases with with an epidemiological link outside the household setting. An outbreak was defined as “not genetically homogeneous” if a graph defined by (i) the sequenced outbreak cases as nodes and (ii) inter-case genetic distances as edges contained, after removal of edges corresponding to genetic distances > 1, more than one connected component; an individual case was classified as erroneously assigned to the outbreak if it was not part of the largest connected component of the graph so-defined (i.e., all genetically homogeneous outbreaks contained no erroneously assigned cases according to this definition). A community case (here defined as a case not part of any other outbreak of the same category as the outbreak under investigation) was defined as connected to an outbreak if it was connected to any case of the considered outbreak in the Integrated Case Graph; for outbreaks with ≥ 1 sequenced cases and all outbreak-linked sequenced community cases, the corresponding links were classified as “sequencing-confirmed” if the genetic distance between the linked community case and any sequenced outbreak case was *≤* 1. For outbreaks with ≥ 1 sequenced case, we also identified community cases that were only linked to the outbreak on a genetic basis, i.e. independent of epidemiological information; for this analysis, genetically linked community cases were defined as community cases that exhibited a genetic distance of 0, and that had a case registration date difference to any outbreak case of *≤* 14 days. Two outbreaks A and B were classified as epidemiologically linked if any case of outbreak A was linked to any case of outbreak B in the Integrated Case Graph; two outbreaks A and B were classified as genetically linked if any sequenced case of outbreak A had a genetic distance *≤* 1 to any sequenced case of outbreak B.

For school outbreaks, an in-depth analysis was carried out (Supplementary Figure 14), comprising the following steps and based on the definition of an outbreak as a group of cases connected by suspected or confirmed transmission events in a shared defined (typically institutional) context. First, epidemiological identification of cases falsely assigned to the outbreak: An evaluation of all available test and symptom onset dates for all outbreak cases was carried out and cases for which the available test and symptom onset data were not compatible with the other outbreak cases were identified. Second, identification of missing cases based on contact tracing and genetic data: For each outbreak with ≥ 1 sequenced case, we identified all cases that (i) were linked to at least one outbreak case by structured forward contact tracing data; (ii) were not already assigned to the outbreak; (iii) exhibited a genetic distance of 0 to at least one case in the outbreak; (iv) belonged, based on the available case data, to the same (typically institutional) context as the outbreak (e.g., were students or teachers at the school at which the outbreak had taken place). Third, analysis of non-structured and backward contact tracing data: For a subset of outbreaks (identified by column “Analysis of non-structured contact information” in table Supplementary Table 7), contact tracing information entered in a non-structured manner into the SurvNet application (i.e., using free-text entry fields) as well as backward contact tracing data were also analyzed during the previous step. Fourth, genetically driven identification of missing or linked cases: For a subset of outbreaks with ≥ 1 sequenced case (identified by column “Outbreak-external links investigated based on individual case genetic distance checks” in table Supplementary Table 7), we identified cases that (i) were not already linked to the outbreak by either direct assignment to the outbreak or via any contact tracing data; (ii) exhibited a genetic distance of ≤ 1 and a PCR test date difference of ≤ 4 weeks to at least one outbreak case; (iii) met one of the following three criteria of linkage to the outbreak: “missing outbreak case”, defined as belonging to the (institutional) context of the outbreak; “direct epidemiological linkage”, defined as a direct epidemiological link (e.g. shared household, family connection, workplace) to at least one outbreak case; “indirect epidemiological linkage”, defined as a direct epidemiological link to another case X with a direct epidemiological link to at least one outbreak case. For connections of the latter (indirect epidemiological) type, we furthermore required that “X” was either non-sequenced or that its viral genome was compatible (distance ≤ 1) to the outbreak; if “X” was not sequenced and found to represent the only connection between a sequenced case classified as “indirectly epidemiologically linked” and the outbreak, “X” was classified as an outbreak-linked case with indirect genetic support and further classified into a missing outbreak case or a linked community sample. Fifth, assessment of potential outbreak-associated transmission chains: We carried out a comprehensive manual assessment and analysis of potential infection chains associated with the investigated outbreaks. This led, for example, to the conclusion that the only sequenced case of outbreak CVD-DÜS-SCHULE-2021-0182 was likely a mis-assigned (see Supplementary Note 1). Sixth, identification of related outbreaks: Two outbreaks were classified as related if there was, according to the criteria defined above, at least one case from the one outbreak that exhibited direct or indirect genetic linkage to at least one case from the other outbreak and if the connection between the two outbreaks was also supported by a direct or indirect epidemiological connection between at least one case from the one outbreak to at least one case from the other outbreak.

### Cluster Explanation Rate

Given a single cluster C of N genetically related cases, and a set of epidemiological connections between them, the Cluster Explanation Rate (CER) measures the proportion of cases in the cluster that a putative infection source can be identified for based on a hypothetical transmission chain constructed from the epidemiological connections for the cases in the cluster. When generalized to multiple clusters, the CER is computed by summing over the number of cases with a putative infection source and the total number of cases across the considered clusters. Finally, at the level of a complete dataset, the CER is determined by first clustering all sequenced cases by genetic similarity and then computing the CER of the clusters so-determined; at this level, the CER provides a measure of how well the collected epidemiological data explain overall observed genetic similarities.

Formally, let *G’= (V, E’)* be a directionalized and augmented version of the case graph *C= (V, E)*; directionalization and augmentation are required for ensuring that hypothetical transmission chains considered for computing the CER are consistent with respect to symptom and/or case registration dates and for including hypothetical transmission chains that involve intermediate non-sequenced cases (see below). All CER computations are based on *G’*. We first define the CER of a single cluster *C*, i.e. of a subset of nodes of the case graph *G’*, of genetically related cases; let *t_g_* denote the genetic distance threshold that was used for the clustering (defined in detail below). Let the set *E’(C)* represent the set of genetically supported epidemiological connections between the cases in *C*; *E^’^(C)* is defined as the subset of directed edges between *c* in *G’* that connect two cases with a pairwise genetic distance *≤ t_g_*. A hypothetical transmission chain *h* is defined as an acyclic subset of *E’(C)* such that no node *c* ε *c* has more than one incoming edge from the set *h* (note that the definition allows for partial transmission chains that do not connect all nodes; epidemiologically, such transmission chains may be interpreted to include cases outside of the cluster *C*; Supplementary Figure 20; in addition, no constraints are placed on the number of outgoing edges, i.e. the set of considered transmission chains includes “branching” transmission chains or “networks”). The CER for cluster *C* in case graph *G’* is then defined as

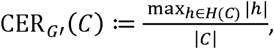

where *H(C)* is the set of all possible hypothetical transmission chains for the cases in cluster *C* and their epidemiological connections *E’(C)*; *|h|* is the size of the hypothetical transmission chain *h* in edges; *|C|* is the number of cases in *c*; and *max_hϵH_ |h|* is computed according to the heuristic algorithm defined below.

For a set *W* of multiple clusters of genetically related cases in case graph *G’*, we define the CER as

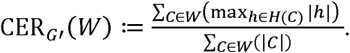

Finally, to define the CER for a case graph *G’*, the sequenced cases in *G’* are clustered into groups of genetically related isolates, based on a similarity threshold *t_g_*. To do so, we consider a graph that consists of the sequenced cases in *G’* as nodes and that includes all pairwise genetic distances as undirected edges; we remove all edges corresponding to genetic distances > *t_g_*; and, finally, we define the set *genetic_clusters(G’)* as the set of remaining connected components with size > 1 in the graph. Any two cases *c*_1_ and *c*_2_ from the same cluster either have genetic similarity *≤ t_g_* or they are connected by a path of sequenced cases with intermediate pairwise distances between *≤ t_g_*; furthermore, the computed clusters are non-overlapping. Based on the computed clusters, we define the CER for a case graph *G’* as

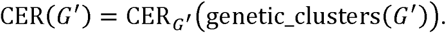

Directionalization, yielding a directed case graph *G’= (V, E’)*, enables the definition of hypothetical transmission chains at the level of a directed graph between cases, capturing the concept of directionality of transmission and ensuring that all considered pairwise links between cases are consistent with respect to symptom onset (or, if unavailable, case registration) dates. In *G’*, a directed edge *(c*_1_*, c*_2_) represents a possible hypothetical transmission event from *c*_1_ to *c*_2_. Directionalization is followed by potential augmentation of the graph with “jump edges”, representing links in hypothetical transmission chains between sequenced cases that involve intermediate non-sequenced cases. For directionalization, *E^’^* is constructed by including, for each (undirected) edge *e= (c*_1_*, c*_2_*) E E*, one or two directed edges, as defined below. For all edges except for edges of type “backward contact tracing”, directionalization is controlled by the parameter *t_d_*, specifying the allowed difference in symptom onset dates in days between linked cases (according to the definition below). Denote the date of symptom onset of case *c* as *s(c)*; if the symptom onset date for case *c* is unavailable, *s(c)* is set to the case registration date of *c* minus 3 days, based on an empirical analysis of the difference between symptom onset and case registration dates for samples for which both dates are available (Supplementary Fig. 23). We distinguish between the following cases: (i) if abs*(s(c*_1_*) - s(c*_2_*)) ≤ t_d_*, edges *(c*_1_*, c*_2_) and *(c*_2_*, c*_1_) are added to *E^’^*; (ii) if *s(c*_1_*) - s(c*_2_*) > t_d_*, edge *(c*_2_*, c*_1_) is added to *E^’^*; (iii) otherwise, *s(c*_2_*) - s(c*_1_*) > t_d_* is true, and edge *(c*_1_*, c*_2_) is added to *E^’^*. Note that these definitions imply that one undirected edge in the original case graph may be represented by two directed edges in the directed case graph, if the observed symptom onset dates are consistent with transmission in both directions, depending on the threshold *t_d_*. For undirected edges *(c*_1_*, c*_2_) of type “backward contact tracing” in the original case graph *G*, a directed edge from *c*_1_ to *c*_2_ is added to *E^’^* if case *c*_2_ specified case *c*_1_ as their assumed infection source during infection source during backward contact tracing; and a directed edge from *c*_2_ to *c*_l_ is added to *E^’^* if case *c*_1_ specified case *c*_2_ as their assumed infection source during backward contact tracing. For augmentation with “jump distance” parameter *t_j_ =* 0, no additional edges are added to *G’*. For augmentation with *t_j_ =* 1, an edge from *c*_1_ to *c*_2_ is added to *E^’^* for all instances of the form *(c*_1_*, a, c*_2_), in which (i) *c*_1_ and *c*_2_ represent two sequenced cases; (ii) *a* represents a non-sequenced case; (iii) there are no edges directly connecting *c*_1_ to *c*_2_; (iv) there is at least one edge from *c*_1_ to *a*; (v) there is at least one edge from *a* to *c*_2_; (v) the genetic distance between *c*_1_ to *c*_2_ is ≤ *(t_j_ +* 1*) * t_g_*. Augmentation for *t_j_ =*2 proceeds analogously for instances of the form *(c*_1_*, a, b, c*_2_).

The computed *CER(G’)* depends on the parameter used for the genetic clustering (*t_g_*), the “maximum symptom onset difference” parameter used for directionalization (*t_d_*), and the “jump distance” parameter (*t_j_*). To assess the impact of different parameter choices on the computed CER, all CER computations are carried out for all combinations (i.e., the outer product) of the parameter values *t_g_ = {*0,1*}*, *t_d_ = {*0,3*}* and *t_j_ = {*0,1,2*}*. For *t_d_*, a value of 0 implies constraining the direction of hypothetical transmission whenever there is any difference in symptom onset dates (i.e., a conservative choice); -3 was chosen empirically based on inspection of observed symptom onset dates for sample pairs connected by edges of type “backward contact tracing”, which are intrinsically directed (Supplementary Fig. 23). Directionalization results for different edge types and the two employed thresholds are visualized in Supplementary Figure 21.

max*_h_*_∈_*_H(c)_ |h|* for a cluster *C* is computed according to the following procedure. Initialize the variable *h’* as the empty set; *h’* is populated progressively with edges from *E’(C)*, and, after termination, the set *h^’^* represents a heuristic approximation of arg max*_h_*_∈_*_H(C)_ |h|*.

The following steps are carried out until termination. First, define *M* as the set of nodes *c* ∈ *c* that are neither the origin nor the destination of any of the edges currently in *h’*. Second, for each *c* ∈ *M*, define reachable*(c)* as the set of other nodes in *M* that can be reached from *c*, employing paths that consist of the directed edges *E’(c)* and that only involve intermediate nodes that are also part of *M*. reachable*(c)* can be computed using a depth-first search. Third, find *c_max_ =* arg max*_c∈M_ |*reachable*(c)|* by iterating through the current members of *M*. If *|*reachable*(c_max_)| =* 0, terminate; otherwise, let *B* be the set of edges underlying the paths connecting *c_max_* to reachable*(c)*. If necessary, remove edges from *B* so that *B* does not contain any cycles and so that there is no node in reachable*(c)* with more than one incoming edge from *B*. Finally, add the edges in *B* to *h^’^* and go back to the first step.

## Data Availability

Code developed during the present study is available upon request to the authors.

## Ethics statement

This study was approved by the ethics committee of the Medical Faculty of Heinrich Heine University Düsseldorf (#2020–839).

## No competing interests

All authors have completed the ICMJE uniform disclosure form and declare: no support from any organization for the submitted work; no financial relationships with any organizations that might have an interest in the submitted work in the previous three years; no other relationships or activities that could appear to have influenced the submitted work.

## Acknowledgements

We thank Gil McVean, Adam Phillippy and Marc Janssen for helpful discussions. We also thank the contact tracing team of Düsseldorf Health Authority for their commitment and the countless extra hours.

## Data and code availability

All Python and R custom scripts were uploaded to github (https://github.com/DiltheyLab/SARS-CoV-2-GenomicsEnhancedContactTracing-Code). Since all individual data remains with the health authorities in Düsseldorf, we included a set of random mock data for reproducibility.

## References

Allen, K., A. Marmor and D. Pourmarzi (2022). “Pens down: An outbreak of the B.1.617.2 SARS-CoV-2 variant in an Australian high school, August 2021.“ Commun Dis Intell (2018) 46.

Andersen, K. G., B. J. Shapiro, C. B. Matranga, R. Sealfon, A. E. Lin, L. M. Moses, O. A. Folarin, A. Goba, I. Odia, P. E. Ehiane, M. Momoh, E. M. England, S. Winnicki, L. M. Branco, S. K. Gire, E. Phelan, R. Tariyal, R. Tewhey, O. Omoniwa, M. Fullah, R. Fonnie, M. Fonnie, L. Kanneh, S. Jalloh, M. Gbakie, S. Saffa, K. Karbo, A. D. Gladden, J. Qu, M. Stremlau, M. Nekoui, H. K. Finucane, S. Tabrizi, J. J. Vitti, B. Birren, M. Fitzgerald, C. McCowan, A. Ireland, A. M. Berlin, J. Bochicchio, B. Tazon-Vega, N. J. Lennon, E. M. Ryan, Z. Bjornson, D. A. Milner, Jr., A. K. Lukens, N. Broodie, M. Rowland, M. Heinrich, M. Akdag, J. S. Schieffelin, D. Levy, H. Akpan, D. G. Bausch, K. Rubins, J. B. McCormick, E. S. Lander, S. Gunther, L. Hensley, S. Okogbenin, C. Viral Hemorrhagic Fever, S. F. Schaffner, P. O. Okokhere, S. H. Khan, D. S. Grant, G. O. Akpede, D. A. Asogun, A. Gnirke, J. Z. Levin, C. T. Happi, R. F. Garry and P. C. Sabeti (2015). “Clinical Sequencing Uncovers Origins and Evolution of Lassa Virus.” Cell 162(4): 738–750.

Bludau, A., A. Jack, N. Fischer, J. Dreesman, C. Drosten, R. Egelkamp, L. Ehlkes, F. Feil, A. Grundhoff, H. Grundmann, P. Kreuzer, M. Monazahian, I. Overesch, D. Schmitt, M. Troger, A. von Reiswitz, J. Weber, A. Dilthey, C. Hornberg, S. Reuter and S. Scheithauer (2025). “Use of integrated genomic surveillance by local public health authorities: Recommendations based on a mixed-methods study of current adoption, applications and success factors, Germany, 2023.” Euro Surveill 30(13).

COVID-19 Genomics UK (COG-UK) (2020). “An integrated national scale SARS-CoV-2 genomic surveillance network.“ Lancet Microbe 1(3): e99–e100.

Faensen, D., H. Claus, J. Benzler, A. Ammon, T. Pfoch, T. Breuer and G. Krause (2006). “SurvNet@RKI--a multistate electronic reporting system for communicable diseases.” Euro Surveill 11(4): 100–103.

Faria, N. R., E. C. Sabino, M. R. Nunes, L. C. Alcantara, N. J. Loman and O. G. Pybus (2016). “Mobile real-time surveillance of Zika virus in Brazil.” Genome Med 8(1): 97.

Ferretti, L., C. Wymant, J. Petrie, D. Tsallis, M. Kendall, A. Ledda, F. Di Lauro, A. Fowler, A. Di Francia, J. Panovska-Griffiths, L. Abeler-Dorner, M. Charalambides, M. Briers and C. Fraser (2024). “Digital measurement of SARS-CoV-2 transmission risk from 7 million contacts.” Nature 626(7997): 145–150.

Garcia-Bernardo, J., C. Hedde-von Westernhagen, T. Emery and A. J. van Hoek (2024). “Assessing COVID-19 transmission through school and family networks using population-level registry data from the Netherlands.“ Sci Rep 14(1): 31248.

Gudbjartsson, D. F., A. Helgason, H. Jonsson, O. T. Magnusson, P. Melsted, G. L. Norddahl, J. Saemundsdottir, A. Sigurdsson, P. Sulem, A. B. Agustsdottir, B. Eiriksdottir, R. Fridriksdottir, E. E. Gardarsdottir, G. Georgsson, O. S. Gretarsdottir, K. R. Gudmundsson, T. R. Gunnarsdottir, A. Gylfason, H. Holm, B. O. Jensson, A. Jonasdottir, F. Jonsson, K. S. Josefsdottir, T. Kristjansson, D. N. Magnusdottir, L. le Roux, G. Sigmundsdottir, G. Sveinbjornsson, K. E. Sveinsdottir, M. Sveinsdottir, E. A. Thorarensen, B. Thorbjornsson, A. Love, G. Masson, I. Jonsdottir, A. D. Moller, T. Gudnason, K. G. Kristinsson, U. Thorsteinsdottir and K. Stefansson (2020). “Spread of SARS-CoV-2 in the Icelandic Population.” N Engl J Med 382(24): 2302–2315.

Hall, R. N., A. Jones, E. Crean, V. Marriott, N. Pingault, A. Marmor, T. Sloan-Gardner, K. Kennedy, K. Coleman, V. Johnston and B. Schwessinger (2023). “Public health interventions successfully mitigated multiple incursions of SARS-CoV-2 Delta variant in the Australian Capital Territory.“ Epidemiol Infect 151: e30.

Harris, S. R., E. J. Cartwright, M. E. Torok, M. T. Holden, N. M. Brown, A. L. Ogilvy-Stuart, M. J. Ellington, M. A. Quail, S. D. Bentley, J. Parkhill and S. J. Peacock (2013). “Whole-genome sequencing for analysis of an outbreak of meticillin-resistant Staphylococcus aureus: a descriptive study.” Lancet Infect Dis 13(2): 130–136.

Hjorleifsson, K. E., S. Rognvaldsson, H. Jonsson, A. B. Agustsdottir, M. Andresdottir, K. Birgisdottir, O. Eiriksson, E. S. Eythorsson, R. Fridriksdottir, G. Georgsson, K. R. Gudmundsson, A. Gylfason, G. Haraldsdottir, B. O. Jensson, A. Jonasdotti, A. Jonasdottir, K. S. Josefsdottir, N. Kristinsdottir, B. Kristjansdottir, T. Kristjansson, D. N. Magnusdottir, R. Palsson, L. le Roux, G. M. Sigurbergsdottir, A. Sigurdsson, M. I. Sigurdsson, G. Sveinbjornsson, E. A. Thorarensen, B. Thorbjornsson, M. Thordardottir, A. Helgason, H. Holm, I. Jonsdottir, F. Jonsson, O. T. Magnusson, G. Masson, G. L. Norddahl, J. Saemundsdottir, P. Sulem, U. Thorsteinsdottir, D. F. Gudbjartsson, P. Melsted and K. Stefansson (2022). “Reconstruction of a large-scale outbreak of SARS-CoV-2 infection in Iceland informs vaccination strategies.” Clin Microbiol Infect 28(6): 852–858.

Houwaart, T., S. Belhaj, E. Tawalbeh, D. Nagels, Y. Frohlich, P. Finzer, P. Ciruela, A. Sabria, M. Herrero, C. Andres, A. Anton, A. Benmoumene, D. Asskali, H. Haidar, J. von Dahlen, J. Nicolai, M. Stiller, J. Blum, C. Lange, C. Adelmann, B. Schroer, U. Osmers, C. Grice, P. P. Kirfel, H. Jomaa, D. Strelow, L. Hulse, M. Pigulla, P. Kreuzer, A. Tyshaieva, J. Weber, T. Wienemann, M. Kohns Vasconcelos, K. Hoffmann, N. Lubke, S. Hauka, M. Andree, C. J. Scholz, N. Jazmati, K. Gobels, R. Zotz, K. Pfeffer, J. Timm, L. Ehlkes, A. Walker, A. T. Dilthey, C.-O. I. German and C.-O. I. German (2022). “Integrated genomic surveillance enables tracing of person-to-person SARS-CoV-2 transmission chains during community transmission and reveals extensive onward transmission of travel-imported infections, Germany, June to July 2021.” Euro Surveill 27(43).

Katoh, K. and D. M. Standley (2013). “MAFFT multiple sequence alignment software version 7: improvements in performance and usability.” Mol Biol Evol 30(4): 772–780.

Krause, G., D. Altmann, D. Faensen, K. Porten, J. Benzler, T. Pfoch, A. Ammon, M. H. Kramer and H. Claus (2007). “SurvNet electronic surveillance system for infectious disease outbreaks, Germany.” Emerg Infect Dis 13(10): 1548–1555.

Lane, C. R., N. L. Sherry, A. F. Porter, S. Duchene, K. Horan, P. Andersson, M. Wilmot, A. Turner, S. Dougall, S. A. Johnson, M. Sait, A. Goncalves da Silva, S. A. Ballard, T. Hoang, T. P. Stinear, L. Caly, V. Sintchenko, R. Graham, J. McMahon, D. Smith, L. E. Leong, E. M. Meumann, L. Cooley, B. Schwessinger, W. Rawlinson, S. J. van Hal, N. Stephens, M. Catton, C. Looker, S. Crouch, B. Sutton, C. Alpren, D. A. Williamson, T. Seemann and B. P. Howden (2021). “Genomics-informed responses in the elimination of COVID-19 in Victoria, Australia: an observational, genomic epidemiological study.“ Lancet Public Health 6(8): e547–e556.

Leclerc, Q. J., N. M. Fuller, L. E. Knight, C. C.-W. Group, S. Funk and G. M. Knight (2020). “What settings have been linked to SARS-CoV-2 transmission clusters?” Wellcome Open Res 5: 83.

Manica, M., P. Poletti, S. Deandrea, G. Mosconi, C. Ancarani, S. Lodola, G. Guzzetta, V. d’Andrea, V. Marziano, A. Zardini, F. Trentini, A. Odone, M. Tirani, M. Ajelli and S. Merler (2023). “Estimating SARS-CoV-2 transmission in educational settings: A retrospective cohort study.” Influenza Other Respir Viruses 17(1): e13049.

Mate, S. E., J. R. Kugelman, T. G. Nyenswah, J. T. Ladner, M. R. Wiley, T. Cordier-Lassalle, A. Christie, G. P. Schroth, S. M. Gross, G. J. Davies-Wayne, S. A. Shinde, R. Murugan, S. B. Sieh, M. Badio, L. Fakoli, F. Taweh, E. de Wit, N. van Doremalen, V. J. Munster, J. Pettitt, K. Prieto, B. W. Humrighouse, U. Stroher, J. W. DiClaro, L. E. Hensley, R. J. Schoepp, D. Safronetz, J. Fair, J. H. Kuhn, D. J. Blackley, A. S. Laney, D. E. Williams, T. Lo, A. Gasasira, S. T. Nichol, P. Formenty, F. N. Kateh, K. M. De Cock, F. Bolay, M. Sanchez-Lockhart and G. Palacios (2015). “Molecular Evidence of Sexual Transmission of Ebola Virus.” N Engl J Med 373(25): 2448–2454.

McCrone, J. T., V. Hill, S. Bajaj, R. E. Pena, B. C. Lambert, R. Inward, S. Bhatt, E. Volz, C. Ruis, S. Dellicour, G. Baele, A. E. Zarebski, A. Sadilek, N. Wu, A. Schneider, X. Ji, J. Raghwani, B. Jackson, R. Colquhoun, A. O’Toole, T. P. Peacock, K. Twohig, S. Thelwall, G. Dabrera, R. Myers, C.-G. U. Consortium, N. R. Faria, C. Huber, Bogoch, II, K. Khan, L. du Plessis, J. C. Barrett, D. M. Aanensen, W. S. Barclay, M. Chand, T. Connor, N. J. Loman, M. A. Suchard, O. G. Pybus, A. Rambaut and M. U. G. Kraemer (2022). “Context-specific emergence and growth of the SARS-CoV-2 Delta variant.” Nature 610(7930): 154–160.

Mellmann, A., S. Bletz, T. Boking, F. Kipp, K. Becker, A. Schultes, K. Prior and D. Harmsen (2016). “Real-Time Genome Sequencing of Resistant Bacteria Provides Precision Infection Control in an Institutional Setting.” J Clin Microbiol 54(12): 2874–2881.

Mossong, J., N. Hens, M. Jit, P. Beutels, K. Auranen, R. Mikolajczyk, M. Massari, S. Salmaso, G. S. Tomba, J. Wallinga, J. Heijne, M. Sadkowska-Todys, M. Rosinska and W. J. Edmunds (2008). “Social contacts and mixing patterns relevant to the spread of infectious diseases.” PLoS Med 5(3): e74.

Nicolai, J., A. J. Miller, N. Hahn, J. Fazaal, A. Bunte, J. Silvery, M. Rosenstengel, P. Hoffmann, K. U. Ludwig, C. Tiemann, L. E. Rose and A. T. Dilthey (2025). “Large-scale SARS-CoV-2 sequencing indicates prior community circulation of the viral strain associated with Germany’s largest meat processing plant.“ Sci Rep 15(1): 38466.

Phuong, H. T., A. Bartz, A. K. Jarynowski, B. Lange, C. I. Jarvis, N. Rubsamen, R. T. Mikolajczyk, S. Scholz, T. Berger, T. Heinsohn, V. Belik, A. Karch and V. K. Jaeger (2025). “Changes in social contact patterns in Germany during the SARS-CoV-2 pandemic - an analysis based on the COVIMOD study.” BMC Infect Dis 25(1): 588.

Pullano, G., E. Valdano, N. Scarpa, S. Rubrichi and V. Colizza (2020). “Evaluating the effect of demographic factors, socioeconomic factors, and risk aversion on mobility during the COVID-19 epidemic in France under lockdown: a population-based study.” Lancet Digit Health 2(12): e638–e649.

Rambaut, A., E. C. Holmes, A. O’Toole, V. Hill, J. T. McCrone, C. Ruis, L. du Plessis and O. G. Pybus (2020). “A dynamic nomenclature proposal for SARS-CoV-2 lineages to assist genomic epidemiology.” Nat Microbiol 5(11): 1403–1407.

Rodiah, I., P. Vanella, A. Kuhlmann, V. K. Jaeger, M. Harries, G. Krause, A. Karch, W. Bock and B. Lange (2023). “Age-specific contribution of contacts to transmission of SARS-CoV-2 in Germany.” Eur J Epidemiol 38(1): 39–58.

Ryu, S., D. Kim, J. S. Lim, S. T. Ali and B. J. Cowling (2022). “Serial Interval and Transmission Dynamics during SARS-CoV-2 Delta Variant Predominance, South Korea.” Emerg Infect Dis 28(2): 407–410.

Seemann, T., C. R. Lane, N. L. Sherry, S. Duchene, A. Goncalves da Silva, L. Caly, M. Sait, S. A. Ballard, K. Horan, M. B. Schultz, T. Hoang, M. Easton, S. Dougall, T. P. Stinear, J. Druce, M. Catton, B. Sutton, A. van Diemen, C. Alpren, D. A. Williamson and B. P. Howden (2020). “Tracking the COVID-19 pandemic in Australia using genomics.” Nat Commun 11(1): 4376.

Shu, Y. and J. McCauley (2017). “GISAID: Global initiative on sharing all influenza data - from vision to reality.” Euro Surveill 22(13).

Sivertsen, A., N. Mortensen, U. Solem, E. Valen, M. F. Bullita, K. A. Wensaas, S. Litleskare, G. Rortveit, H. M. S. Grewal and E. Ulvestad (2024). “Comprehensive contact tracing during an outbreak of alpha-variant SARS-CoV-2 in a rural community reveals less viral genomic diversity and higher household secondary attack rates than expected.” mSphere 9(8): e0011424.

Walker, A., T. Houwaart, P. Finzer, L. Ehlkes, A. Tyshaieva, M. Damagnez, D. Strelow, A. Duplessis, J. Nicolai, T. Wienemann, T. Tamayo, M. Kohns Vasconcelos, L. Hulse, K. Hoffmann, N. Lubke, S. Hauka, M. Andree, M. P. Daumer, A. Thielen, S. Kolbe-Busch, K. Gobels, R. Zotz, K. Pfeffer, J. Timm, A. T. Dilthey and C.-O. I. German (2022). “Characterization of Severe Acute Respiratory Syndrome Coronavirus 2 (SARS-CoV-2) Infection Clusters Based on Integrated Genomic Surveillance, Outbreak Analysis and Contact Tracing in an Urban Setting.” Clin Infect Dis 74(6): 1039–1046.

Wong, N. S., S. S. Lee, T. H. Kwan and E. K. Yeoh (2020). “Settings of virus exposure and their implications in the propagation of transmission networks in a COVID-19 outbreak.” Lancet Reg Health West Pac 4: 100052.

