## Supplementary Figures for "Genomics-enhanced contact tracing enabled the characterization of SARS-CoV-2 transmission chains and infection contexts in the general population during community transmission"

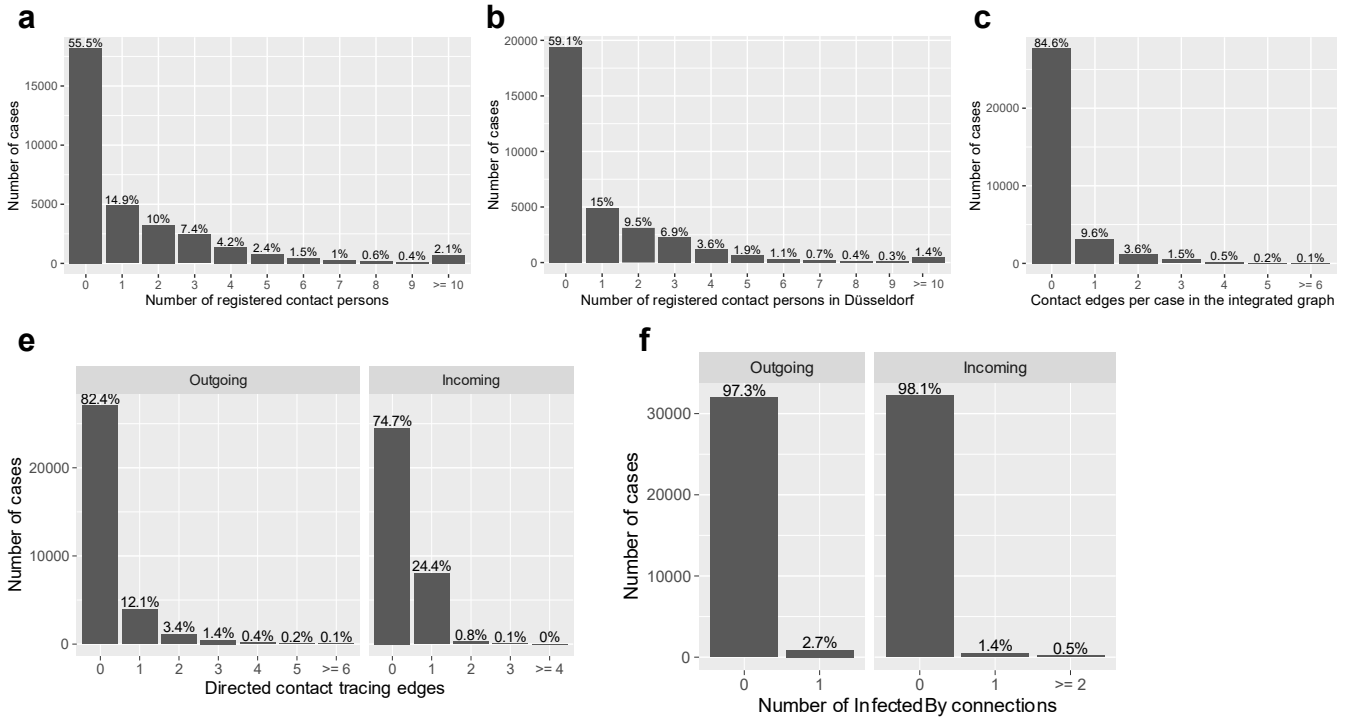

**Supplementary Figure 1: Forward and backward contact tracing.** **a**, Histogram over the number of registered forward contact tracing records per case. **b**, Histogram over the number of registered forward contact tracing records per case, limited to contacts registered (residential address) in the city Düsseldorf. **c**, Histogram over the number of "forward contact tracing"-type edges per case in the Integrated Case Graph. **d**, Histogram over the number of registered incoming or outgoing forward contact tracing links per case in the directionalized ($t_d = 0$; see Methods) version of the Integrated Case Graph. **e**, Histograms over the number of "backward contact tracing"-type edges per case in the directionalized version of the Integrated Case Graph, stratified by direction of the edge.

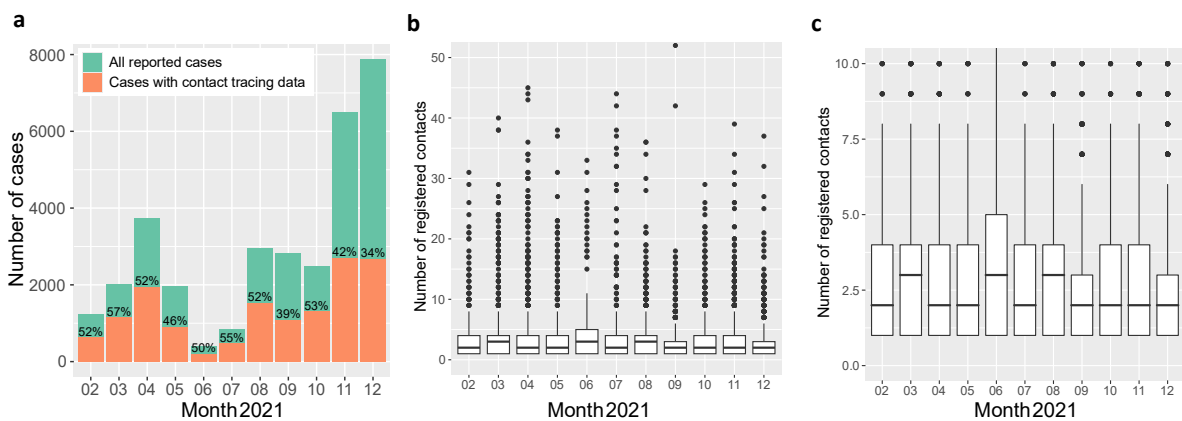

**Supplementary Figure 2: Forward contact tracing records over time.** **a**, Shown is the monthly total number of registered SARS-CoV-2 cases registered and the proportion of cases with at least one attached forward contact tracing record (independent of whether the contact was mapped onto another case or not). **b**, Distribution over the number of forward contact tracing records per case, limited to cases with at least one attached contact tracing record and stratified by month of the year. **c**, Same data as in panel B., with a truncated y-axis focusing on the region between 1 and 10 forward contact tracing records.

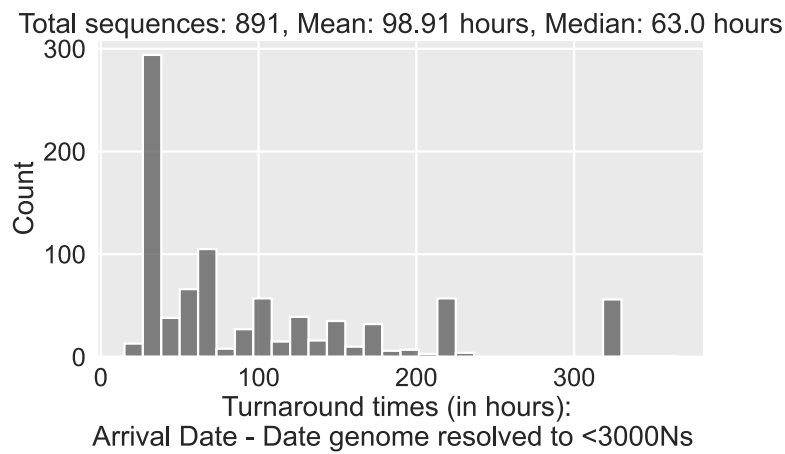

**Supplementary Figure 3: SARS-CoV-2 sequencing turnaround times.** Distribution of SARS-CoV-2 sequencing turnaround times, measured from SARS-CoV-2 sample arrival at Heinrich Heine University to first availability of a resolved SARS-CoV-2 genome with <3000 unresolved (i.e. “N”) characters on the dashboard of the Integrated Genomic Surveillance System Düsseldorf, tracked for a subset of $n = 891$ samples that were collected between April and June 2022. Samples that were not successfully sequenced to the desired level of SARS-CoV-2 genome completeness were not included.

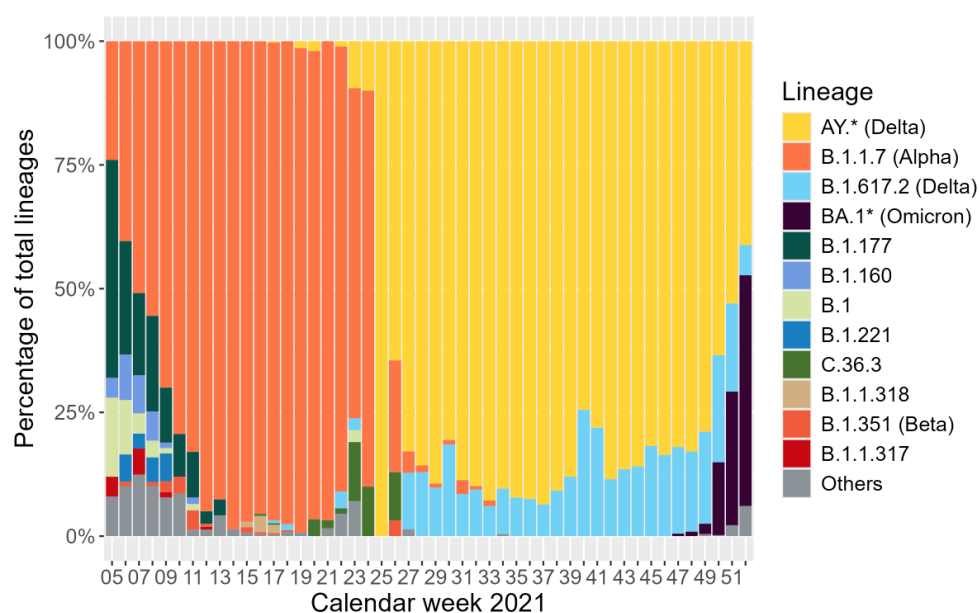

**Supplementary Figure 5: SARS-CoV-2 variant distribution over time.** Pango lineages [Rambaut et al. 2020] of the sequenced SARS-CoV-2 genomes, stratified by week.

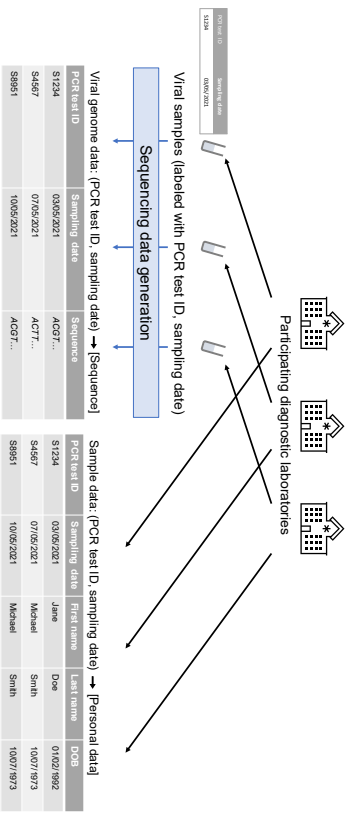

Patient/resolved viral genome data: (First name, last name, DOB) → [Sequence]

| First name | Last name | DOB | Sampling date | Sequence |
| --- | --- | --- | --- | --- |
| Jane | Smith | 01/02/1992 | 03/05/2021 | ACGT... |
| Michael | Smith | 10/07/1973 | 07/05/2021 | ACGT... |
| Michael | Smith | 10/07/1973 | 10/05/2021 | ACGT... |

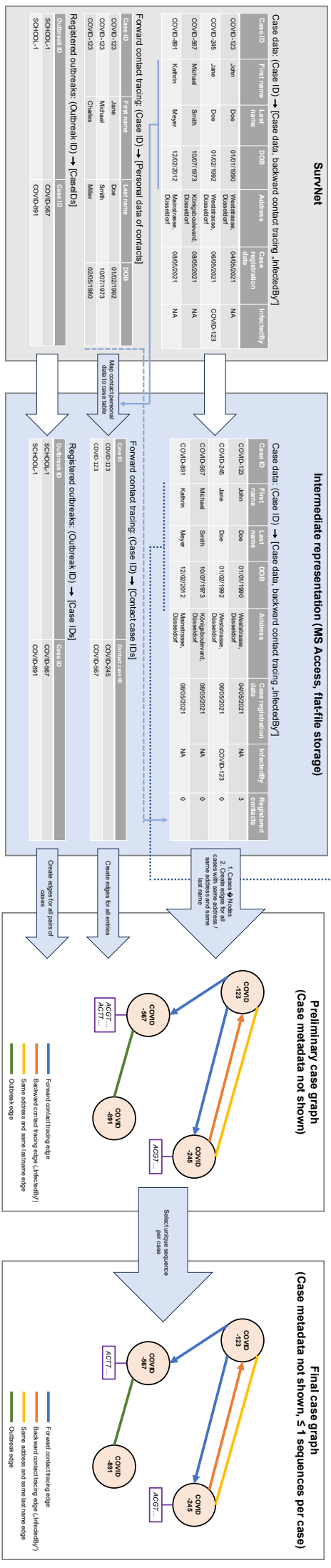

**Supplementary Figure 4: Data integration approach.** Shown are the different steps of data integration leading to the Integrated Case Graph. Case, contact tracing and outbreak data were exported from the SurvNet database hosted at Düsseldorf Health Department and transferred into an intermediate MS Access application. SARS-CoV-2 samples were labeled with PCR test IDs and sampling dates; based on these, the first and last names and dates of births of the tested individuals could be obtained and merged with Düsseldorf Health Department case registration data. All case- or sample-identifying data (incl. names, dates of birth, sampling dates, address data) are hypothetical. In a final step, if a case was sequenced more than once, a unique sequence

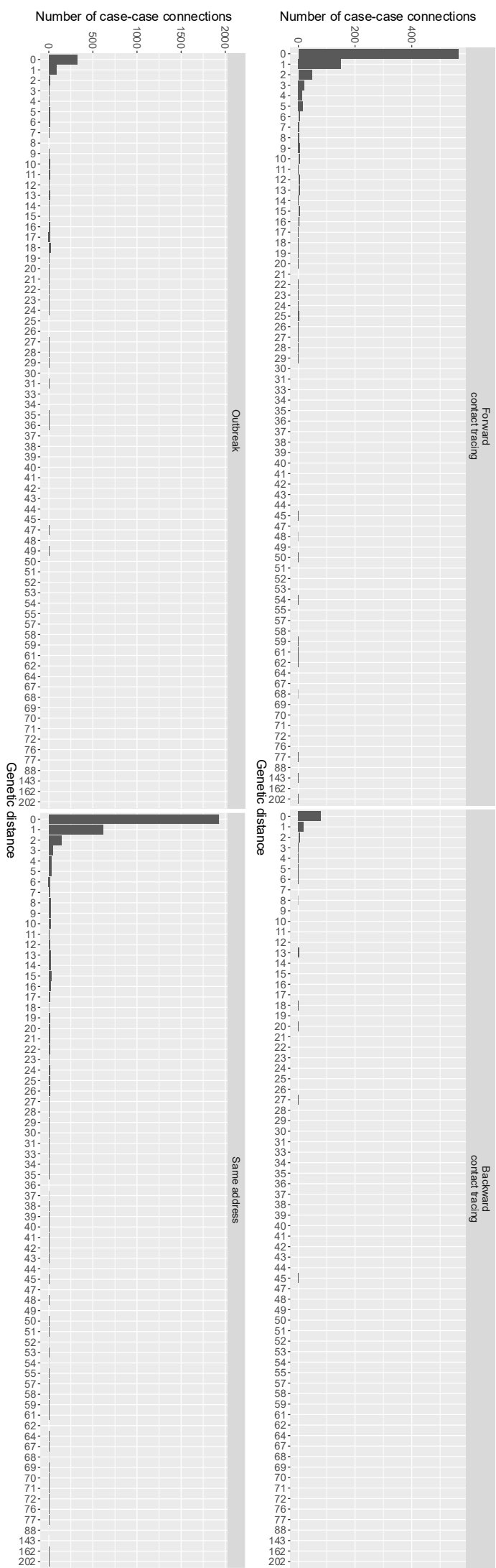

**Supplementary Figure 6: Distribution of genetic distances for contact tracing and “sameAddress”-type edges.** Distribution of SARS-CoV-2 genetic distances for case pairs linked by different types of edges in the graph, stratified by edge type and only shown for case pairs for which a genetic distance could be calculated.

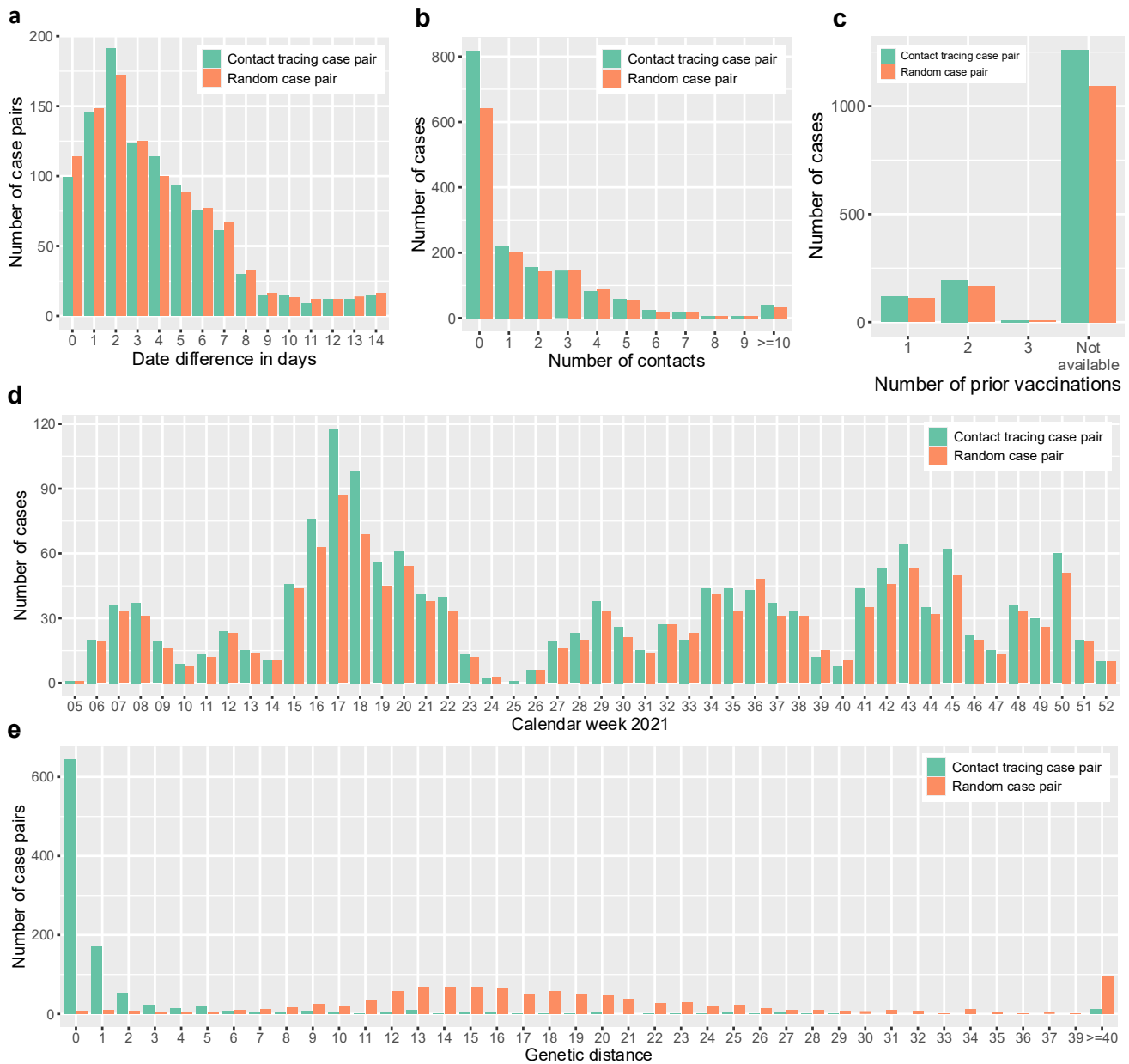

**Supplementary Figure 7: Matched control dataset with scrambled contact tracing links.** **a**, Difference between case registration dates for pairs of sequenced cases connected by real forward or backward contact tracing links and scrambled contact tracing edges. **b**, Number of contacts per case, comparing cases that are part of pairs of sequenced cases connected by real or scrambled contact tracing edges. **c**, Vaccination status of cases, comparing cases that are part of pairs of sequenced cases connected by real or scrambled contact tracing edges. **d**, Registration date of cases, comparing cases that are part of pairs of sequenced cases connected by real or scrambled contact tracing edges. **e**, Genetic differences between cases, comparing cases that are part of pairs of sequenced cases connected by real or scrambled contact tracing edges.

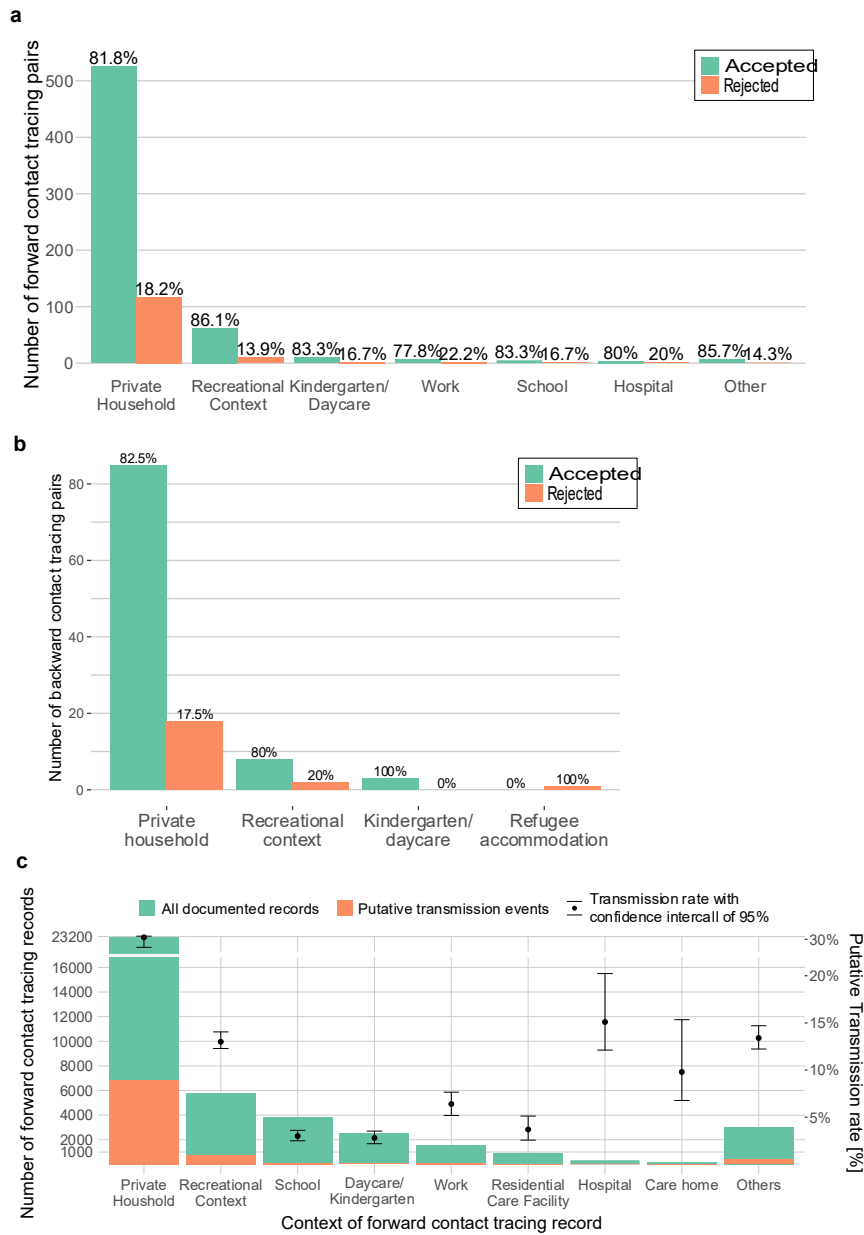

**Supplementary Figure 8: Contact tracing infection contexts, genetic evaluation and transmission rates.** **a**, Genetic validation of infection contexts captured by forward tracing; shown is the number of case pairs that were connected by a forward contact tracing edge assigned to the corresponding putative context, stratified by whether the genetic distance between the connected cases was $\leq 1$ ("accepted") or $> 1$ ("rejected"); edges for which no genetic distance was available were not included. **b**, Genetic validation of infection contexts captured by backward contact tracing; shown is the number of case pairs that were connected by a backward contact tracing edge assigned to the corresponding putative context; genetic validation analogous to panel A. **c**, Forward contact tracing contacts by context and associated putative context-specific transmission rates. Putative transmission rates were calculated for index cases registered until 17 December 2021.

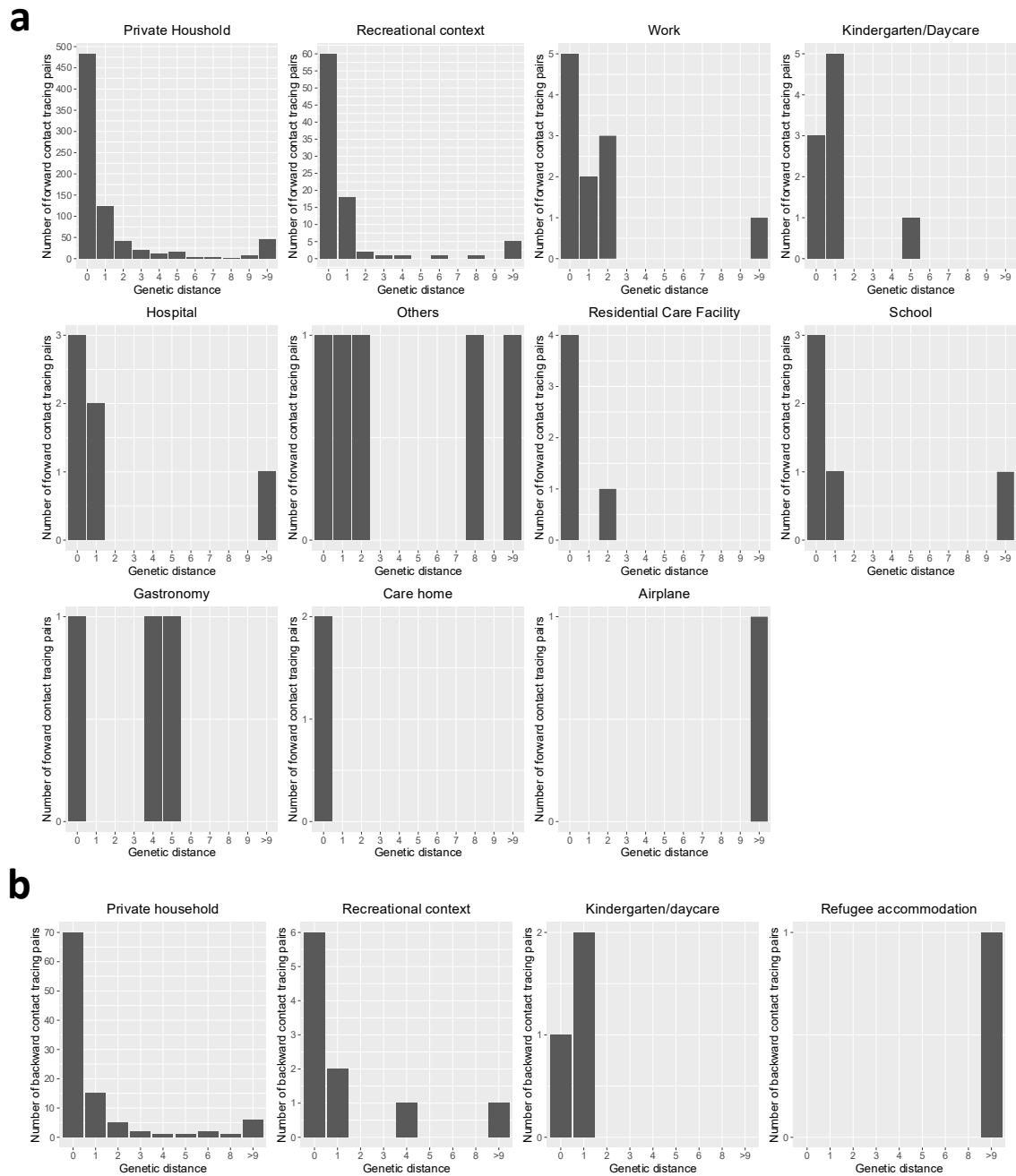

**Supplementary Figure 9: Context-specific distribution of genetic distances for contact tracing edges.** Distribution of SARS-CoV-2 genetic distances for case pairs linked by forward (panel A) and backward (panel B) contact tracing edges in the graph, stratified by edge type and infection contexts, and only shown for case pairs for which a genetic distance could be calculated.

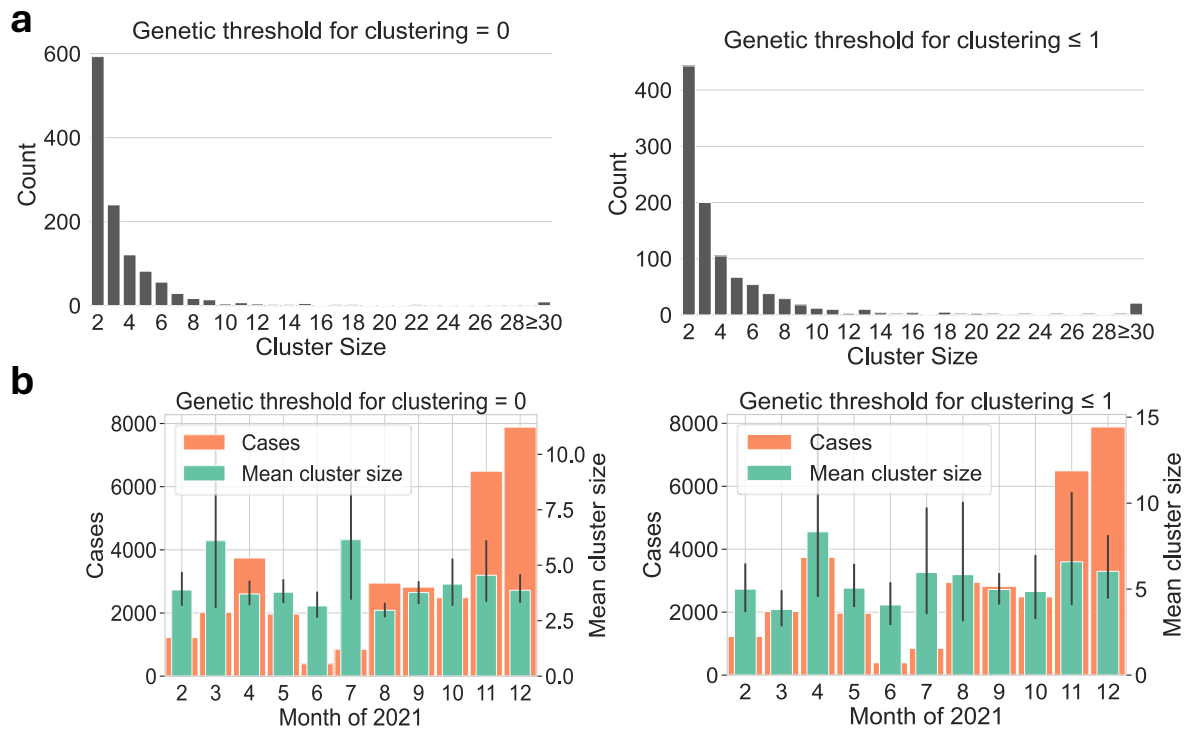

**Supplementary Figure 10: CER genetic cluster sizes.** **a**, Distributions of the size of genetic clusters computed for determining the CER, shown for genetic clustering thresholds $t_g = 0$ (left subpanel) and $t_g = 1$ (right subpanel). **b**, Mean sizes of genetic clusters computed for determining the CER, by month of the year and shown for genetic clustering thresholds $t_g = 0$ (left subpanel) and $t_g = 1$ (right subpanel). Clusters were assigned to months based on the average case registration date of the included cases. Error bars show 95% confidence intervals. Orange bars in the background show the total number of SARS-CoV-2 registered in Düsseldorf in the corresponding month.

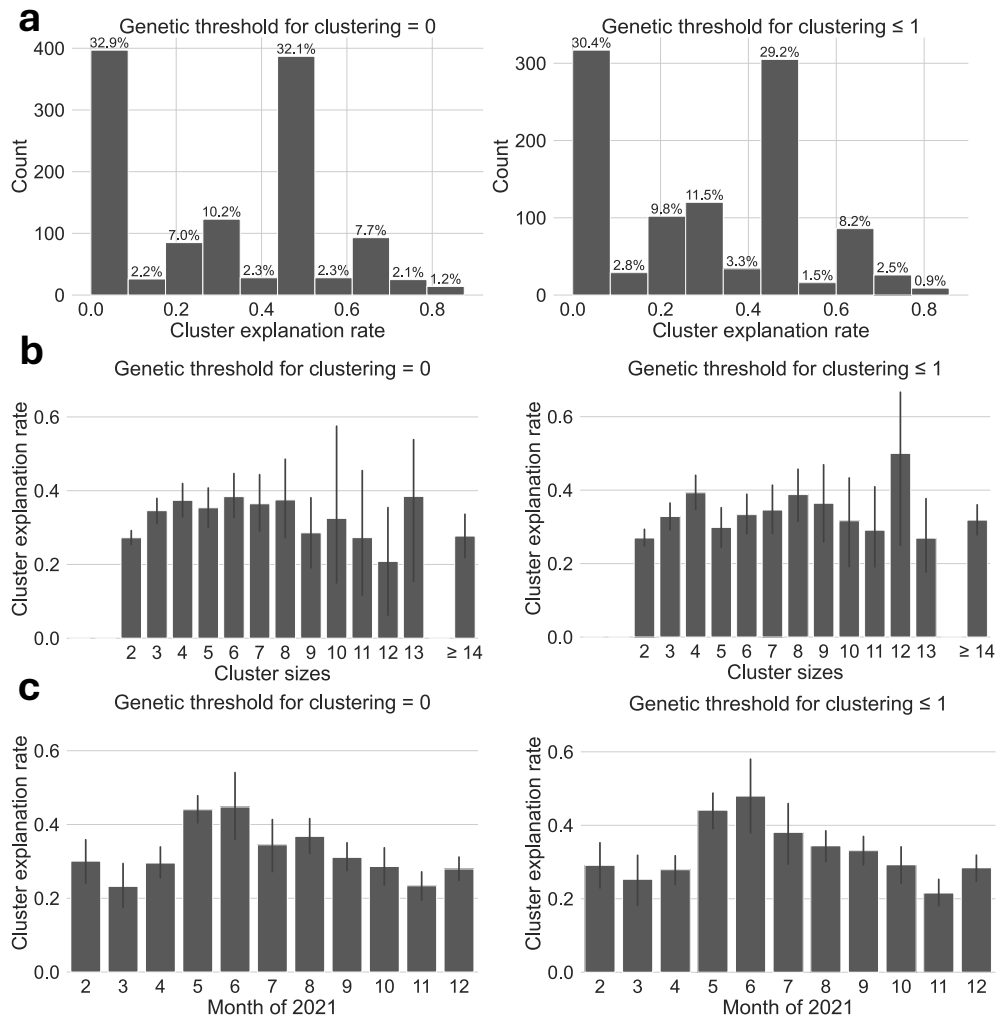

**Supplementary Figure 11: CER parameters and CER over time.** **a**, Distribution of per-cluster CER values, shown for genetic clustering thresholds $t_g = 0$ (left subpanel) and $t_g = 1$ (right subpanel). **b**, CER by cluster size, for genetic clustering thresholds $t_g = 0$ (left subpanel) and $t_g = 1$ (right subpanel). Shown is the average of per-cluster CER values for all clusters of the specified size in cases, with error bars indicating 95% confidence intervals. **c**, CER by month, for genetic clustering thresholds $t_g = 0$ (left subpanel) and $t_g = 1$ (right subpanel). Shown is the average of per-cluster CER values for all clusters assigned to the specified month, with error bars indicating 95% confidence intervals; clusters were assigned to months based on the average case registration date of the included cases. All plots in all panels based on $t_d = -3$ and $t_j = 0$.

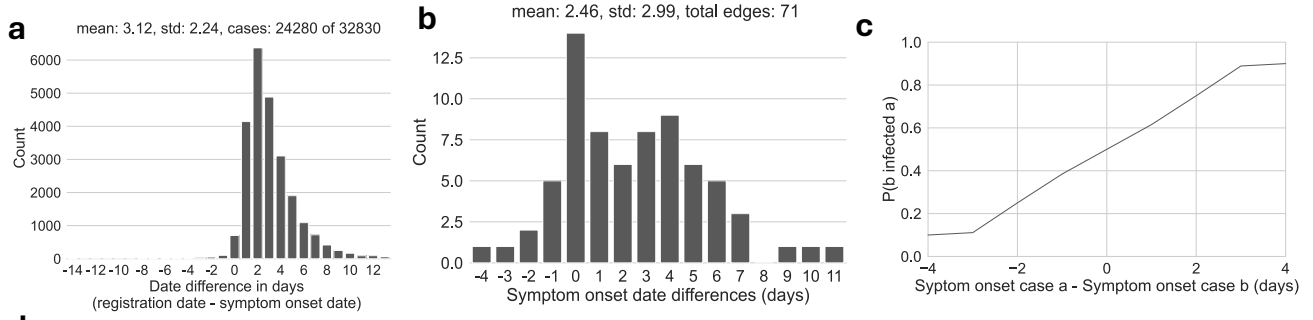

**d**

| | | All edges $t_g = 0$ $t_g = 1$ | | |
| --- | --- | --- | --- | --- |
|  | Maximum count of directed edges | 78,104 | 5,646 | 7,372 |
| $t_d = 0$ | Realized count of directed edges (after directionalization) | 44,371 | 3,406 | 4,439 |
|  | ... of these, expected number of edges with erroneous direction | 9,693 | 949 | 1,218 |
|  | Removed potential directed edges | 33,733 | 2,240 | 2,933 |
|  | ... of these, expected number of edges with correct direction | 4,330 | 364 | 462 |
| $t_d = 3$ | Realized count of directed edges (after directionalization) | 59,690 | 4,665 | 6,024 |
|  | ... of these, expected number of edges with erroneous direction | 21,067 | 1,874 | 2,380 |
|  | Removed potential directed edges | 18,414 | 981 | 1,348 |
|  | ... of these, expected number of edges with correct direction | 382 | 29 | 38 |

**Supplementary Figure 12: Symptom onset and case registration dates.** **a**, Difference between the case registration and symptom onset dates of individual cases. Cases with missing symptom onset data were excluded. **b**, Symptom onset date differences for pairs of sequenced cases with genetic distance $\leq 1$ and connected by edges of type “backward contact tracing”; shown is the difference in symptom onset dates along the assumed directionality of infection as recorded during backward contact tracing (i.e., if case $x$ specified case $y$ as their putative infection source, the plot shows the symptom onset of $x$ minus the symptom onset of $y$; if two cases specified each other as assumed infection sources, two date differences are included). All case pairs comprising cases with missing symptom onset data were excluded. **c**, Transmission directionality probability distribution, based on a simple Bayesian model. Assume there are two cases $a$ and $b$; either $a$ infected $b$ or $b$ infected $a$; define the observed data $x$ as the difference in symptom onset date between $a$ and $b$, i.e. $x = \text{symptom\_onset}(a) - \text{symptom\_onset}(b)$. The probability that $b$ infected $a$ conditional on the observed data is $P(b \rightarrow a|x) = \frac{\Pr(b \rightarrow a) \times P(x|b \rightarrow a)}{\Pr(b \rightarrow a) \times P(x|b \rightarrow a) + \Pr(a \rightarrow b) \times P(x|a \rightarrow b)}$. Assuming uniform priors, we obtain $P(b \rightarrow a|x) = \frac{P(x|b \rightarrow a)}{P(x|b \rightarrow a) + P(x|a \rightarrow b)}$. We empirically estimated a probability distribution $P_{\text{Backward}}$ on the difference in symptom onset dates between the infected case and the infecting case observed in backward contact tracing; specifically, $P_{\text{Backward}}(d) = \frac{|(c_1, c_2) \in E'_B : \text{symptom\_onset}(c_2) - \text{symptom\_onset}(c_1) = d|}{|E'_B|}$, where $E'_B$ is the set of edges of type “backward contact tracing” in the Integrated Case Graph. We define $P(x|b \rightarrow a) = P_{\text{Backward}}(x)$. With respect to $P(x|a \rightarrow b)$, we note that $x$ is, conditional on the assumption “$a \rightarrow b$” that $a$ infected $b$, the difference in symptom onset dates between the infecting case and the infected case, i.e., the opposite of the required parameterization for $P_{\text{Backward}}(d)$. To obtain the difference in symptom onset dates between the infected case and the infected case under the assumption that $a$ infected $b$, $x$ can be multiplied by $-1$; we thus obtain $P(x|b \rightarrow a) = P_{\text{Backward}}(-x)$. This gives $P(b \rightarrow a|x) = \frac{P_{\text{Backward}}(x)}{P_{\text{Backward}}(x) + P_{\text{Backward}}(-x)}$. **d**, Expected numbers of directed edges with correct and incorrect directionality, stratified by whether they were retained or removed during graph directionalization (with parameter $t_d$ set to 0 or 3) and shown separately for different thresholds on the genetic distance between the connected cases (no evaluation of genetic distances / $t_g = 0$ / $t_g = 1$). The probability that an edge $(b, a)$ has the correct directionality was given by $P(b \rightarrow a|x)$ (see panel C.), with $P(b \rightarrow a|x)$ set to 0 for $x \leq -5$ and to 1 for $x \geq 5$.

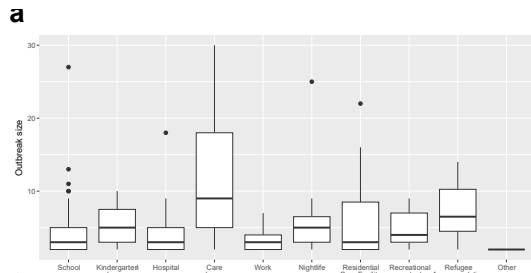

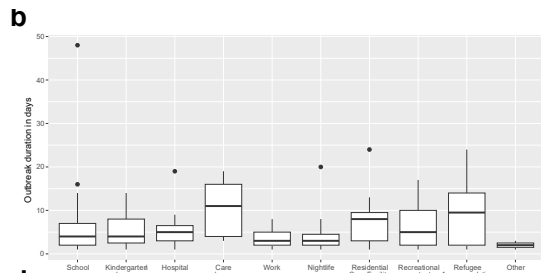

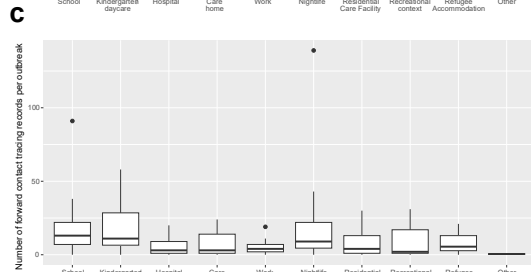

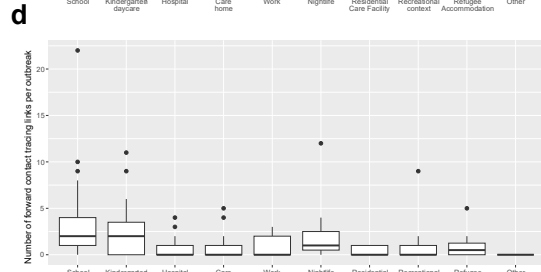

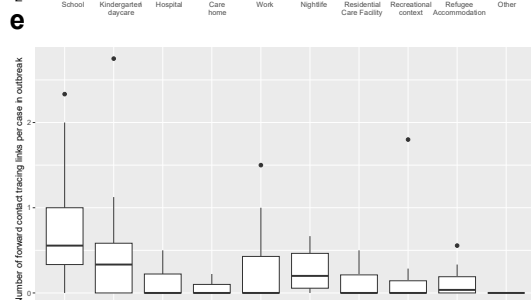

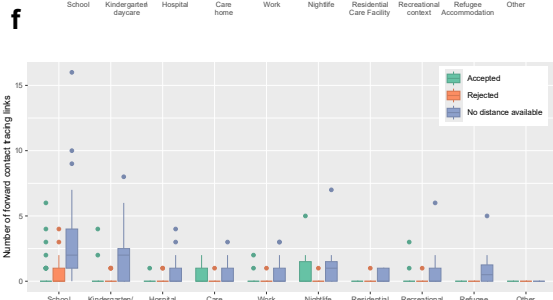

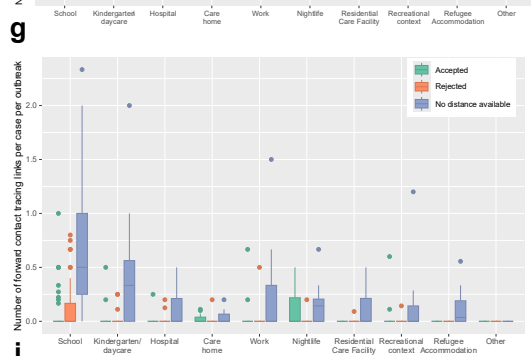

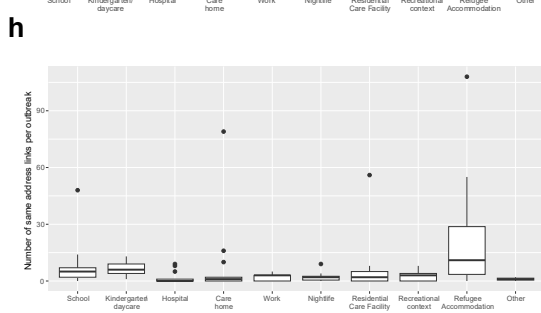

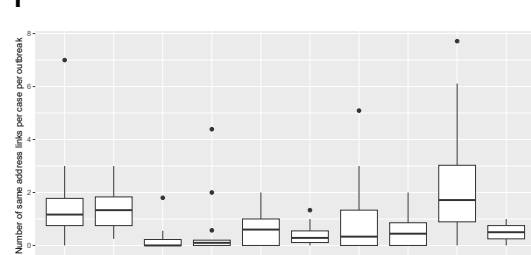

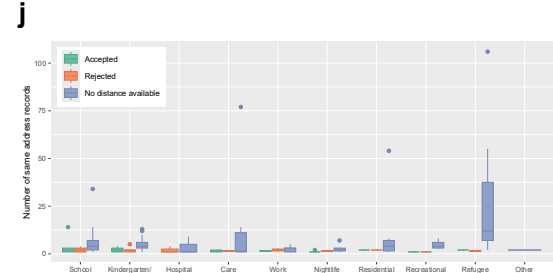

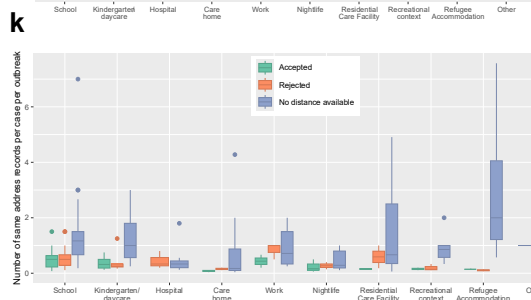

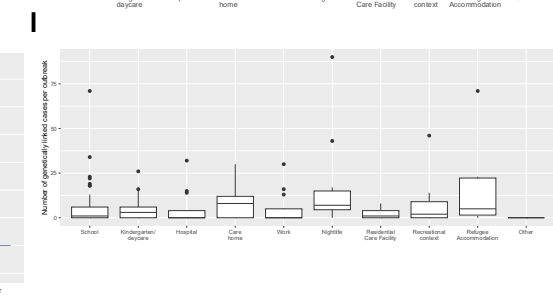

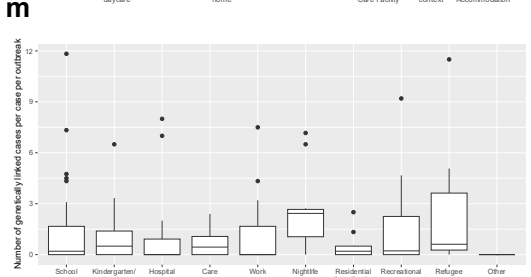

**Supplementary Figure 13: Integrated outbreak analysis, additional metrics.** **a**, Outbreak sizes, stratified by outbreak type; outbreak size was defined as the number of cases assigned to the outbreak by Düsseldorf Health Authority. **b**, Outbreak durations, stratified by outbreak type; the duration of an outbreak was defined as the time in days from the earliest to the latest case registration date of the cases assigned to the outbreak. **c**, Forward contact tracing records per outbreak (defined as the cumulative number of forward contact tracing records of all outbreak-assigned cases, independent of whether the contacts were mapped onto other cases or not), stratified by outbreak type. **d**, “Forward contact tracing”-type edges connecting outbreak cases to non-outbreak cases in the Integrated Case Graph, per outbreak and stratified by outbreak type. **e**, Like panel D but normalized by outbreak size. **f**, “Forward contact tracing”-type edges connecting outbreak cases to non-outbreak cases in the Integrated Case Graph, per outbreak and stratified by outbreak type and by whether the connected non-outbreak case had a genetic distance of $\leq 1$ to the outbreak (“accepted”), whether it had a genetic distance $\geq 2$ (“rejected”), or whether a genetic distance could not be computed (“No distance available”). The genetic distance between a linked non-outbreak case and an outbreak was defined as the minimum genetic distance between the linked case and any case of the corresponding outbreak, and a distance could be computed if the outbreak contained at least one sequenced case and if the linked case was sequenced. **g**, Like panel F but normalized by outbreak size. **h**, “sameAddress”-type edges connecting outbreak cases to non-outbreak cases in the Integrated Case Graph, per outbreak and stratified by outbreak type. **i**, Like panel H but normalized by outbreak size. **j**, Like panel F but for “sameAddress”-type edges. **k**, Like panel G but for “sameAddress”-type edges. **l**, Number of genetically linked community cases, per outbreak and by outbreak type; genetically linked community cases were defined as cases not linked to the analyzed outbreak that had a genetic distance of 0 to at least one case of the corresponding outbreak. **m**, Like panel L but normalized by outbreak size.

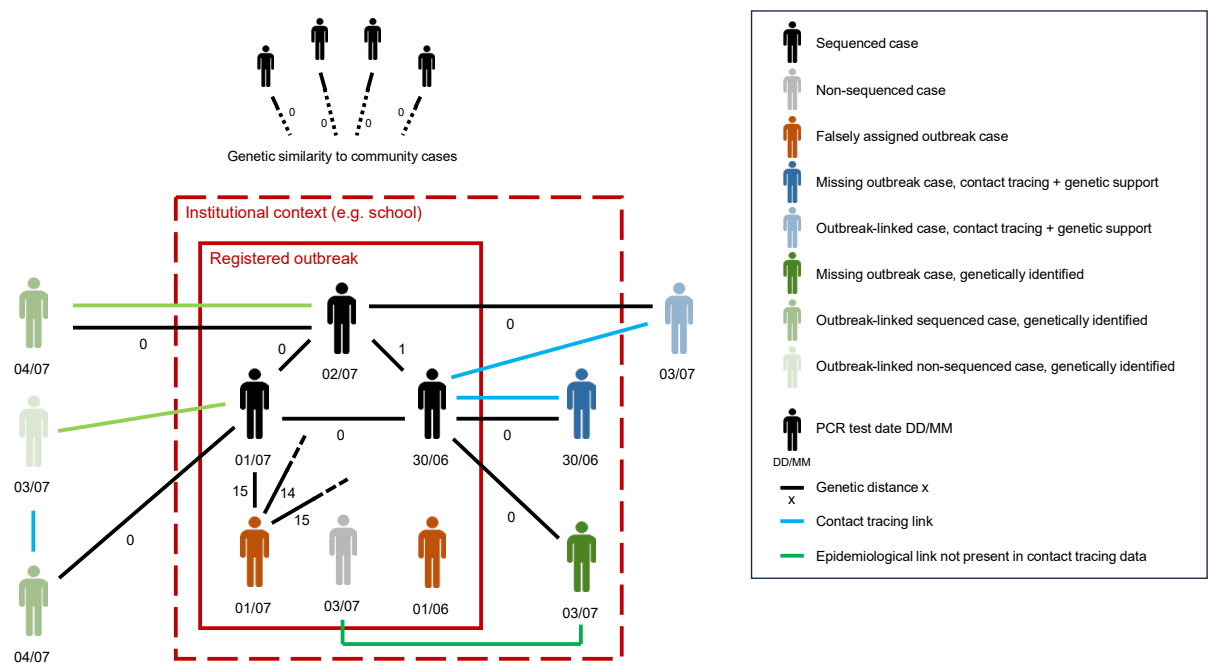

**Supplementary Figure 14: Manual investigation of outbreaks.** Shown is an outbreak comprising 6 cases, 4 of which are sequenced and 2 of which are not. 2 cases were likely erroneously assigned to the outbreak (shaded in red), as indicated by PCR test dates in one case and genetic distances to other outbreak cases in the other case. Two cases are linked to the outbreak via contact tracing; one of these is, based on belonging to the same institutional context as the other outbreak cases, classified as a “missing” outbreak case (shaded in dark blue), and the other case is classified as an “outbreak-linked” case (shaded in light blue); the association between the cases and the outbreaks is supported by sequencing data in both instances. Three additional cases not assigned to the outbreak exhibit a genetic distance of 0 to at least one outbreak case. The first of the three cases is, based on belonging to the same institutional context as the other outbreak cases, classified as a “missing” outbreak case (shaded in dark green). For the second of the three cases, an epidemiological connection to the outbreak that is not present in the contact tracing data is discovered and the case is classified as outbreak-linked based on the discovered connection (shaded in medium-intensity green). Finally, for the third case (shaded in medium-intensity green), an epidemiological connection to the outbreak that involves a non-sequenced additional case (shaded light green) is discovered. The third case and the non-sequenced additional case are therefore classified as outbreak-linked; in the case of the non-sequenced additional case, its classification as outbreak-linked is based on the fact that it represents the only detected connection that can explain the genetic identity between the third sample and the outbreak.

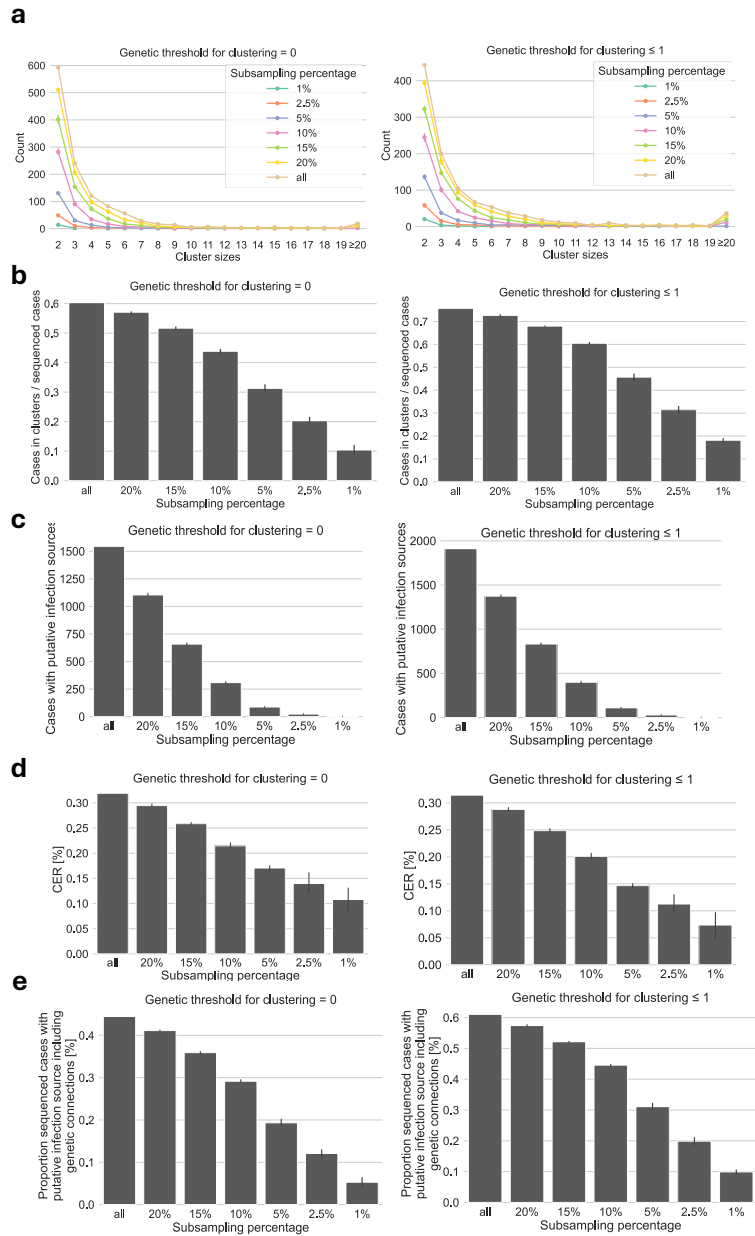

**Supplementary Figure 15: Subsampling of proportion of sequenced cases for the CER.** **a**, Distribution of cluster sizes for different proportions of sequenced cases (x-axis) and $t_g = 0$ (left-hand panel) and $t_g = 1$ (right-hand panel). **b**, Absolute number of cases with a putative transmission source in the computed hypothetical transmission chains, for different proportions of sequenced cases (x-axis) and based on clusters computed for $t_g = 0$ (left-hand panel) and $t_g = 1$ (right-hand panel). **c**, Cluster Explanation Rate (CER), for different proportions of sequenced cases (x-axis) and for $t_g = 0$ (left-hand panel) and $t_g = 1$ (right-hand panel). **d**, Proportion of cases with a putative infection source based on the assumption that all genetic links correspond to an epidemiological link, for different proportions of sequenced cases (x-axis) and $t_g = 0$ (left-hand panel) and $t_g = 1$ (right-hand panel).

Sequencing rate 1%

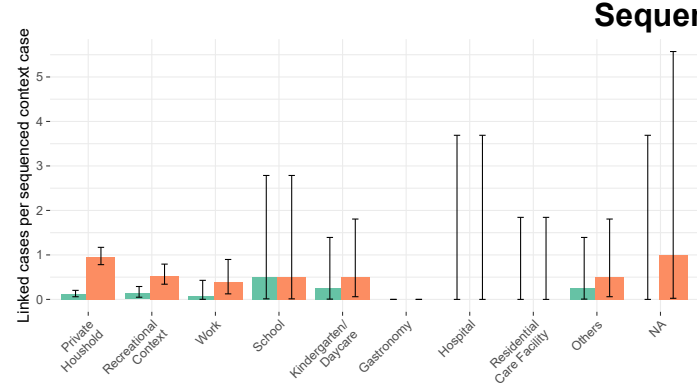

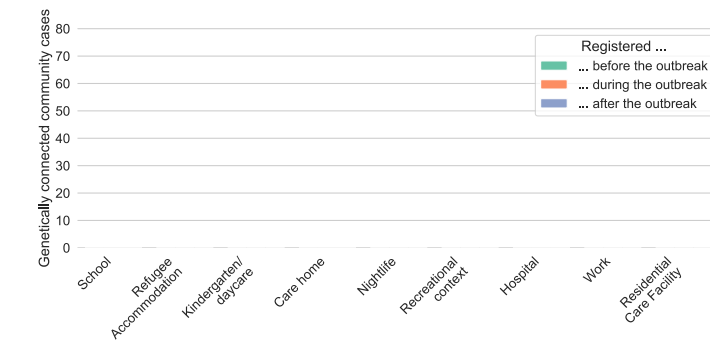

Sequencing rate 2.5%

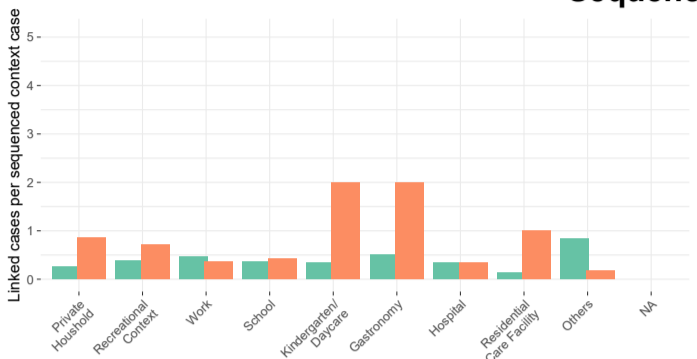

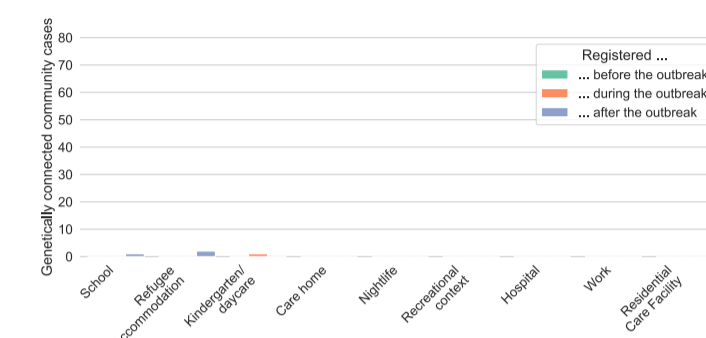

Sequencing rate 5%

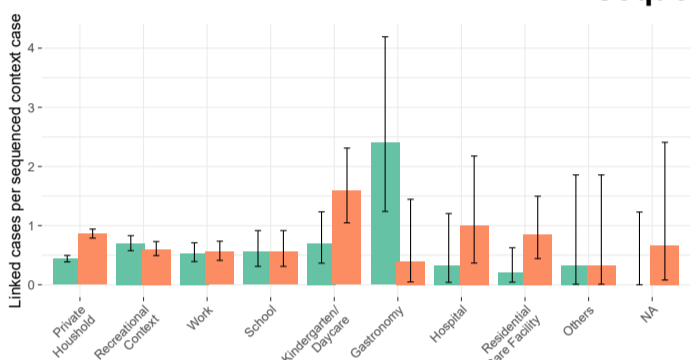

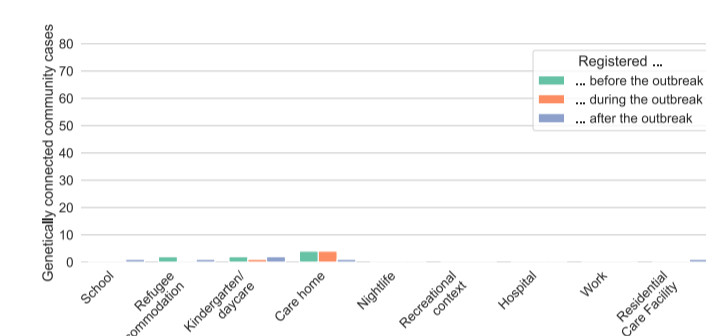

Sequencing rate 10%

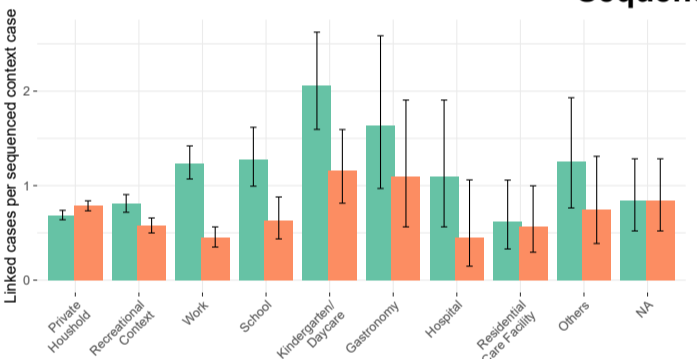

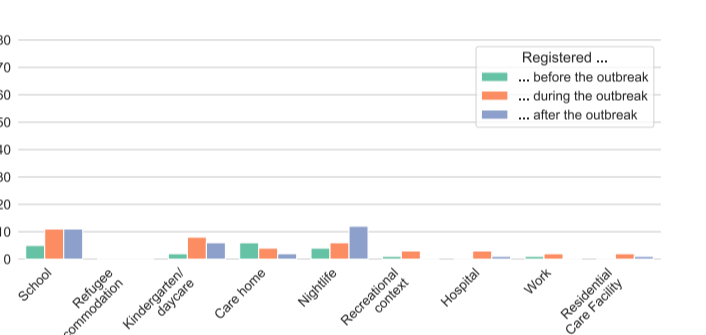

Sequencing rate 15%

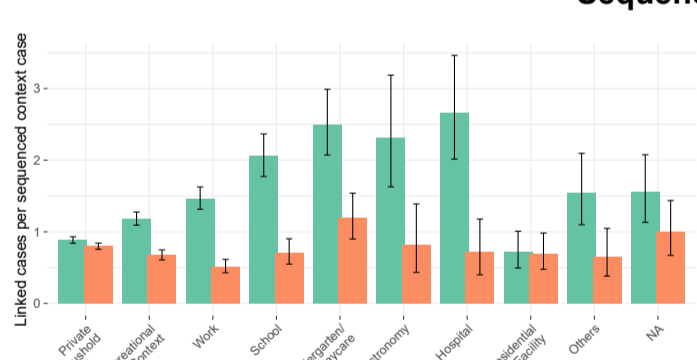

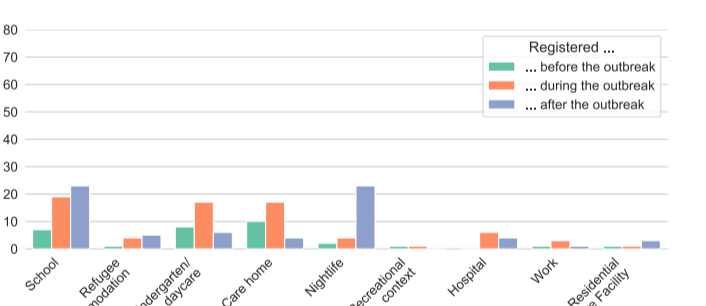

Sequencing rate 20%

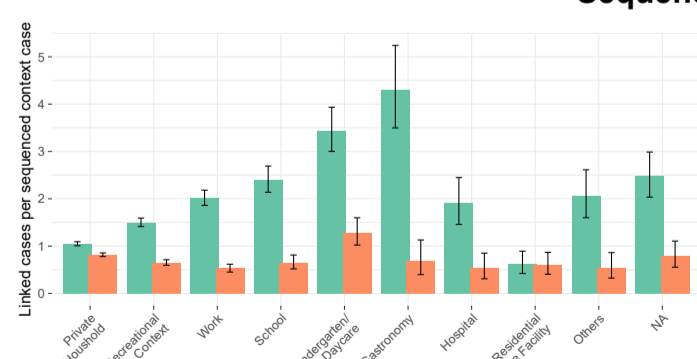

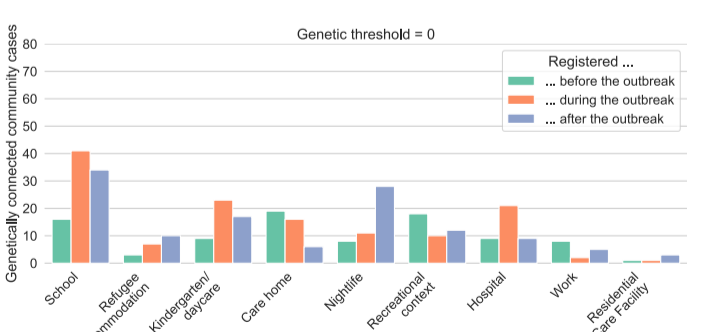

Sequencing rate 24.5%

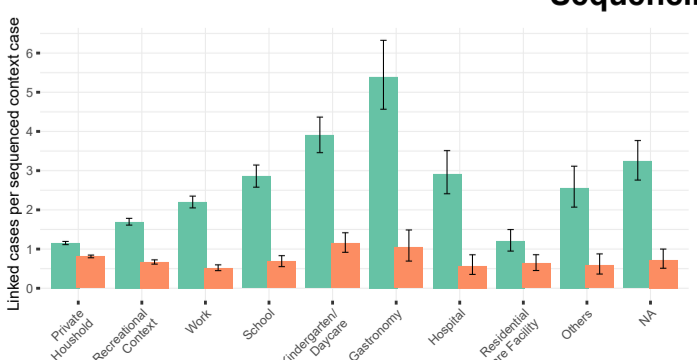

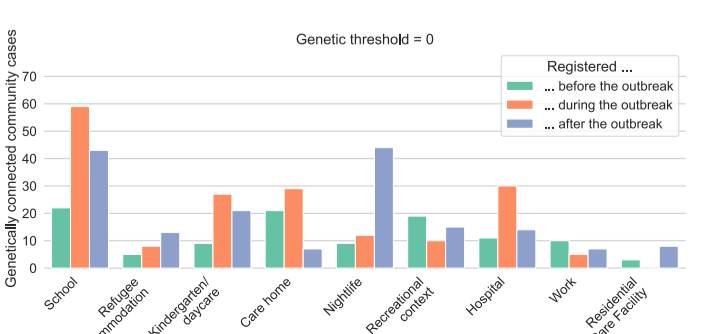

**Supplementary Figure 16: Subsampling of proportion of sequenced cases for characterization of large-scale transmission patterns.** Left-hand panels: Average number of genetically linked cases (genetic distance 0 and difference in registration in dates $\leq 14$ days) compared to the average number of sequenced cases linked via classical contact tracing for cases with contacts of the given category at different proportions of sequenced cases. Right-hand panels: Cases genetically linked to outbreaks and stratified by outbreak type and relative case registration date at different proportions of sequenced cases; 8 outbreaks that were associated with the Delta and Omicron variants and that overlapped, with their surrounding 2-week windows, with the expansion phases of these variants were not included.

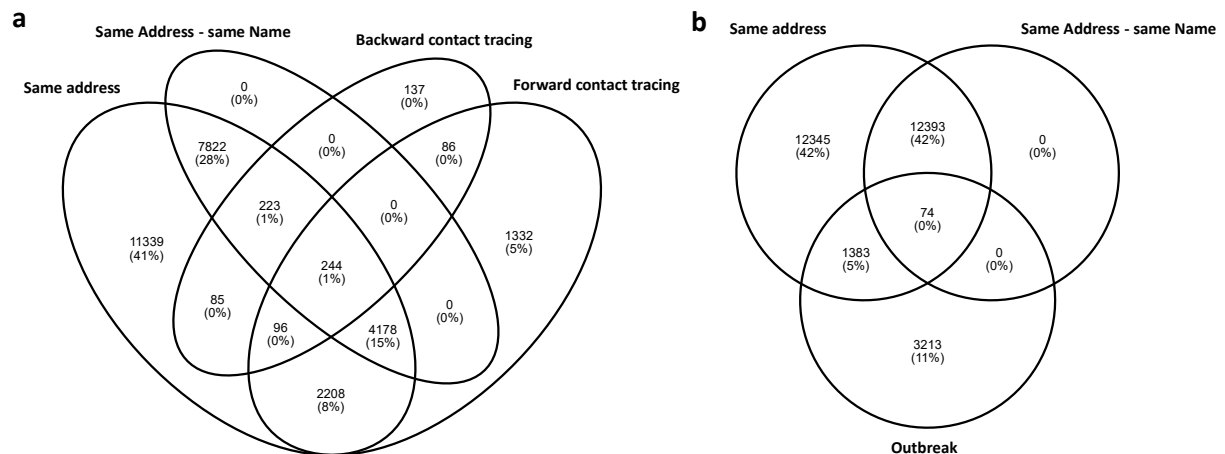

**Supplementary Figure 17: Overlap of “sameAddress”-type edges in the Integrated Case Graph with other edge types. a, Overlap with contact tracing edges.** Shown is the overlap between edges of type “sameAddress”, “forward contact tracing”, and “backward contact tracing” in the Integrated Case Graph. **b, Overlap with “outbreak”-type edges.** For both panels, two edges of different types were counted as overlapping if they connected the same two cases. The label “Same Address - same Name” refers to edges of type “sameAddress-sameName”; the label “Same address”, to edges of type “sameAddress-differentName”.

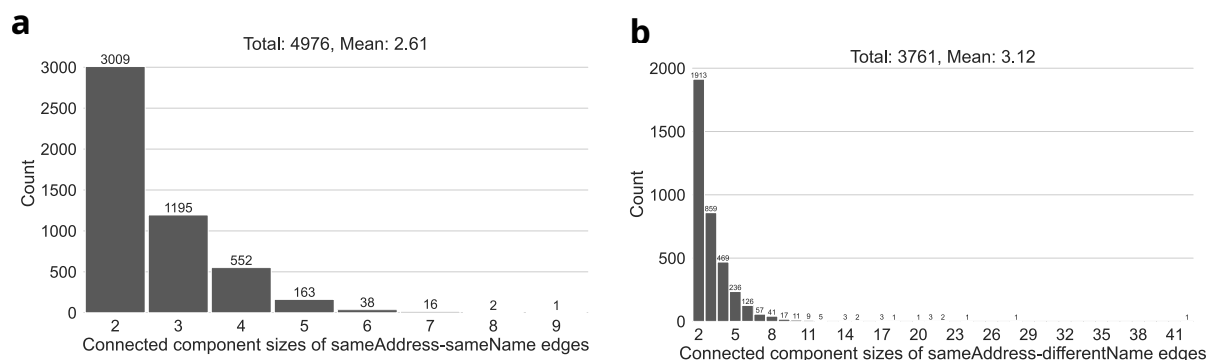

**Supplementary Figure 18: Size of “sameAddress”-type connected components. a, Distribution of the size of connected components in the Integrated Case Graph when only considering edges of type “sameAddress-sameName”.** **b, Distribution of the size of connected components in the Integrated Case Graph when only considering edges of type “sameAddress-differentName”.**

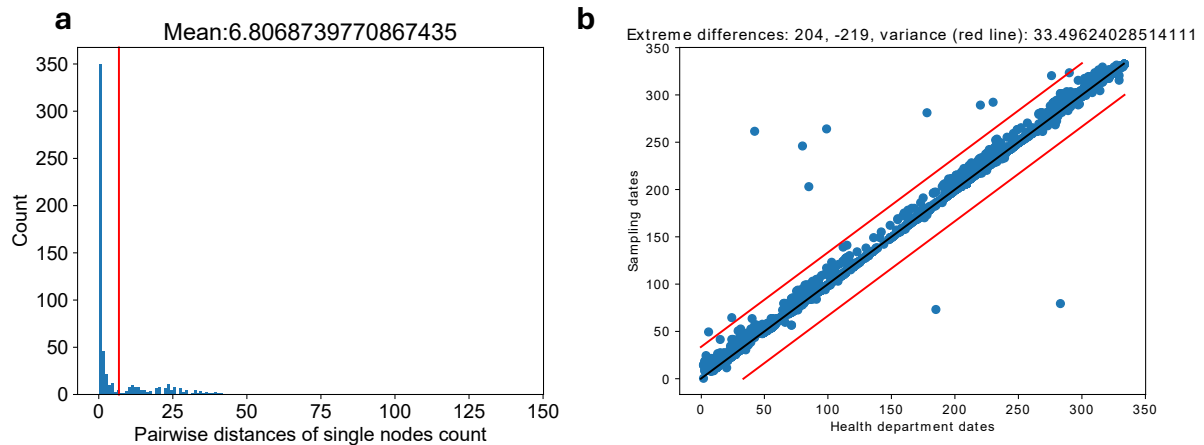

**Supplementary Figure 19: SARS-CoV-2 sample selection and assignment.** **a**, Pairwise genetic distances between SARS-CoV-2 genomes assigned to the same case, prior to the selection of at most one viral genome sequencing record for all cases. The red line shows the mean of the distribution. **b**, Scatterplot comparing, for each generated SARS-CoV-2 genome after quality control and before outlier removal, the sampling date of the diagnostic swab used for SARS-CoV-2 sequencing (Y axis) to the Düsseldorf Health Authority registration date of the case that the viral genome sequencing record was assigned to (see Methods). The diagonal (difference = 0) is shown as a black line; red lines show the variance. Before proceeding to the selection of at most one unique viral genome per case, SARS-CoV-2 sequencing records which exhibited a difference between case registration and sample collection dates of $< -21$ or $> 7$ days were removed (see Methods).

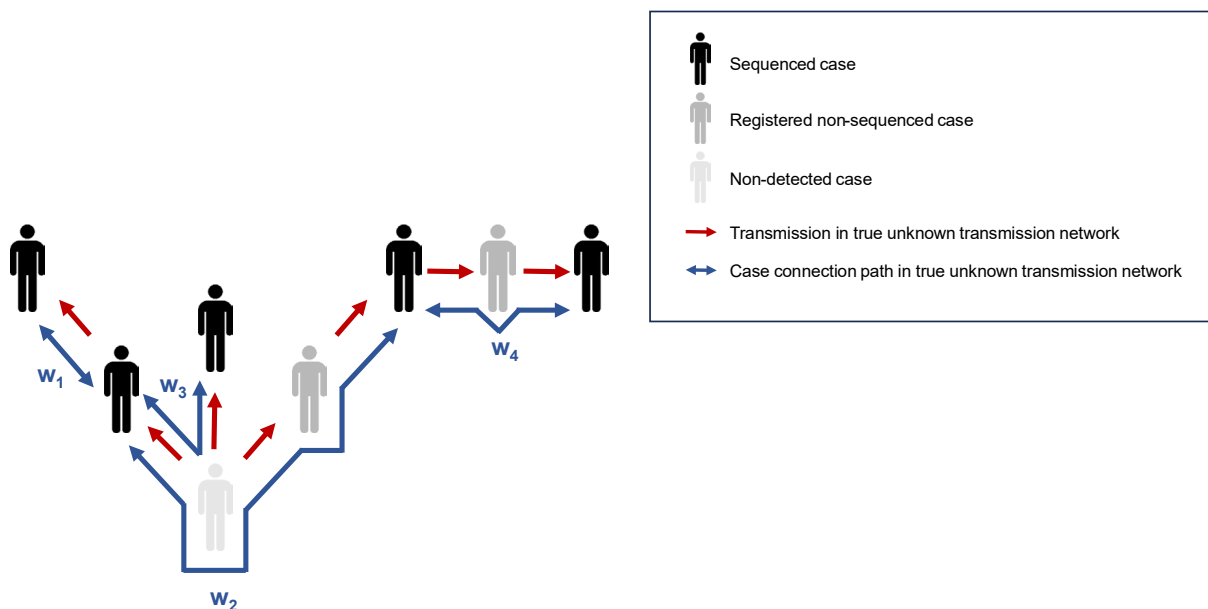

**Supplementary Figure 20: SARS-CoV-2 example transmission network.** Transmission network of 8 cases, comprising 5 sequenced cases, 2 cases which were registered but not sequenced, and 1 undetected case. Red arrows show transmissions in the (generally unknown) complete transmission network; blue connections show 4 case connection paths ($w_1 - w_4$) between sequenced cases in the complete network, which connect the sequence pairs most closely related in terms of the unknown full network. A hypothetical transmission chain for the 5 sequenced cases may imply a putative infection source for up to 80% (4 / 5) of cases if “jump edges” (see Methods) are included and if case connection path $w_4$ can be found in the known epidemiological connections.

**a**

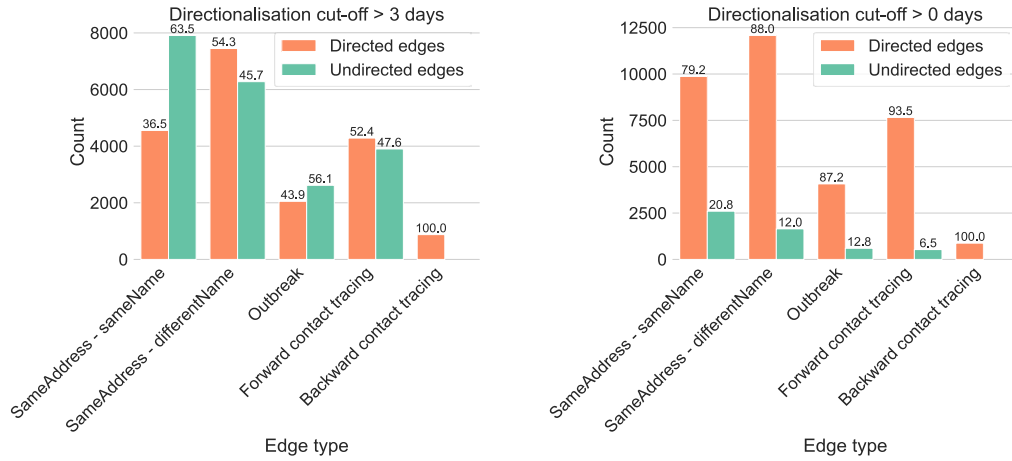

**b**

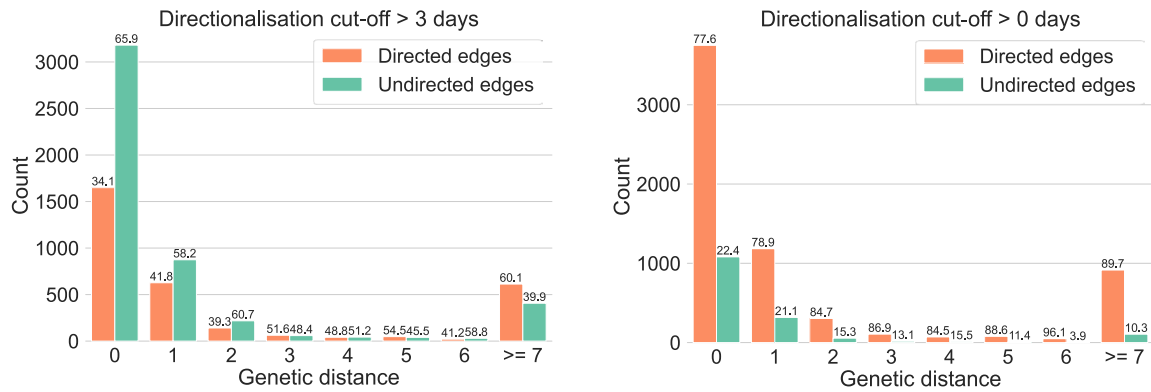

**Supplementary Figure 21: Graph directionalization parameter $t_d$.** The plots show, for two different values (0 and 3) of the “maximum symptom onset difference” directionalization parameter $t_d$ (“Directionalization cut-off”), the number of edges in the Integrated Case Graph that were converted into one (“Directed edges”) or two directed edges (“Undirected edges”) in the directionalized version of the Integrated Case Graph, stratified by edge type (panel **a**) or by the genetic distance between the connected cases (panel **b**; only for case pairs consisting of two sequenced cases). Directionalization captures the concept of directionality of transmission and ensures that the symptom onset dates of cases connected in hypothetical transmission chains are compatible with the directionality of hypothetical transmission. In the process of directionalization, undirected edges of the form $(c_1, c_2)$ are converted into either a single directed edge of the form $(c_1, c_2)$ or $(c_2, c_1)$, or into a pair of directed edges of the form $(c_1, c_2)$ and $(c_2, c_1)$. Setting $t_d = 0$ implies that all connections (with the exception of links induced by backward contact tracing, see below) between two cases in the Integrated Case Graph become “directed” (i.e., are represented by a single directed edge) whenever there is any difference in symptom onset dates, and that all considered hypothetical transmissions always follow the direction of increasing symptom onset. The directionalization of edges of type “backward contact tracing” is carried out based on the individual cases’ assessment of their assumed infection source; a “backward contact tracing” edge from case $c_2$ to case $c_1$ in the directionalized graph is created whenever $c_1$ specified $c_2$ as their assumed infection source, independent of symptom onset dates.
