## Supplementary Note 1 for "Genomics-enhanced contact tracing enabled the characterization of SARS-CoV-2 transmission chains and infection contexts in the general population during community transmission"

### Supplementary Note: Analysis details of selected outbreaks

#### COV-DÜS-SCHULE-2021-0078

COV-DÜS-SCHULE-2021-0078 was a school outbreak registered during domination of the „Alpha“ variant and comprised 2 registered cases, both of whom were sequenced and who were pupils in the same class. Scanning for cases genetically related to the outbreak uncovered 2 missing outbreak cases (Cases 3 and 4); one of the two cases attended the same class as the outbreak cases, while the other one attended a different class at the same school, implying that COV-DÜS-SCHULE-2021-0078 was, in contrast to the classification of the outbreak by Düsseldorf Health Authority as a single-class outbreak, a multi-class outbreak. Genetic relatedness of the two missing cases to the outbreak was established, in one case, directly (Case 4), and, in the other case, indirectly (Case 3); Case 4 was sequenced and found to be genetically identical (genetic distance $\leq 1$) to the two sequenced outbreak cases; Case 3 was not itself sequenced but represented the only identified epidemiological link between a community case (Case 5; parent of the missing case) exhibiting genetic similarity to the outbreak (genetic distance $\leq 1$). Overall, three community cases (parents of pupils) were found to be linked (epidemiologically and genetically supported) to the outbreak; one of these (Case 5) was only identified through an analysis of genetic similarities. In two of the three cases, the linked community cases were likely infected by outbreak samples; in the third case, the linked community case (Case 5) may have infected the outbreak case based on an analysis of symptom onset dates and may thus represent the beginning of the infection chain leading to outbreak COV-DÜS-SCHULE-2021-0078.

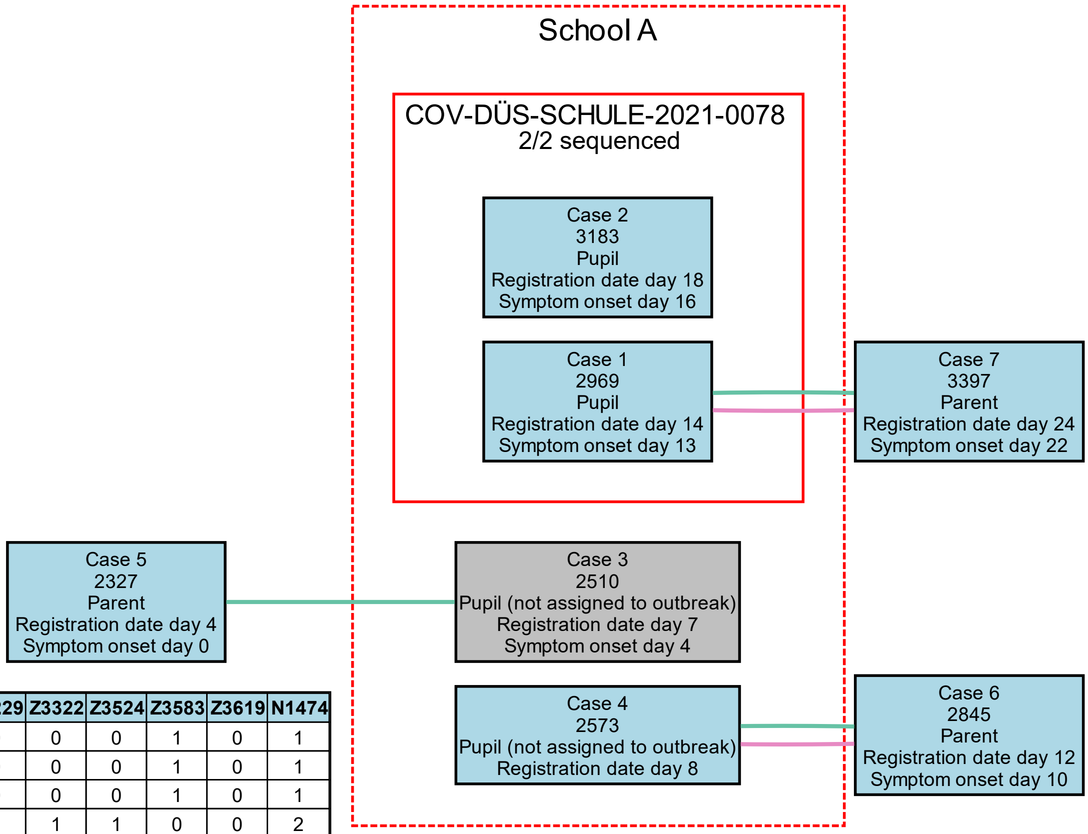


| | Z3229 | Z3322 | Z3524 | Z3583 | Z3619 | N1474 |
|---|---|---|---|---|---|---|
| **Z3229** | 0 | 0 | 0 | 1 | 0 | 1 |
| **Z3322** | 0 | 0 | 0 | 1 | 0 | 1 |
| **Z3524** | 0 | 0 | 0 | 1 | 0 | 1 |
| **Z3583** | 1 | 1 | 1 | 0 | 0 | 2 |
| **Z3619** | 0 | 0 | 0 | 0 | 0 | 1 |
| **N1474** | 1 | 1 | 1 | 2 | 1 | 0 |

**Genetic clusters**

1

Not sequenced

**Edge color legend**

Forward Contact Tracing

Backward Contact Tracing

sameAddress-sameName

sameAddress-differentName

Manual analysis

#### CVD-DÜS-SCHULE-2021-0107 and CVD-DÜS-SCHULE-2021-0108

CVD-DÜS-SCHULE-2021-0107 and CVD-DÜS-SCHULE-2021-0108 were two separately registered multi-class outbreaks at the same school, comprising a total of 4 (2 of which were sequenced) and 26 (6 of which were sequenced) cases, respectively. The outbreaks fell into the period of domination by the "Delta" variant of SARS-CoV-2. Analysis of genetic similarities showed that the two outbreaks were genetically related (genetic distance $\leq 1$) and that they therefore should have been classified as a single outbreak. Searching for genetically related non-outbreak cases identified two additional cases (pupils attending the same school) that should likely have been assigned to the outbreak. For the first case (Case 24), this assignment was based on genetic similarity between the missing case and sequenced outbreak samples. For the second case (Case 23), the classification as a missing outbreak was based on the fact that the case represented the only identified epidemiological link between the outbreak and another genetically related community sample (Case 38; genetic distance $\leq 1$); of note, however, Case 38 reported symptom onset two days before Case 23. In total, the outbreak exhibited 14 sequencing-confirmed links to community cases that were also epidemiologically supported; two of these (Cases 32 and 38) were only identified through an analysis of genetic similarities. The 14 linked community samples included 7 parents of outbreak cases and 6 pupils attending other schools; none of these, however, were assigned to other outbreaks.

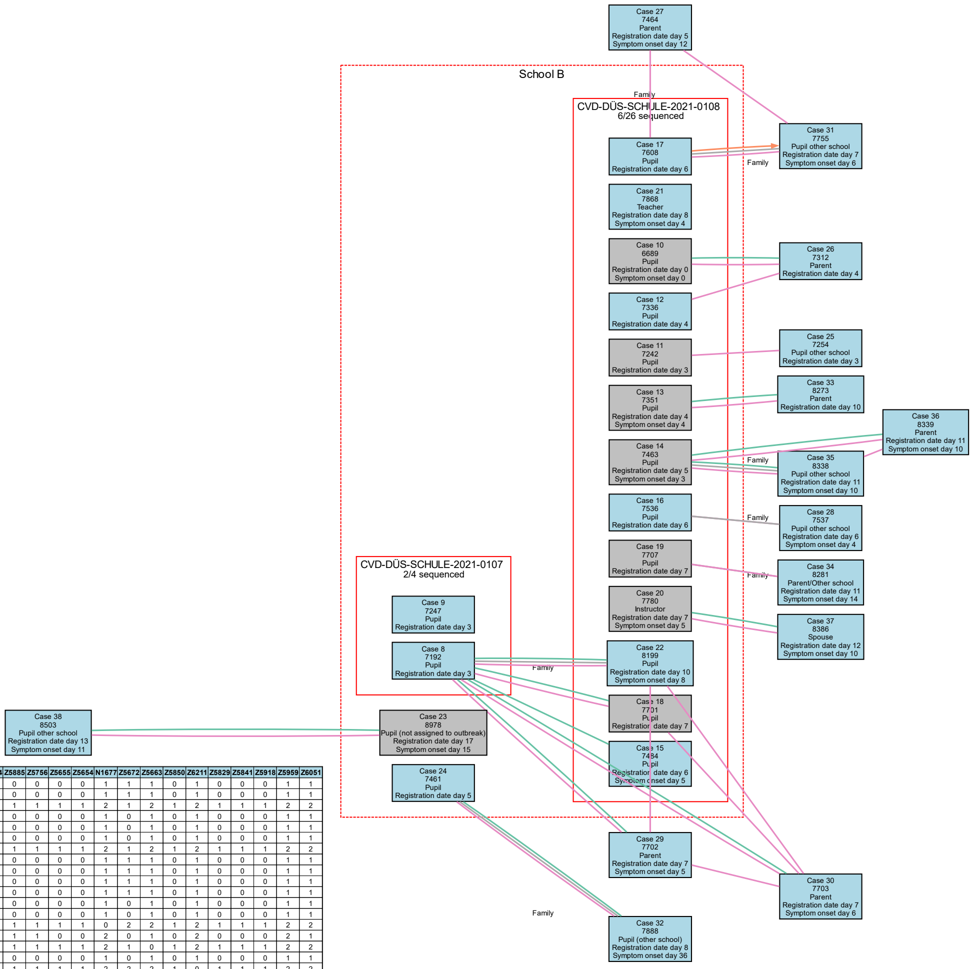


| | Z5438 | Z5893 | Z5424 | Z5486 | Z5487 | Z6248 | Z6052 | Z5605 | Z5884 | Z5885 | Z5756 | Z5655 | Z5654 | N1677 | Z5672 | Z5663 | Z5850 | Z6211 | Z5829 | Z5841 | Z5918 | Z5959 | Z6051 |
|---|---|---|---|---|---|---|---|---|---|---|---|---|---|---|---|---|---|---|---|---|---|---|---|
| **Z5438** | 0 | 0 | 1 | 0 | 0 | 0 | 1 | 0 | 0 | 0 | 0 | 0 | 0 | 1 | 1 | 1 | 0 | 1 | 0 | 0 | 0 | 1 | 1 |
| **Z5893** | 0 | 0 | 1 | 0 | 0 | 0 | 1 | 0 | 0 | 0 | 0 | 0 | 0 | 1 | 1 | 1 | 0 | 1 | 0 | 0 | 0 | 1 | 1 |
| **Z5424** | 1 | 1 | 0 | 1 | 1 | 1 | 2 | 1 | 1 | 1 | 1 | 1 | 1 | 2 | 1 | 2 | 1 | 2 | 1 | 1 | 1 | 2 | 2 |
| **Z5486** | 0 | 0 | 1 | 0 | 0 | 0 | 1 | 0 | 0 | 0 | 0 | 0 | 0 | 1 | 0 | 1 | 0 | 1 | 0 | 0 | 0 | 1 | 1 |
| **Z5487** | 0 | 0 | 1 | 0 | 0 | 0 | 1 | 0 | 0 | 0 | 0 | 0 | 0 | 1 | 0 | 1 | 0 | 1 | 0 | 0 | 0 | 1 | 1 |
| **Z6248** | 0 | 0 | 1 | 0 | 0 | 0 | 1 | 0 | 0 | 0 | 0 | 0 | 0 | 1 | 0 | 1 | 0 | 1 | 0 | 0 | 0 | 1 | 1 |
| **Z6052** | 1 | 1 | 2 | 1 | 1 | 1 | 0 | 1 | 1 | 1 | 1 | 1 | 1 | 2 | 1 | 2 | 1 | 2 | 1 | 1 | 1 | 2 | 2 |
| **Z5605** | 0 | 0 | 1 | 0 | 0 | 0 | 1 | 0 | 0 | 0 | 0 | 0 | 0 | 1 | 1 | 1 | 0 | 1 | 0 | 0 | 0 | 1 | 1 |
| **Z5884** | 0 | 0 | 1 | 0 | 0 | 0 | 1 | 0 | 0 | 0 | 0 | 0 | 0 | 1 | 1 | 1 | 0 | 1 | 0 | 0 | 0 | 1 | 1 |
| **Z5885** | 0 | 0 | 1 | 0 | 0 | 0 | 1 | 0 | 0 | 0 | 0 | 0 | 0 | 1 | 1 | 1 | 0 | 1 | 0 | 0 | 0 | 1 | 1 |
| **Z5756** | 0 | 0 | 1 | 0 | 0 | 0 | 1 | 0 | 0 | 0 | 0 | 0 | 0 | 1 | 1 | 1 | 0 | 1 | 0 | 0 | 0 | 1 | 1 |
| **Z5655** | 0 | 0 | 1 | 0 | 0 | 0 | 1 | 0 | 0 | 0 | 0 | 0 | 0 | 1 | 0 | 1 | 0 | 1 | 0 | 0 | 0 | 1 | 1 |
| **Z5654** | 0 | 0 | 1 | 0 | 0 | 0 | 1 | 0 | 0 | 0 | 0 | 0 | 0 | 1 | 0 | 1 | 0 | 1 | 0 | 0 | 0 | 1 | 1 |
| **N1677** | 1 | 1 | 2 | 1 | 1 | 1 | 2 | 1 | 1 | 1 | 1 | 1 | 1 | 0 | 2 | 2 | 1 | 2 | 1 | 1 | 1 | 2 | 2 |
| **Z5672** | 1 | 1 | 1 | 0 | 0 | 0 | 1 | 1 | 1 | 1 | 1 | 0 | 0 | 2 | 0 | 1 | 0 | 2 | 0 | 0 | 0 | 2 | 1 |
| **Z5663** | 1 | 1 | 2 | 1 | 1 | 1 | 2 | 1 | 1 | 1 | 1 | 1 | 1 | 2 | 1 | 0 | 1 | 2 | 1 | 1 | 1 | 2 | 2 |
| **Z5850** | 0 | 0 | 1 | 0 | 0 | 0 | 1 | 0 | 0 | 0 | 0 | 0 | 0 | 1 | 0 | 1 | 0 | 1 | 0 | 0 | 0 | 1 | 1 |
| **Z6211** | 1 | 1 | 2 | 1 | 1 | 1 | 2 | 1 | 1 | 1 | 1 | 1 | 1 | 2 | 2 | 2 | 1 | 0 | 1 | 1 | 1 | 2 | 2 |
| **Z5829** | 0 | 0 | 1 | 0 | 0 | 0 | 1 | 0 | 0 | 0 | 0 | 0 | 0 | 1 | 0 | 1 | 0 | 1 | 0 | 0 | 0 | 1 | 1 |
| **Z5841** | 0 | 0 | 1 | 0 | 0 | 0 | 1 | 0 | 0 | 0 | 0 | 0 | 0 | 1 | 0 | 1 | 0 | 1 | 0 | 0 | 0 | 1 | 1 |
| **Z5918** | 0 | 0 | 1 | 0 | 0 | 0 | 1 | 0 | 0 | 0 | 0 | 0 | 0 | 1 | 0 | 1 | 0 | 1 | 0 | 0 | 0 | 1 | 1 |
| **Z5959** | 1 | 1 | 2 | 1 | 1 | 1 | 2 | 1 | 1 | 1 | 1 | 1 | 1 | 2 | 2 | 2 | 1 | 2 | 1 | 1 | 1 | 0 | 2 |
| **Z6051** | 1 | 1 | 2 | 1 | 1 | 1 | 2 | 1 | 1 | 1 | 1 | 1 | 1 | 2 | 1 | 2 | 1 | 2 | 1 | 1 | 1 | 2 | 0 |

**Genetic clusters**

1

Not sequenced

**Edge color legend**

Forward Contact Tracing

Backward Contact Tracing

sameAddress-sameName

sameAddress-differentName

Manual analysis

#### CVD-DÜS-SCHULE-2021-0169 and CVD-DÜS-KITA-2021-0164

CVD-DÜS-SCHULE-2021-0169 and CVD-DÜS-KITA-2021-0164 were two outbreaks at a school and at a preschool ("KiTa" in the German educational system), comprising a total of 3 (3 of which were sequenced) and 5 (1 of which was sequenced) cases, respectively. The outbreaks fell into the period of domination by the "Delta" variant of SARS-CoV-2. Analysis of community samples genetically related to CVD-DÜS-KITA-2021-0164 uncovered two cases (children attending the same preschool; Cases 40 and 41) that should have been assigned to the outbreak. Samples from outbreak CVD-DÜS-SCHULE-2021-0169 were also found to be genetically related (genetic distance $\leq 1$) to outbreak CVD-DÜS-KITA-2021-0164. An epidemiological connection between the two outbreaks was confirmed through a sequenced community case (Case 45) that was genetically related to both outbreaks; this case was found to be a family member (parent) of cases in both outbreaks. Analysis of symptom onset dates suggested that the infection chain connecting the two outbreaks started with the cases in CVD-DÜS-KITA-2021-0164. In total, the outbreak exhibited 2 sequencing-confirmed links to community cases that were also epidemiologically supported; 2 additional community cases (parents; Cases 47 and 48) that were epidemiologically linked to a case of CVD-DÜS-SCHULE-2021-0169 exhibited genetic distances $> 1$ to outbreak samples and were thus not classified as sequencing-confirmed linked samples. The automatically extracted symptom onset date of Case 38 exhibited an "off-by-one-month" error; true symptom onset date was on Day 5. This error was corrected for Figure 5 but not in the (automatically generated) visualization shown below.

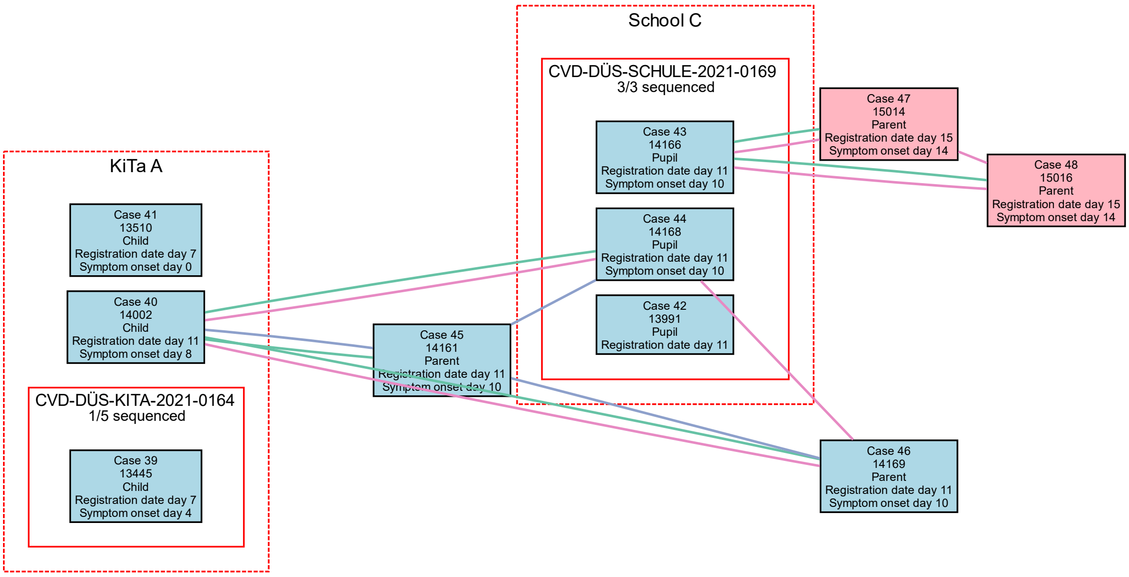


| | Z8027 | Z8053 | Z8328 | Z8280 | Z8277 | Z8290 | Z8281 | Z8284 | Z9393 | Z9392 |
|---|---|---|---|---|---|---|---|---|---|---|
| **Z8027** | 0 | 1 | 1 | 1 | 1 | 1 | 1 | 2 | 3 | 3 |
| **Z8053** | 1 | 0 | 0 | 0 | 0 | 0 | 0 | 1 | 2 | 2 |
| **Z8328** | 1 | 0 | 0 | 0 | 0 | 0 | 0 | 1 | 3 | 3 |
| **Z8280** | 1 | 0 | 0 | 0 | 0 | 0 | 0 | 1 | 3 | 3 |
| **Z8277** | 1 | 0 | 0 | 0 | 0 | 0 | 0 | 1 | 2 | 2 |
| **Z8290** | 1 | 0 | 0 | 0 | 0 | 0 | 0 | 1 | 3 | 3 |
| **Z8281** | 1 | 0 | 0 | 0 | 0 | 0 | 0 | 1 | 3 | 3 |
| **Z8284** | 2 | 1 | 1 | 1 | 1 | 1 | 1 | 0 | 4 | 4 |
| **Z9393** | 3 | 2 | 3 | 3 | 2 | 3 | 3 | 4 | 0 | 0 |
| **Z9392** | 3 | 2 | 3 | 3 | 2 | 3 | 3 | 4 | 0 | 0 |

**Genetic clusters**

1

2

Not sequenced

**Edge color legend**

Forward Contact Tracing

Backward Contact Tracing

sameAddress-sameName

sameAddress-differentName

Manual analysis

#### CVD-DÜS-SCHULE-2021-0175 and CVD-DÜS-SCHULE-2021-0180

CVD-DÜS-SCHULE-2021-0175 and CVD-DÜS-SCHULE-2021-0180 were two outbreaks registered at two different schools D and E, comprising a total of 5 (0 of which were sequenced) and 2 (1 of which was sequenced) cases, respectively. The outbreaks fell into the period of domination by the "Delta" variant of SARS-CoV-2. A potential link between the two outbreaks was suggested by the identification of a pair of siblings (Case 50 and Case 55) that were cases of two outbreaks (one sibling in each outbreak). An analysis of samples genetically related to the sequenced case in CVD-DÜS-SCHULE-2021-0180 uncovered two genetically related community cases (Cases 57 and 58), who were family members of another case (Case 54), who was found to be a pupil attending School D; furthermore, no epidemiological connections were found between either Case 57 or 58 and outbreak CVD-DÜS-SCHULE-2021-0180. A link between the two outbreaks was therefore also supported genetically based on the parsimonious assumption that the viral genomes associated with outbreak CVD-DÜS-SCHULE-2021-0175 were likely highly related to these of outbreak CVD-DÜS-SCHULE-2021-0180, and that Case 54 should have been classified as a case of outbreak CVD-DÜS-SCHULE-2021-0180. Furthermore, genetic similarity of the two outbreaks was also supported by the detection of another case (Case 60) that had a genetic distance of 1 to the cases linked to CVD-DÜS-SCHULE-2021-0175 and a genetic distance of 2 to the single sequenced case of CVD-DÜS-SCHULE-2021-0180. Analysis of symptom onset dates suggested that the infection chain connecting the two outbreaks started in School D. In total, the outbreaks exhibited 4 epidemiologically supported links to community cases that were supported by sequencing data.

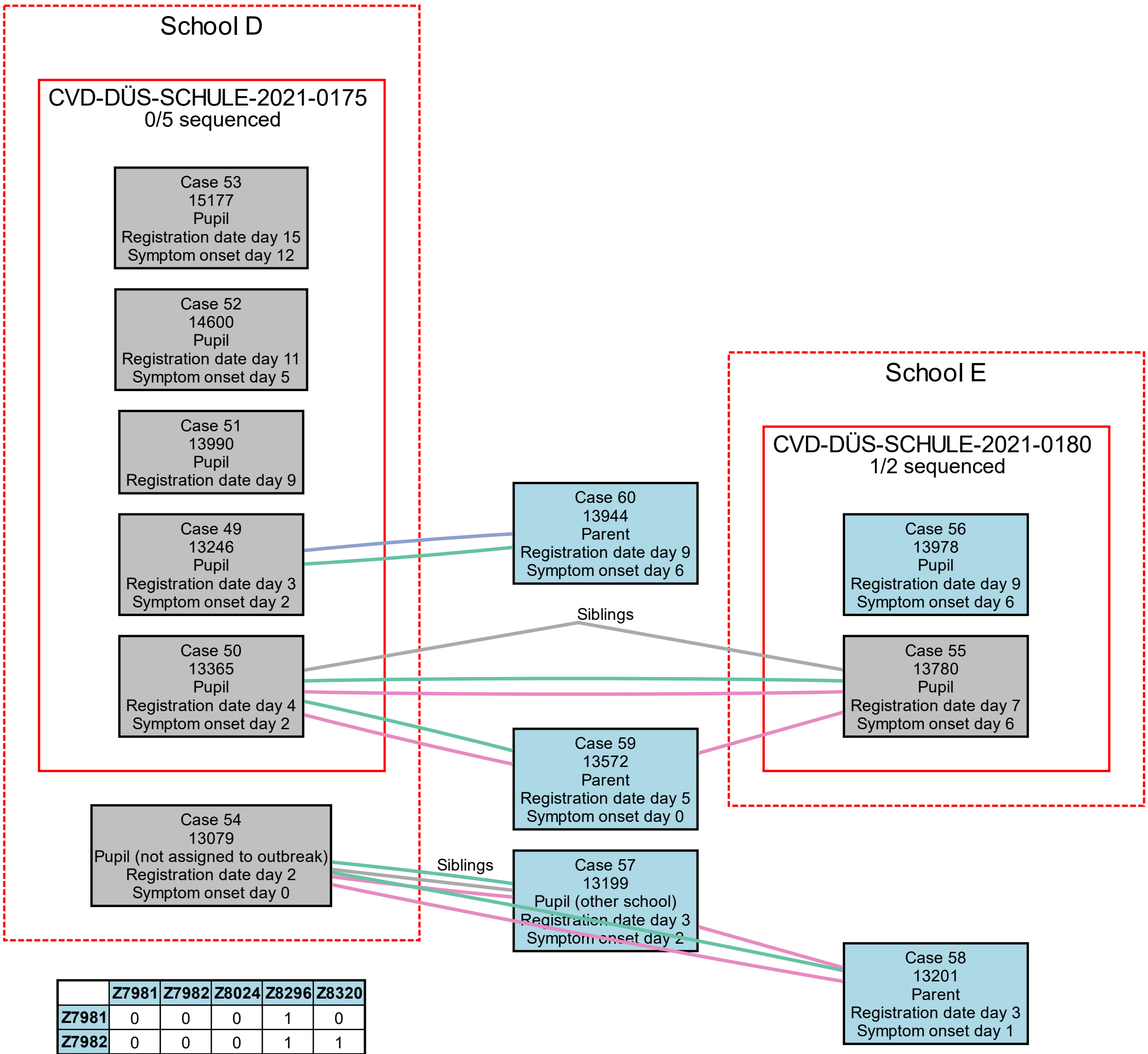


| | Z7981 | Z7982 | Z8024 | Z8296 | Z8320 |
|---|---|---|---|---|---|
| Z7981 | 0 | 0 | 0 | 1 | 0 |
| Z7982 | 0 | 0 | 0 | 1 | 1 |
| Z8024 | 0 | 0 | 0 | 1 | 0 |
| Z8296 | 1 | 1 | 1 | 0 | 2 |
| Z8320 | 0 | 1 | 0 | 2 | 0 |

**Genetic clusters**

- 1
- Not sequenced

**Edge color legend**

- Forward Contact Tracing
- Backward Contact Tracing
- sameAddress-sameName
- sameAddress-differentName
- Manual analysis

#### CVD-DÜS-SCHULE-2021-0182

CVD-DÜS-SCHULE-2021-0182 was a single-class school outbreak comprising 4 pupils at School F; of the 4 cases, 1 was sequenced. The outbreak fell into the period dominated by the "Delta" variant of SARS-CoV-2. Analysis of genetic similarities suggested that the four registered cases were likely infected with different viral strains, indicating that the outbreak was likely not clonal. While only one outbreak case (Case 61) was sequenced, sequencing data were also available for three contacts (Cases 65, 66, 67) of two additional outbreak samples (Cases 63 and 64). The viral sequences of the three contacts were found to be identical, and no epidemiological connection except for having had contact with cases of outbreak CVD-DÜS-SCHULE-2021-0182 was found between the case pair 65 and 66 (siblings) and case 67. This suggested (i) that the viral genotype of outbreak cases 63 and 64 was likely identical to the viral genotype of the 3 contacts; (ii) that the viral genotype of these cases was therefore likely different from that of the sequenced outbreak case 61 (genetic distance ~9), based on the observed genetic distances between case 61 and cases 65, 66, and 67; and, finally, (iii) that the outbreak was therefore likely not clonal. In addition, cases 65 and 66 were pupils in another class of School F, suggesting that these two cases should have been registered as outbreak cases and that CVD-DÜS-SCHULE-2021-0182 was not a single-class outbreak, but a multi-class outbreak. In total, the outbreak exhibited 1 epidemiologically and genetically supported link to another community case, who was a pupil at a different school.

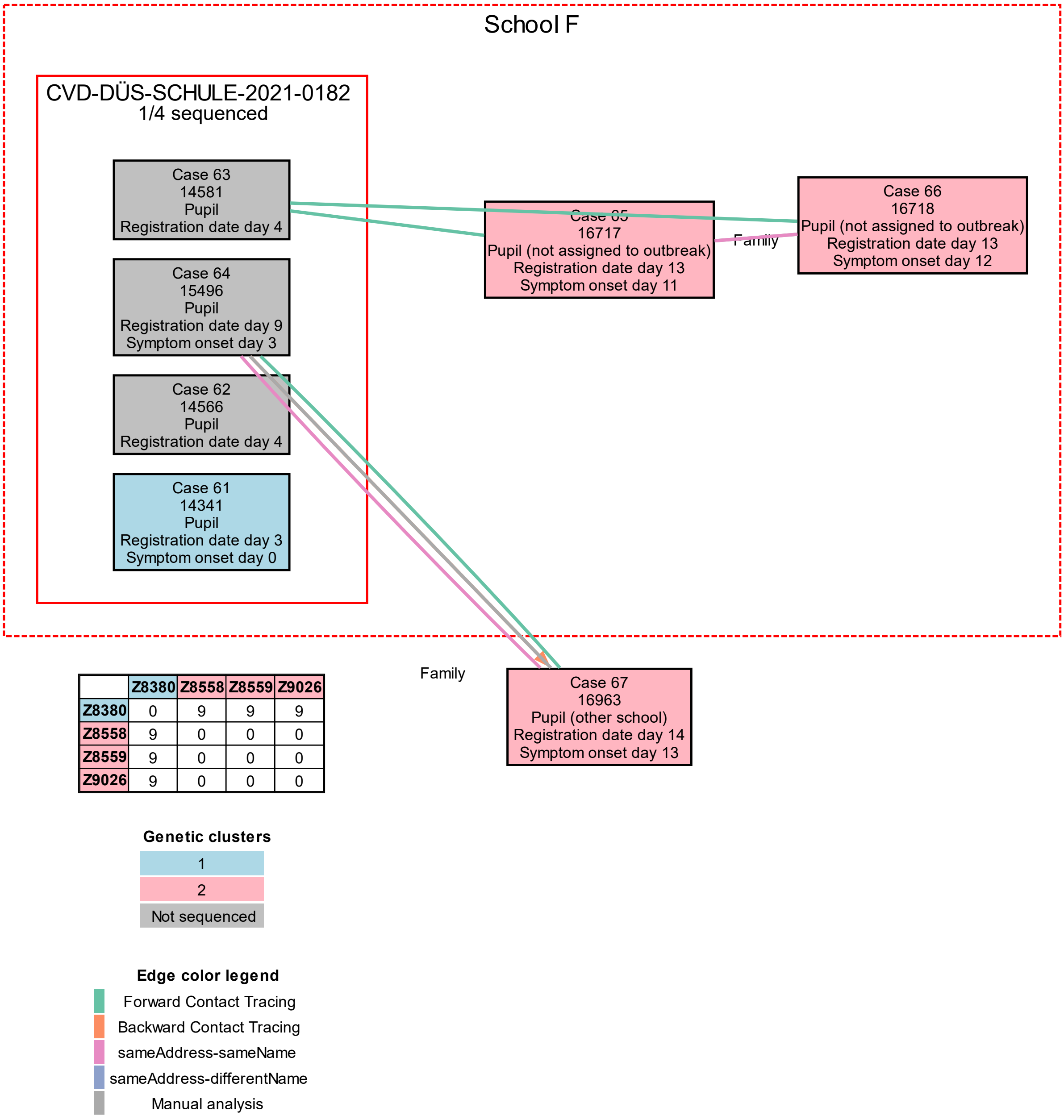


| | Z8380 | Z8558 | Z8559 | Z9026 |
|---|---|---|---|---|
| Z8380 | 0 | 9 | 9 | 9 |
| Z8558 | 9 | 0 | 0 | 0 |
| Z8559 | 9 | 0 | 0 | 0 |
| Z9026 | 9 | 0 | 0 | 0 |

#### CVD-DÜS-SCHULE-2021-0199

CVD-DÜS-SCHULE-2021-0199 was a multi-class school outbreak comprising 6 pupils at School G; of the 6 cases, 2 were sequenced. The outbreak fell into the period dominated by the "Delta" variant of SARS-CoV-2. Analysis of genetic similarities showed that one additional case (Case 73), a pupil at the same school who was also a registered contact of Case 71, should have been registered as a case of the outbreak. The outbreak exhibited 1 epidemiologically supported link to a community case (a family member of an outbreak case) with unambiguous genetic support (genetic distance 0) and another epidemiologically supported link to a community case (also a family member of an outbreak case) with genetic similarity 2.

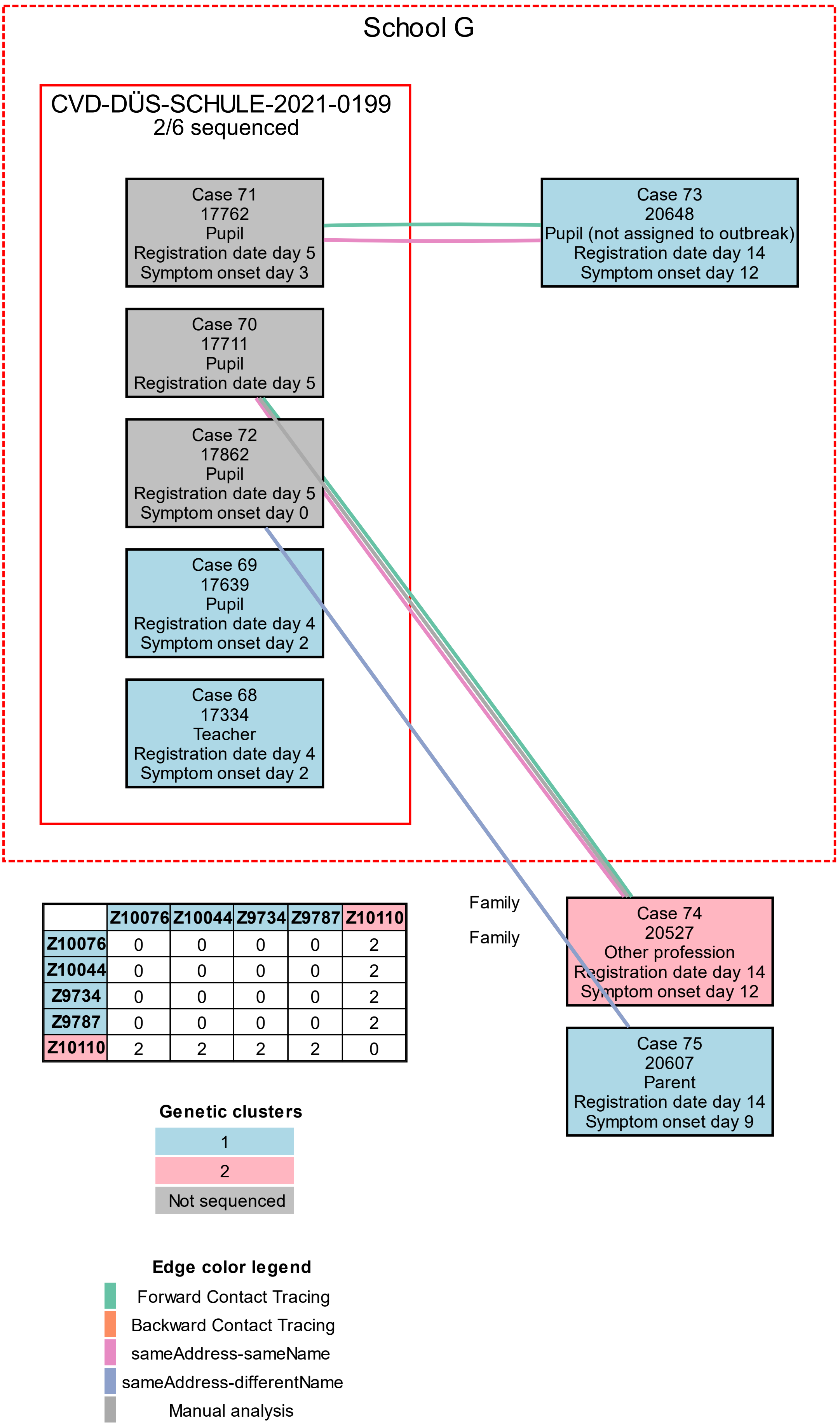


| | Z10076 | Z10044 | Z9734 | Z9787 | Z10110 |
|---|---|---|---|---|---|
| Z10076 | 0 | 0 | 0 | 0 | 2 |
| Z10044 | 0 | 0 | 0 | 0 | 2 |
| Z9734 | 0 | 0 | 0 | 0 | 2 |
| Z9787 | 0 | 0 | 0 | 0 | 2 |
| Z10110 | 2 | 2 | 2 | 2 | 0 |

#### CVD-DÜS-SCHULE-2021-0207

CVD-DÜS-SCHULE-2021-0207 was a single-class school outbreak comprising 8 pupils at School H; of the 8 cases, 4 were sequenced. The outbreak fell into the period dominated by the "Delta" variant of SARS-CoV-2. Analysis of genetic similarities identified a genetically linked missing outbreak case (Case 81); Case 81 was a pupil at the same school, but in a different class than the other outbreak cases, indicating that outbreak CVD-DÜS-SCHULE-2021-0207 was not a single-class outbreak, but a multi-class outbreak. The outbreak exhibited 2 epidemiologically and genetically supported links to community cases (parents of outbreak cases).

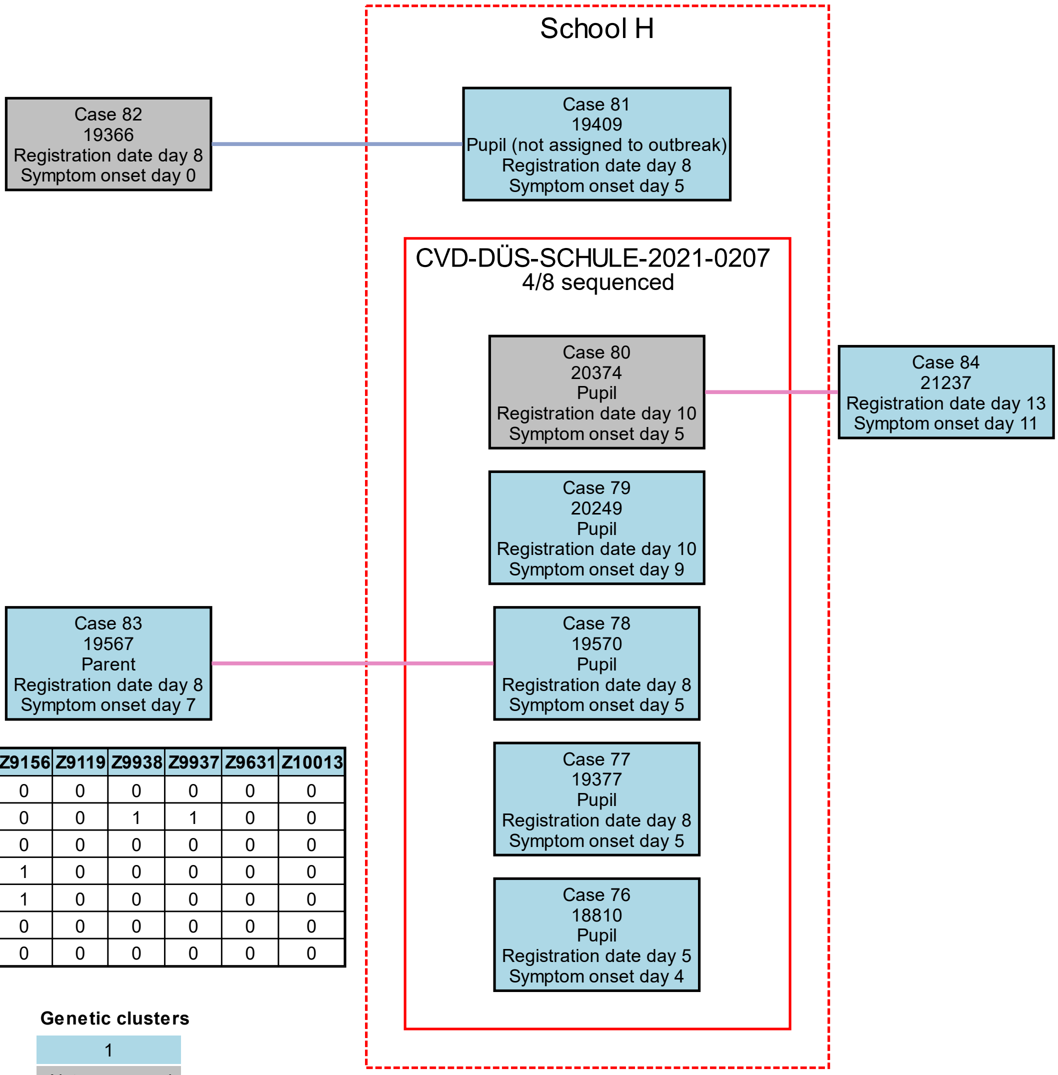


| | Z8908 | Z9156 | Z9119 | Z9938 | Z9937 | Z9631 | Z10013 |
|---|---|---|---|---|---|---|---|
| **Z8908** | 0 | 0 | 0 | 0 | 0 | 0 | 0 |
| **Z9156** | 0 | 0 | 0 | 1 | 1 | 0 | 0 |
| **Z9119** | 0 | 0 | 0 | 0 | 0 | 0 | 0 |
| **Z9938** | 0 | 1 | 0 | 0 | 0 | 0 | 0 |
| **Z9937** | 0 | 1 | 0 | 0 | 0 | 0 | 0 |
| **Z9631** | 0 | 0 | 0 | 0 | 0 | 0 | 0 |
| **Z10013** | 0 | 0 | 0 | 0 | 0 | 0 | 0 |

**Genetic clusters**

1
Not sequenced

**Edge color legend**

Forward Contact Tracing
Backward Contact Tracing
sameAddress-sameName
sameAddress-differentName
Manual analysis

#### CVD-DÜS-SCHULE-2021-0212, CVD-DÜS-SCHULE-2021-0231 and CVD-DÜS-SCHULE-2021-0251

CVD-DÜS-SCHULE-2021-0212, CVD-DÜS-SCHULE-2021-0231 and CVD-DÜS-SCHULE-2021-0251 were three separately registered single-class outbreaks at the same school (School J), comprising 10, 11, and 6 cases, of whom 0, 1, and 3 were sequenced, respectively. The outbreaks fell into the period of domination by the "Delta" variant of SARS-CoV-2. Analysis of genetic similarities showed that one case (Case 90) had likely been erroneously assigned to outbreak CVD-DÜS-SCHULE-2021-0251 (genetic distance to other outbreak cases $\geq 24$); in addition, the analysis showed that outbreaks CVD-DÜS-SCHULE-2021-0231 and CVD-DÜS-SCHULE-2021-0251 were genetically related; a link between the two outbreaks (over and above overlapping case registration dates and the fact that both outbreaks took place at the same school) was also supported by Case 86 (a case of CVD-DÜS-SCHULE-2021-0231) reported having had contact to Case 92 (a case of CVD-DÜS-SCHULE-2021-0251). Furthermore, analysis of community cases genetically related to the outbreaks uncovered two cases (Cases 95 and 96) that had not been assigned to any of the outbreaks but who were pupils in the same class as the cases of CVD-DÜS-SCHULE-2021-0212, suggesting that outbreak CVD-DÜS-SCHULE-2021-0212 was also connected to the other two outbreaks. In addition, analysis of genetically related community cases also detected a third case (Case 97), attending the same school but in a different class than the cases of the three investigated outbreaks. Taken together, these findings suggested that CVD-DÜS-SCHULE-2021-0212, CVD-DÜS-SCHULE-2021-0231 and CVD-DÜS-SCHULE-2021-0251 were not three separate single-class outbreaks, but belonged to a larger multi-class outbreak at School J; and that a total of 3 additional cases should have been registered as outbreak cases. The outbreak exhibited 7 epidemiologically and genetically supported links to community cases; 3 of the 7 linked community cases were parents of outbreak cases; 2 of the 7 linked community cases were pupils at another school; and 2 of the 7 linked community cases were only identified via the analysis of genetically related samples.

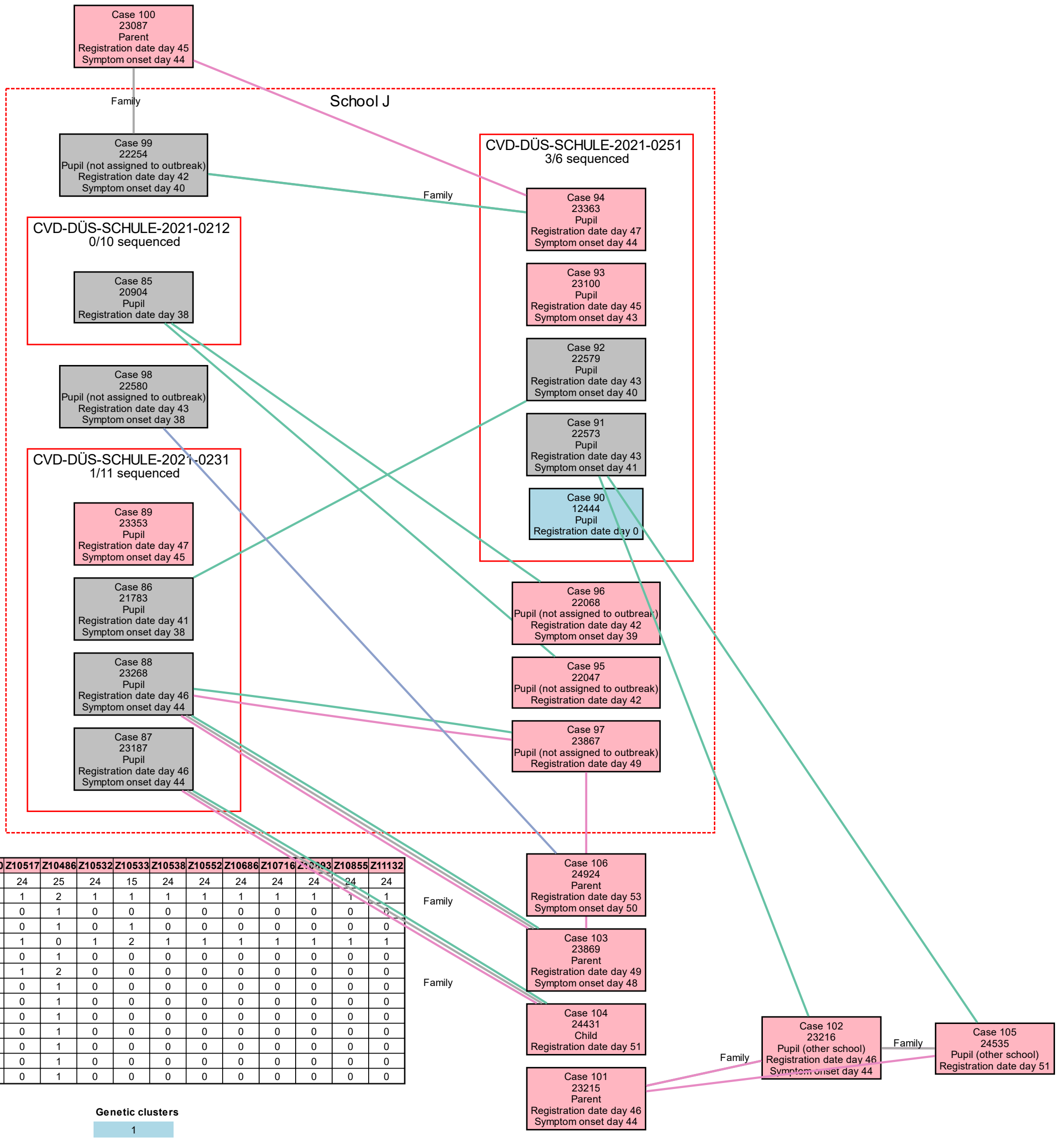


| | Z7676 | Z10271 | Z10390 | Z10517 | Z10486 | Z10532 | Z10533 | Z10538 | Z10552 | Z10686 | Z10716 | Z10893 | Z10855 | Z11132 |
|---|---|---|---|---|---|---|---|---|---|---|---|---|---|---|
| **Z7676** | 0 | 25 | 24 | 24 | 25 | 24 | 15 | 24 | 24 | 24 | 24 | 24 | 24 | 24 |
| **Z10271** | 25 | 0 | 1 | 1 | 2 | 1 | 1 | 1 | 1 | 1 | 1 | 1 | 1 | 1 |
| **Z10390** | 24 | 1 | 0 | 0 | 1 | 0 | 0 | 0 | 0 | 0 | 0 | 0 | 0 | 0 |
| **Z10517** | 24 | 1 | 0 | 0 | 1 | 0 | 1 | 0 | 0 | 0 | 0 | 0 | 0 | 0 |
| **Z10486** | 25 | 2 | 1 | 1 | 0 | 1 | 2 | 1 | 1 | 1 | 1 | 1 | 1 | 1 |
| **Z10532** | 24 | 1 | 0 | 0 | 1 | 0 | 0 | 0 | 0 | 0 | 0 | 0 | 0 | 0 |
| **Z10533** | 15 | 1 | 0 | 1 | 2 | 0 | 0 | 0 | 0 | 0 | 0 | 0 | 0 | 0 |
| **Z10538** | 24 | 1 | 0 | 0 | 1 | 0 | 0 | 0 | 0 | 0 | 0 | 0 | 0 | 0 |
| **Z10552** | 24 | 1 | 0 | 0 | 1 | 0 | 0 | 0 | 0 | 0 | 0 | 0 | 0 | 0 |
| **Z10686** | 24 | 1 | 0 | 0 | 1 | 0 | 0 | 0 | 0 | 0 | 0 | 0 | 0 | 0 |
| **Z10716** | 24 | 1 | 0 | 0 | 1 | 0 | 0 | 0 | 0 | 0 | 0 | 0 | 0 | 0 |
| **Z10893** | 24 | 1 | 0 | 0 | 1 | 0 | 0 | 0 | 0 | 0 | 0 | 0 | 0 | 0 |
| **Z10855** | 24 | 1 | 0 | 0 | 1 | 0 | 0 | 0 | 0 | 0 | 0 | 0 | 0 | 0 |
| **Z11132** | 24 | 1 | 0 | 0 | 1 | 0 | 0 | 0 | 0 | 0 | 0 | 0 | 0 | 0 |

**Genetic clusters**

- 1
- 2
- Not sequenced

**Edge color legend**

- Forward Contact Tracing
- Backward Contact Tracing
- sameAddress-sameName
- sameAddress-differentName
- Manual analysis
